# Estimation of age-stratified contact rates during the COVID-19 pandemic using a novel inference algorithm

**DOI:** 10.1101/2022.01.12.22269157

**Authors:** Christopher M. Pooley, Andrea B. Doeschl-Wilson, Glenn Marion

**Affiliations:** Biomathematics and Statistics Scotland, James Clerk Maxwell Building, The King’s Buildings, Peter Guthrie Tait Road, Edinburgh, EH9 3FD, UK; The Roslin Institute, The University of Edinburgh, Midlothian, EH25 9RG, UK; Scottish COVID-19 Response Consortium, www.gla.ac.uk/scrc/

**Keywords:** COVID-19, Bayesian inference, contact matrix, reproduction number, ABC, MBP

## Abstract

Well parameterised epidemiological models including accurate representation of contacts, are fundamental to controlling epidemics. However, age-stratified contacts are typically estimated from pre-pandemic/peace-time surveys, even though interventions and public response likely alter contacts. Here we fit age-stratified models, including re-estimation of relative contact rates between age-classes, to public data describing the 2020-21 COVID-19 outbreak in England. This data includes age-stratified population size, cases, deaths, hospital admissions, and results from the Coronavirus Infection Survey (almost 9000 observations in all). Fitting stochastic compartmental models to such detailed data is extremely challenging, especially considering the large number of model parameters being estimated (over 150). An efficient new inference algorithm ABC-MBP combining existing Approximate Bayesian Computation (ABC) methodology with model-based proposals (MBP) is applied. Modified contact rates are inferred alongside time-varying reproduction numbers that quantify changes in overall transmission due to pandemic response, and age-stratified proportions of asymptomatic cases, hospitalisation rates and deaths. These inferences are robust to a range of assumptions including the values of parameters that cannot be estimated from available data. ABC-MBP is shown to enable reliable joint analysis of complex epidemiological data yielding consistent parametrisation of dynamic transmission models that can inform data-driven public health policy and interventions.

## Introduction

Dynamic mathematical models, such as the Susceptible-Infectious-Recovered (SIR) transmission model and its numerous variants, have been widely applied over many decades to enhance understanding of infectious disease spread and control in populations (see *e*.*g*. [1]). They have been particularly prominent in guiding public health responses to the COVID-19 pandemic [2], and are increasingly seen as important tools for causal inference in epidemiology [3]. Model results used to inform public health decisions must be defensible, requiring models to be informed by, and be consistent with, the widest range of suitable data sources available [4]. However, as discussed below, the COVID-19 pandemic has shown that tools to enable reliable and routine inference for sufficiently realistic models of disease dynamics from multiple sources of data are currently lacking. Here we address this gap by introducing a novel algorithm that enables efficient inference for complex compartmental models, illustrating its potential by fitting an age-structured national-scale model to multiple data sources (demographic, operational and survey data) arising from the COVID-19 epidemic in England 2020-21. This enables simultaneous inference of many key parameters describing the outbreak, and the benefits of such an integrated approach are exemplified by updating pre-pandemic age-stratified contact rates to account for relative differences in contact between age groups.

Fitting complex compartmental models to multiple data sources, with required uncertainty estimates for the key model parameters, comes with computational challenges. Bayesian inference for stochastic models of disease dynamics has been implemented using a range of methodologies that provide exact or approximate approaches to drawing samples from the posterior distribution. Data augmentation approaches, typically combined with Markov chain Monte Carlo (MCMC), offer exact, albeit computationally expensive inference, but require explicit expressions for the complete-data likelihood (see *e*.*g*. [5] for an application to an individual level model of COVID-19). In contrast, approximate Bayesian computation (ABC) [6], including frequently used sequential Monte Carlo (SMC) versions [7], is likelihood-free, requiring only the ability to simulate from the model, but typically draws samples from a poor approximation to the posterior. Other SMC algorithms, such as particle MCMC (pMCMC) [8] are also likelihood-free, and in principle offer exact samples from the posterior by developing unbiased estimators for the likelihood via simulation. In many cases these perform well, however, their computational cost typically scales poorly with the dimensionality of the data [9], a serious drawback when applying to large datasets such as those used for COVID-19 here. A relevant example is [10], which applies pMCMC to a stochastic age-structured non-spatial model used to look at variation in the COVID-19 reproduction number separately for different regions of England (for comparison this contained 26 parameters and fitted around 1300 data points with an over-dispersed observation model to aid computational speed^1^ [11]). As with others studies, a contact matrix based on the pan-European POLYMOD study [12] was assumed, so limiting the number of parameters to be estimated, and the model was fitted using non-age-structured (*i*.*e*. aggregated) data. Adding in age structure greatly increases the number of individual observations (by a factor of ∼17 here), and we found this makes applying a pMCMC algorithm computationally impractical (results not shown). This motivated development of the new ABC-MBP algorithm described below (see Methods) that combines model-based proposals (MBPs), derived from data-augmentation MCMC, with ABC methods.

Quantifying the effective contact structure of a population (*i*.*e*. how different groups intermix and transmit infection during an epidemic) is central to epidemiological modelling. For diseases with age as a risk factor, such as COVID-19, understanding how such contacts vary with age is vital because it helps quantify the likely impact of age targeted restrictions, *e*.*g*. school closures, care home restrictions or closing night time entertainment. Typically simulation studies are used to assess this impact [13, 14], but accuracy is limited by the degree to which model parameters can correctly be identified given the available data. Approximations of social mixing patterns have been constructed based on wearable proximity sensors [15, 16] or self-recording of contacts [17]. In terms of scale, two studies in the latter category stand out. Firstly, the 2005-6 POLYMOD study [12, 18] recruited 7,290 participants across eight European countries and asked each participant to fill in a diary that recorded the contacts they made on one particular day, along with estimated ages of those contacts. Secondly, the 2017-18 BBC Pandemic study [19] recruited 36,000 volunteers from the UK who used a self-reporting mobile phone app to inform about contacts made. Based on these surveys each study published (pre-pandemic) estimates for the so-called “contact matrix” **C**^**0**^, which captures the social mixing patterns between age groups [20] that have been widely used to model transmission dynamics of COVID-19. In this paper we infer the time-varying reproduction rate *Rt* to quantify the net impact of societal response (lockdowns, social distancing *etc*.) during the pandemic, and, importantly, estimate modifications to **C**^**0**^ that quantify relative changes in mixing between different age groups.

Age stratification in observed COVID-19 cases may be driven by age related variations in susceptibility, contacts, infectivity, probability of being asymptomatic and time spent in latent, infectious and other disease states. This complicates estimation of age-related changes in contacts, but [21] show that viral load does not vary greatly with age or between hospitalised and other cases and thus here we assume infectivity is constant across all age classes and hospitalised and non-hospitalised cases. Furthermore, this study found similar viral load trajectories for children and adults and so we also assume residency times in disease states are not age dependent. In contrast, previous studies have shown age-related differences in contacts [12, 18, 19], susceptibility and proportion of clinical (symptomatic) cases [22] as modelled here (see Methods). We show that use of PCR and seroprevalence survey results, in addition to case reports, allows estimation of age-related fractions of asymptomatic infections alongside *Rt* and age-stratified hospitalisation rates and deaths from hospitals. However, estimation of age-related susceptibility and (relative) contact rates, are confounded and therefore we estimate them in two separate analyses, in each case leading to models that perform similarly and are better predictors than models that don’t account for such heterogeneity. Results (see below) suggest relative overall contact rates are reduced and age-related contact patterns are different to pre-pandemic patterns with greater relative contact rates by younger age classes. We provide an estimate for these changes assuming susceptibility does not vary with age. Consistent with increasing case fatality rate with age, others have found increasing susceptibility with age [22] suggesting our estimates of increased relative contacts by younger groups is conservative. In contrast, our estimation of age-dependent susceptibilities (assuming pre-pandemic contact patterns) are not consistent with increasing age-related susceptibility, further indicating that pre-pandemic relative mixing matrices require re-estimation during the pandemic. In the main text we therefore focus on estimation of age-related contact rates, and defer a comparison to the age-dependent susceptibility model until the discussion.

## Methods

We first describe the model, give a brief description of the data, and then outline the new ABC-MBP Bayesian inference algorithm used to infer model parameters.

### The compartmental model

Figure 1 shows the non-spatial age-stratified compartmental model, representing the number of individuals in a given age-class and disease category. The population is assumed to be stratified into 18 age groups indexed by *a*: the first 16 represent five year age bands (“0-4”, “5-9”,…,”75-79”), then “80+” for all individuals 80 and over, and, finally, a group for care home residents denoted by “CH”^2^. Once an individual in the susceptible compartment S becomes infected by COVID-19 they enter the exposed E compartment where they spend an exponentially distributed residency time *m*E. A branching probability 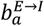 determines whether they then enter an infectious I compartment^3^ or an asymptomatic A compartment. Individuals go on to trace a path in Fig. 1, undergoing exponentially distributed residency times in the various compartments and choosing transitions based on branching probabilities when alternative routes exist, until they eventually reach the terminal recovered R or dead D compartments. Note, the branching probabilities in this diagram all depend on age group *a*. These probabilities will later be estimated from data.

**Figure 1:**
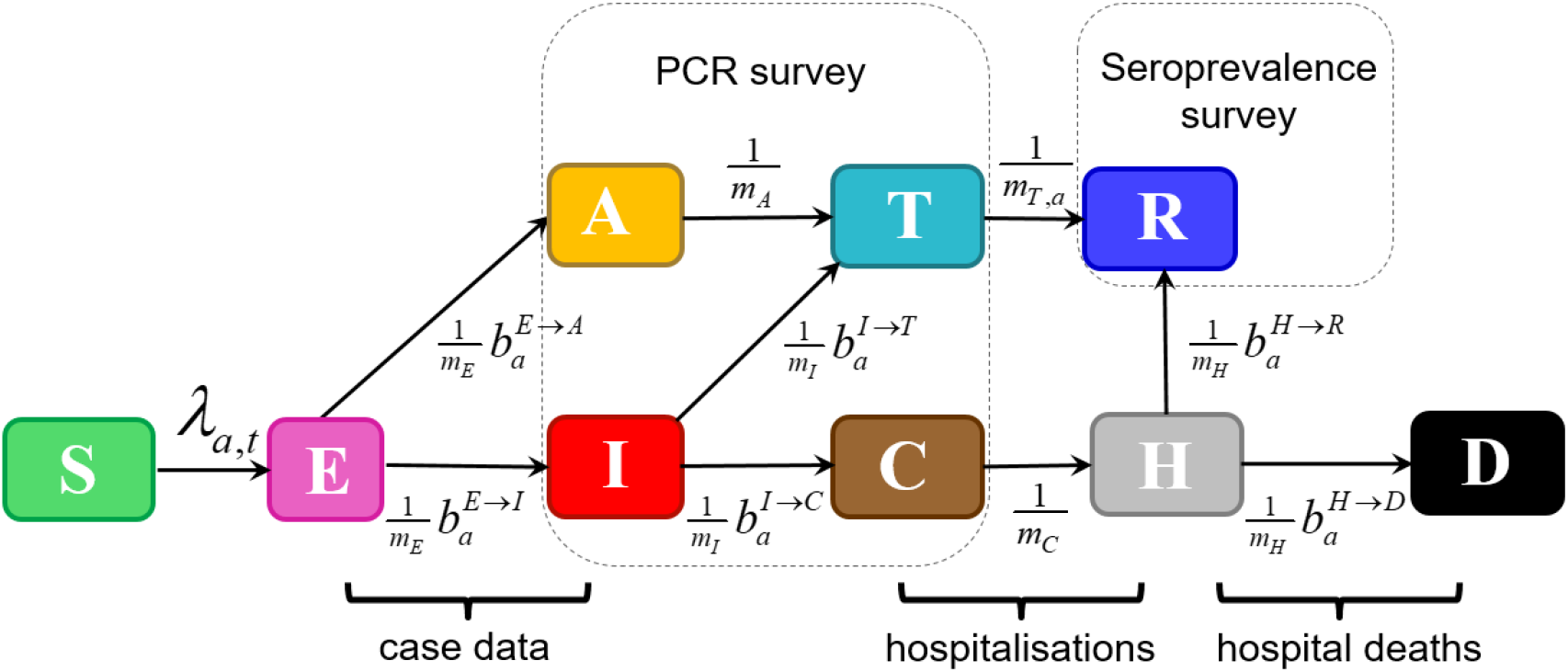
The compartmental model. The compartments are defined as follows: S: Susceptible, E: Exposed, A: Asymptomatic, I: Infectious, T: PCR test-sensitive but non-infectious, C: Infected but non-infectious (due to self-isolation), H: Hospitalised (or potentially care home in the case of care home residents), R: Recovered, and D: Dead. Variable *m*_*c*_ gives the mean residency time in a compartment *c* (with individual residency times exponentially distributed about this mean) and 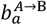 gives the branching probability for an individual in age group *a* going from compartment A to B (note, these probabilities are constrained to add up to one when leaving a given compartment). The term *λ*_*a,t*_ refers to the time-dependent force of infection acting on individuals in age group *a*. The annotations also indicate how data from the COVID-19 epidemic in England inform inference. Survey data are taken to provide an unbiased estimate of proportions of the total population in the sum of classes A, I, T, C (PCR survey) and R (seroprevalence). Operational data is used to inform transition rates E→I (test positive cases for second pandemic wave), C→H (hospitalisations) and H→D (deaths in hospitals and care homes).

The force of infection *λ*_*a,t*_ in Fig. 1 defines the probability per unit time that an individual becomes infected:

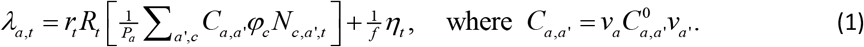

The various terms in this expression are as follows:

#### Reproduction number *R*_*t*_

This time-varying quantity is defined as the expected number of secondary cases directly generated by a case in a population in which nearly all individuals are susceptible. Estimation of this quantity relies on the approach taken by Diekmann *et* al. [23], in which *R*_*t*_ is calculated from the highest eigenvalue of the next generation matrix (see Appendix A in the Supplementary Material for further details). *R*_*t*_ is incorporated into the model by means of a piecewise linear spline. The breakpoints for this spline are placed at roughly two-week intervals (with additional points near to lockdowns where it may be expected that *R*_*t*_ changes rapidly). The quantity *r*_*t*_, which links *R*_*t*_ to the more conventionally used transmission rate, is derived from other model parameters (see Appendix A).

#### Age group population *P*_*a*_

The total population size in age group *a*.

#### Pre-pandemic contact matrix C^0^

This square matrix captures disease relevant contacts between individuals in the 18 age groups. In particular, the element 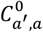 gives the average number of daily contacts for an individual in age group *a* with individuals in age group *a*’. Specification of this fixed matrix is based on data from the BBC Pandemic study [19] (details are provided in Appendix C). A pictorial representation of **C**^**0**^ is given in Fig. 2(a). Note the tri-diagonal structure results from frequent inter-generational contacts^4^.

**Figure 2:**
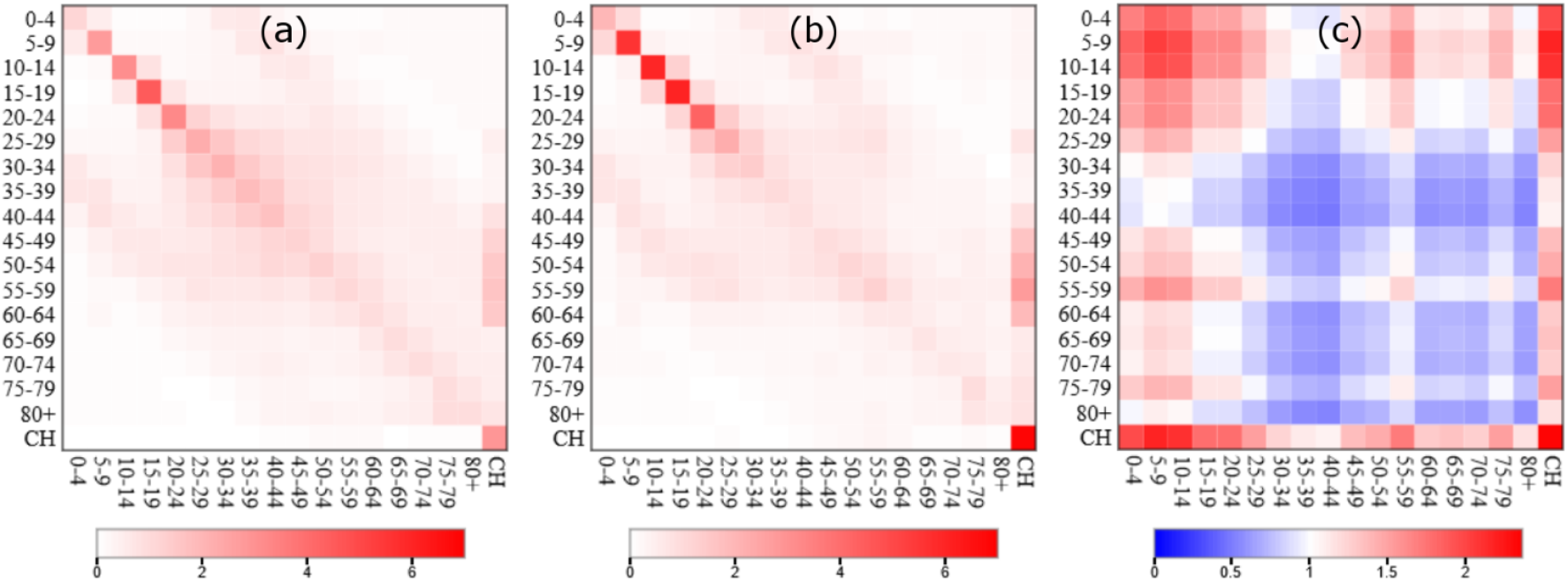
The contact matrix. (a) The pre-pandemic matrix **C**^**0**^ based on the BBC Pandemic study [8] (see Appendix C). For an individual represented by an age group on the *x*-axis this shows the estimated numbers of daily contacts they make with individuals in the same and different age groups on the *y*-axis (darker red colours indicate more frequent contacts). (b) The inferred age-adjusted contact matrix **C** based on COVID-19 data. (c) The factor difference in **C** over **C**^**0**^ (red colours indicate an increase and blue colours a decrease).

#### Age contact factors *v*

Each element of this vector corresponds to a specific age group and multiplies the corresponding columns and rows of the contact matrix **C**^**0**^. It effectively allows the model to enhance or reduce the relative rate of contacts for different age groups in order to generate an “age adjusted” estimate **C** for the contact matrix. These contact factors are constrained to have a weighted average of one, *i*.*e*.

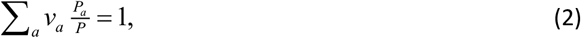

where *a* sums over all age groups and *P* is the total population size^5^.

#### Infectivity *φ*_*c*_

This sets the relative compartmental infectivity. For simplicity we assume only two infectious compartments: I (for which *φ*_I_=1) and A (for which *φ*_A_=0.55, as estimated in Appendix K). All other compartment have *φ*_c_=0.

#### Subpopulation size *N*_*c,a,t*_

The total number of individuals in compartment *c* and age group *a* at time *t*.

#### External force of infection *η*_*t*_

The time-varying probability per unit time an individual becomes infected from a source outside of England (see Appendix D for how this is derived from flight [24] and global COVID-19 death data [25]). For convenience this term is divided by a factor *f*=10^5^ such that it is expressed in units of infections per 100k individuals.

Three important points can be made about Eq.(1): Firstly, it corresponds to a non-spatial model, equivalent to assuming small spatial variation across England for demographic makeup, the contact matrix and external force of infection^6^. Secondly, individuals in all age groups are taken to be equally susceptible to disease (this is revisited later). Lastly, whilst age contact factors ***v*** adjust the relative rates of contact for different age groups compared to the pre-pandemic matrix **C**^**0**^, it is important to remember that the overall effective contact matrix is modulated by the reproduction number:

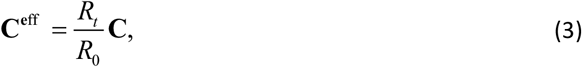

where *R*_*0*_ is the basic reproduction number. This accounts for changes in the overall rate of contacts (through government interventions and modification in social behaviour) as well as measures aimed at blocking transmission on contact (*e*.*g*. wearing face masks, washing hands, social distancing)^7^. Hence, time variation in *R*_*t*_ is effectively a proxy for time variation in the overall rate of effective contacts.

The stochastic dynamics of the model presented above are approximated by a *τ*-leaping algorithm [26] with a fixed time step of *τ*=0.5 days (see Appendix E for details).

#### Data sources

The time period used for the analysis in this study is between 1^st^ January 2020 (before the start of the COVID-19 pandemic in England) and 9th June 2021 (just after the second wave of infections). Figure 1 provides a synoptic illustration of how demographic, operational and survey information are interpreted in the context of the compartmental model to enable inference. Further details and information on how these datasets have been prepared for analysis are given in Appendix F (plots of the raw data can be seen by the black lines in the posterior Supplementary Results, with age-aggregated results in Fig. 6). In total, this data consists of almost 9000 individual observations.

#### Bayesian inference

Collectively the model parameters are referred to as *θ* (these determine the movement of individuals through the compartmental model in Fig. 1). When simulating from the model, individuals start in the susceptible S compartment^8^ and transitions between the compartmental states result in the subpopulations *p*_*a,c,t*_ within age group *a* and compartment *c* changing as a function of time *t*. These dynamics are collectively referred to as the system “state” (actually a set of states for each point in time), and denoted *ξ*. In reality *ξ* is unknown, and from a Bayesian perspective is considered a set of latent variables to be estimated.

The data is collectively denoted *y*. Application of Bayes’ theorem implies that the posterior probability distribution π(*θ,ξ*|*y*) is given by

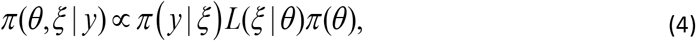

where various terms in this expression are:

#### The observation model π(*y*|*ξ*)

This gives the probability of the data given a system state based on statistical characterisation of the relevant observation processes. Often writing down a true observation model is complicated by the fact that we may not know the error associated with a set of measurements and the observation probability for a set of measurements down a time series may be highly correlated.

Rather than attempt this, this paper follows an approximate Bayesian computational (ABC) approach which relies on introducing a measure of fit between the data *y* and the state *ξ* called the “error function” (EF). A small error function implies a close correspondence between *ξ* and *y*. For ABC the observation model is defined to be non-zero when the error function is less than some specified cut-off EF_cutoff_:

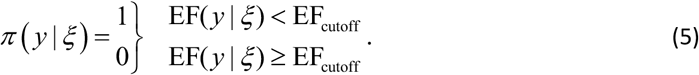

The challenge for inference algorithms is to reduce this cut-off as much as possible to ensure a close correspondence between the system state *ξ* and the observed data *y*. How this is implemented in practice is discussed in the next section.

Several possibilities exist for the choice of EF^9^, but the following proved effective for the problem in this paper:

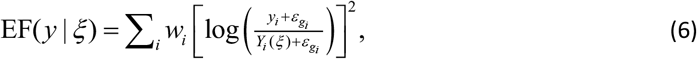

where the sum *i* goes over all individual measurements made on the system (*i*.*e*. each of the data points on the age-stratified time series for cases, deaths, hospital admissions and the CIS results), *y*_*i*_ is the value of the *i*th measurement and *Y*_*i*_ is the equivalent value derived from the state *ξ*. The small constant 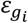 (dependent on the time series *g*_*i*_ to which *i* belongs) is introduced to ensure validity even when *y*_*i*_ or *Y*_*i*_ are zero^10^.

When *y*_*i*_ and *Y*_*i*_ are equal the contribution to the error function in Eq.(6) is zero. When they are unequal the contribution is always positive (and for small deviations, close to the square of the fractional change in the quantity). The weights *w*_*i*_ in Eq.(6) set the relative importance of different measurements. In particular, they place greater weight on larger observed values^11^ and overall give an approximately equal weighting to each of the time series in the data^12^ (see Appendix I for details).

#### The latent process likelihood *L*(*ξ*|*θ*)

This gives the probability of simulating a state *ξ* given a set of model parameters *θ*. The MBPs used below are “likelihood-free”, so this quantity is not explicitly calculated.

#### The prior π(*θ*)

This captures the state of knowledge regarding parameter values before data *y* is considered. Prior specifications are given for all model parameters in Table R1 of the posterior Supplementary Results (see Appendix J for further explanation). Parameters for which the data is uninformative are fixed to plausible values taken from the literature (see Appendix K)^13^.

#### Approximate Bayesian Computation with model-based proposals (ABC-MBP)

In this section we explain the ABC-MBP approach used to perform inference. Note, a “particle” refers to the combination of a set of model parameters *θ* and a system state *ξ*. The algorithm below iterates over a series of generations *G*, and with each generation it successively improves the posterior accuracy:

1. **Initialisation** – A generation index *g* is set to 1. *P* particles^14^, indexed by *p*, are sampled from the model. This is achieved by first sampling 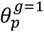 from the prior π(*θ*) and then simulating 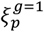 from the model (using the *τ*-leaping algorithm described in Appendix E). The initial error function cut-off is set to infinity 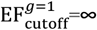.
2. **Particle culling and resampling** – An error function cut-off 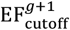 is set such that exactly half the particles have an error function above this value and half below^15^. Particles with error functions below are directly copied to create new particles in the next generation 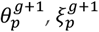. On the other hand, particles with error function above 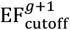 are discarded and replaced by a randomly selected particle from one of those whose error function is below this limit. The generation index *g* is incremented.
3. **Particle mixing –** To avoid degeneracy each particle undergoes a series of MBPs collectively referred to as an “update” (see appendix L for details) [9, 27]. These allow particles to explore parameter and state space subject to the condition that their error function must not exceed 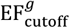. MBPs are the key novelty in this algorithm because they generate efficient MCMC mixing^16^ by making joint proposals in parameter and state space possible whilst at the same time restricting large changes in the error function (this is in contrast to standard ABC schemes that rely on simulation^17^).
4. **Jump to 2 if *g* is less than *G***. Particles in the final generation are used to approximate the posterior distribution.

This algorithm guarantees the error function cut-off monotonically decreases towards zero as the generation number increases. As it does so, particle states are forced to lie closer and closer to the data and posterior estimates similarly approach the true posterior. As the generation number *g* gets higher, however, so more and more MBPs are required in step 3 to avoid particle degeneracy. Therefore, for reasons of computationally practicality, the total generation number *G* is usually considered sufficiently large when posterior distribution estimates have apparently converged to steady state values (see posterior Supplementary Results Fig. R2).

## Results

### Inference protocol and validation

Inference was performed using the data and ABC-MBP approach outlined (see Methods). This was implemented using an open source software package called BEEPmbp (see Appendix M for details). To ensure robustness against lack of convergence, or multimodality in the posterior, each analysis was completed by running BEEPmbp *K*=16 times (chosen to check if randomly initialised starting states converge on the same posterior solution), with each run using *P*=16 particles (sufficiently large to adequately represent the posterior and avoid becoming stuck in metastable states) iterated over *G*=350 generations (large enough to get good estimates for parameters, see Fig. R2 in the posterior Supplementary Results). The results presented below are based on the *K***×***P*=256 samples generated over those runs^18^. The CPU time in total (over all 16 runs) was 4.5 hours when running on 256 cores. Whilst computationally expensive for challenging problems like this one, ABC-MBP is highly parallelisable and can be run relatively quickly given sufficiently powerful computing resources. Inference on representative simulated data correctly identified true model parameters demonstrating the validity of the algorithm for the problem at hand (see Appendix N for details).

### Analysis of England 2020-21 COVID-19 outbreak data

The posterior Supplementary Results provide details of fitting the model (Fig.1) with equation (1) to the COVID-19 outbreak data as described above, including: inferred model parameter distributions (Table R1), estimates for the contact matrix **C** (Table R2), graphs showing fits between raw data and inferred states (Fig. R1)^19^, and convergence down generations (Fig. R2). Here we summarise the key results.

#### Posterior mean estimates for the contact matrix C

are shown in Figure 2(b) with factor differences between **C** and **C**^**0**^ in Fig. 2(c). Based on these, noticeable differences in the contact patterns occur in the majority of age groups. The substantially elevated mixing amongst younger age groups can be understood by examining Fig. 3(a), which shows posterior distributions for the age contact factors (vector ***v*** in Eq.(1)). For younger age groups (aged 24 and below) the values exceed one indicating increased contacts relative to what would be expected from **C**^**0**^. We note this is broadly consistent with [5], who find increased force of infection for those under 60 versus older individuals. Given the reduced severity of symptoms in these age groups it seems plausible that this results from higher contact rates, rather than increased rates of shedding. The pattern in older age groups is somewhat more complex; group 40-44 was found to reduce contacts relative to **C**^**0**^ the most, but interestingly relative rates go up again until the 55-59 age group before reducing for older individuals. It should be noted that, in contrast to other age-groups, absence of data on pre-pandemic contact rates for care-home residents means the estimate for ***v***_***CH***_ (Fig.3(a)) should not be interpreted as an increase in contact rate. Rather, it is simply a reflection of the fact that contact rates in care homes are much higher than the rest of the population^20^ (due to being communal environments with frequent contacts between residents and staff) which has led to a disproportionate number of cases, and sadly deaths, compared to the elderly in the general population.

**Figure 3:**
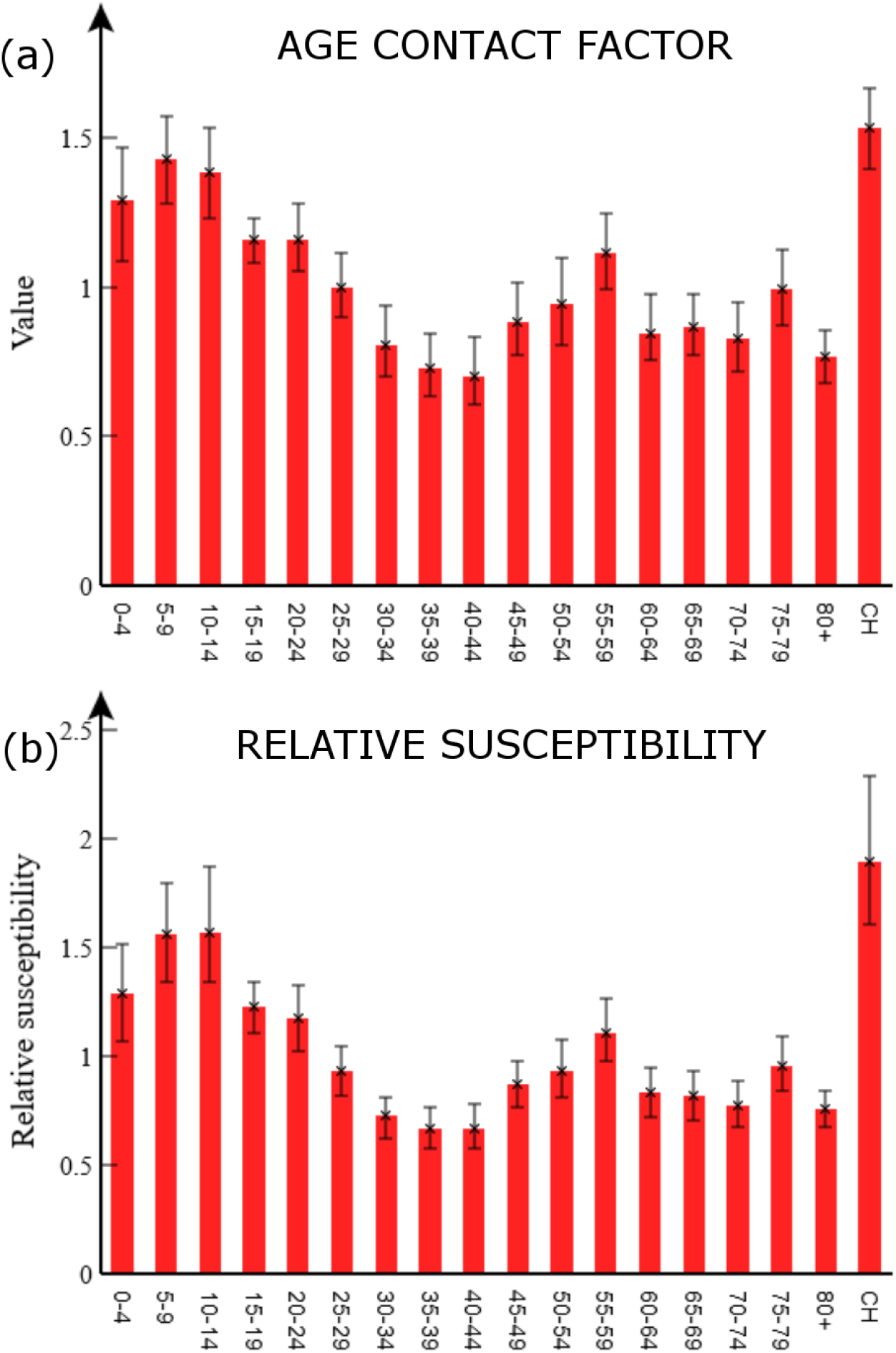
Age contact factors and variation in susceptibility. This shows two alternative models used to fit the data: (a) Age contact factors ***v*** modify the pre-pandemic contact matrix **C**^**0**^ for different age groups in the population (see Eq.(1)). (b) Age groups are given a relative susceptibility to acquiring infection (see Eq.(7)). The error bars show 95% credible intervals.

#### Time variation in the reproduction number

Figure 4(a) shows the posterior distribution for *R*_*t*_ with important events on the timeline shown by vertical lines (see Appendix O and [28]). As mentioned in Eq.(3), this is a proxy for the overall effective contact rate in the population. On the other hand the effective reproduction number 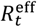 (see Appendix A), as shown in Fig. 4(b), tracks disease progression (above one implies COVID-19 infections are increasing)^21^. These estimates capture the net impact of the many factors that determined the epidemic trajectory. The first lockdown clearly had a huge impact on reducing 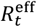 below one and ending the first wave. A gradual increase over the summer and autumn of 2020 (coinciding with restrictions being lifted) led to a second pandemic wave. Despite an already falling reproduction number in the autumn to winter of 2020, a second lockdown was instigated leading 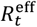 to fall below one once again. Emergence of the more transmissible alpha variant, along with increased mixing during Christmas festivities, led to a substantial rise in cases towards the end of 2020, ahead of a third lockdown. The vaccination program, appears to have helped to curb epidemic spread in early 2021, while emergence of the delta variant, along with easing restrictions, has led to 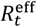 again increasing in the summer of 2021. Detailed analysis of the impact of these events is likely to prove difficult due to the cofounding effects of multiple factors.

**Figure 4:**
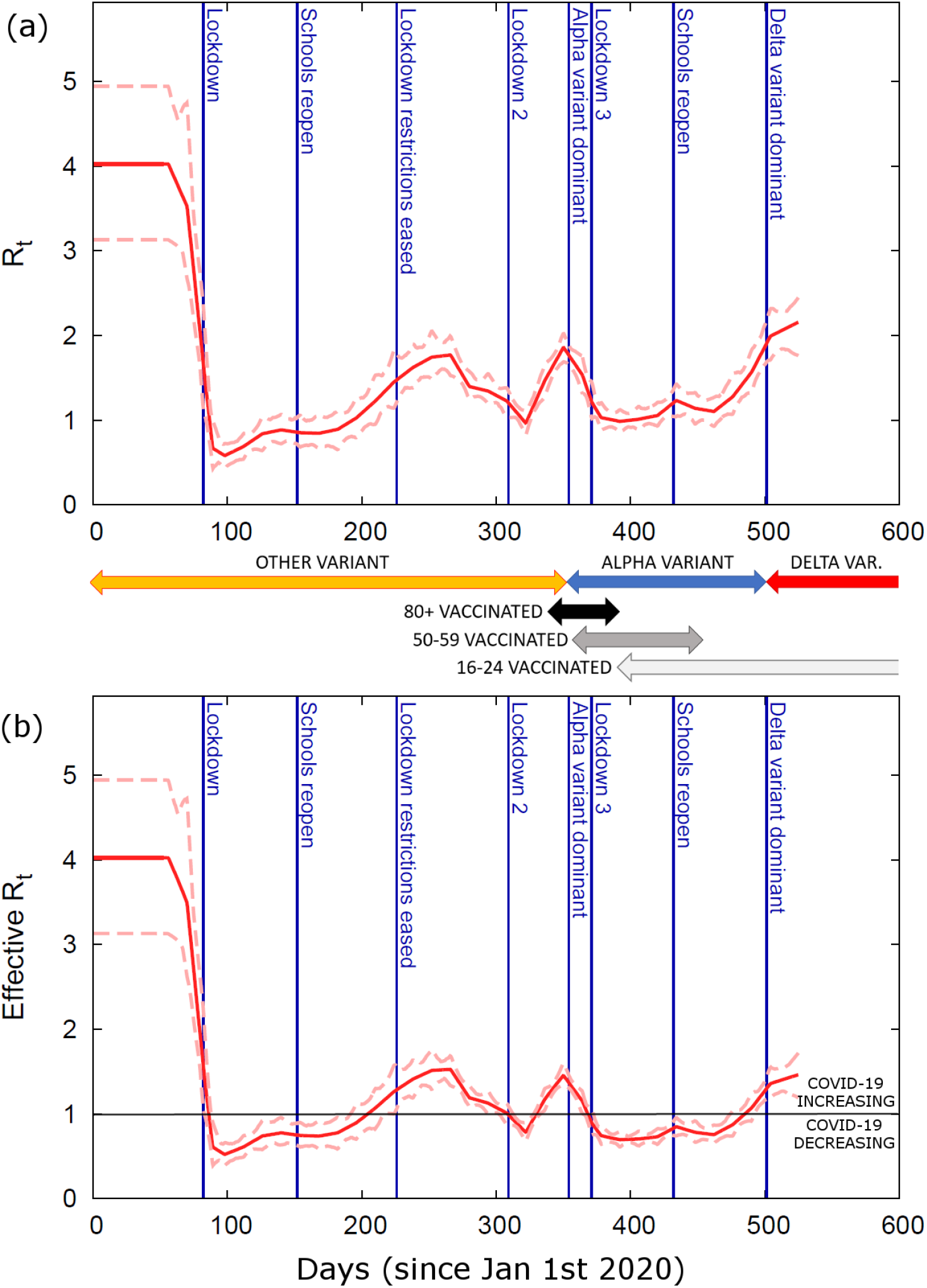
Time variation in reproduction number. (a) The inferred value of the reproduction number *R*_*t*_ as a function of time (red line gives posterior mean and dashed lines indicate the 95% credible interval). This is proportional to the overall effective contact rate (see Eq.(3)). (b) The effective reproduction number 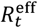 (which accounts for the fact that a fraction of the population is not susceptible). Above/below the horizontal black lines shows where COVID-19 is increasing/decreasing. The vertical blue lines denote important milestones during the epidemic [29]. Arrows showing changes in the dominant COVID-19 variant are estimated from COG UK [35] (see Appendix O) and for vaccination come from CIS [24] (these indicate the time period in which between 5% and 95% of individuals are vaccinated with the first dose for selected age groups).

#### Age-dependence in branching probabilities

Figure 5(a) shows the posterior estimated asymptomatic branching probability 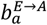 for different age groups *a*. Overall, around 55% of exposed individuals are asymptomatic. This value is consistent with the large range of figures suggested in the literature (*e*.*g*. from 18%-75% [29-31]). Asymptomatic branching probabilities are highest for younger age groups, which has important implications, *e*.*g*. for interpreting school cases rates (which may significantly under-represent true infection levels). The proportion of asymptomatics declines through adulthood and rises again in older age groups (Fig. 5(a)) consistent with other studies based on data prior to vaccine availability (see *e*.*g*. Fig. 2(c) in [22]). The notable exception is in the 80+ and care home groups (which, by definition, are set equal^22^), which have a significantly lower asymptomatic branching probability. This result stems from the fact that although many cases are observed in the 80+ group, measurements taken by the CIS near the end of 2020 show only a small proportion of randomly sampled 80+ individuals are found to be antibody positive. One possibility is that waning immunity is much faster in this group, or that a weaker immune response results in more false negative antibody test results.

**Figure 5:**
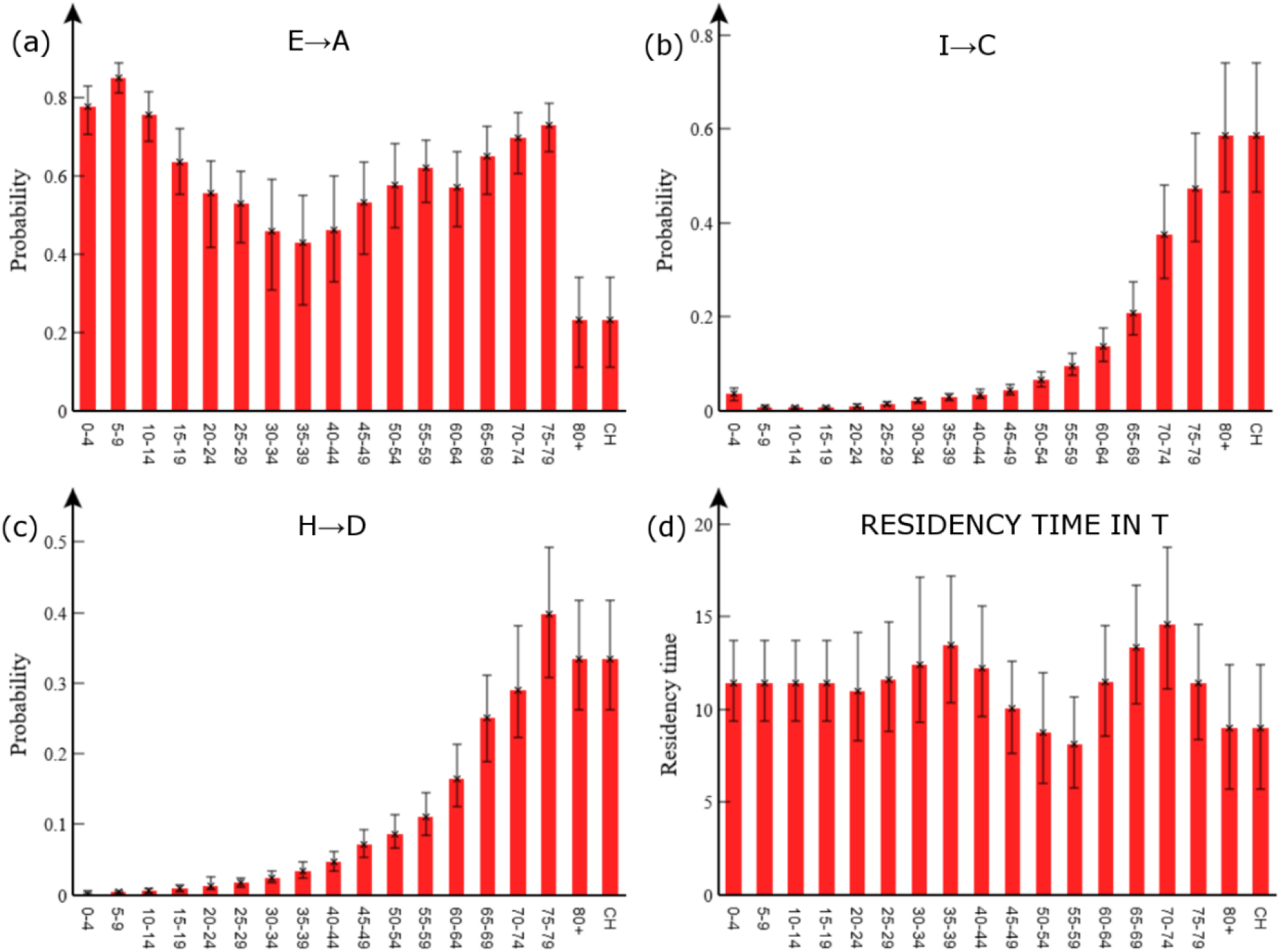
Age-dependent branching probabilities and residency time in T. The probability of: (a) becoming asymptomatic, (b) becoming hospitalised given a case (note, for care home patients “hospitalised” may mean going to hospital or becoming critically ill within a care home), and (c) of death given hospitalised. (d) Shows the residency time in the PCR test-sensitive T compartment. The error bars indicate 95% credible intervals.

**Figure 6:**
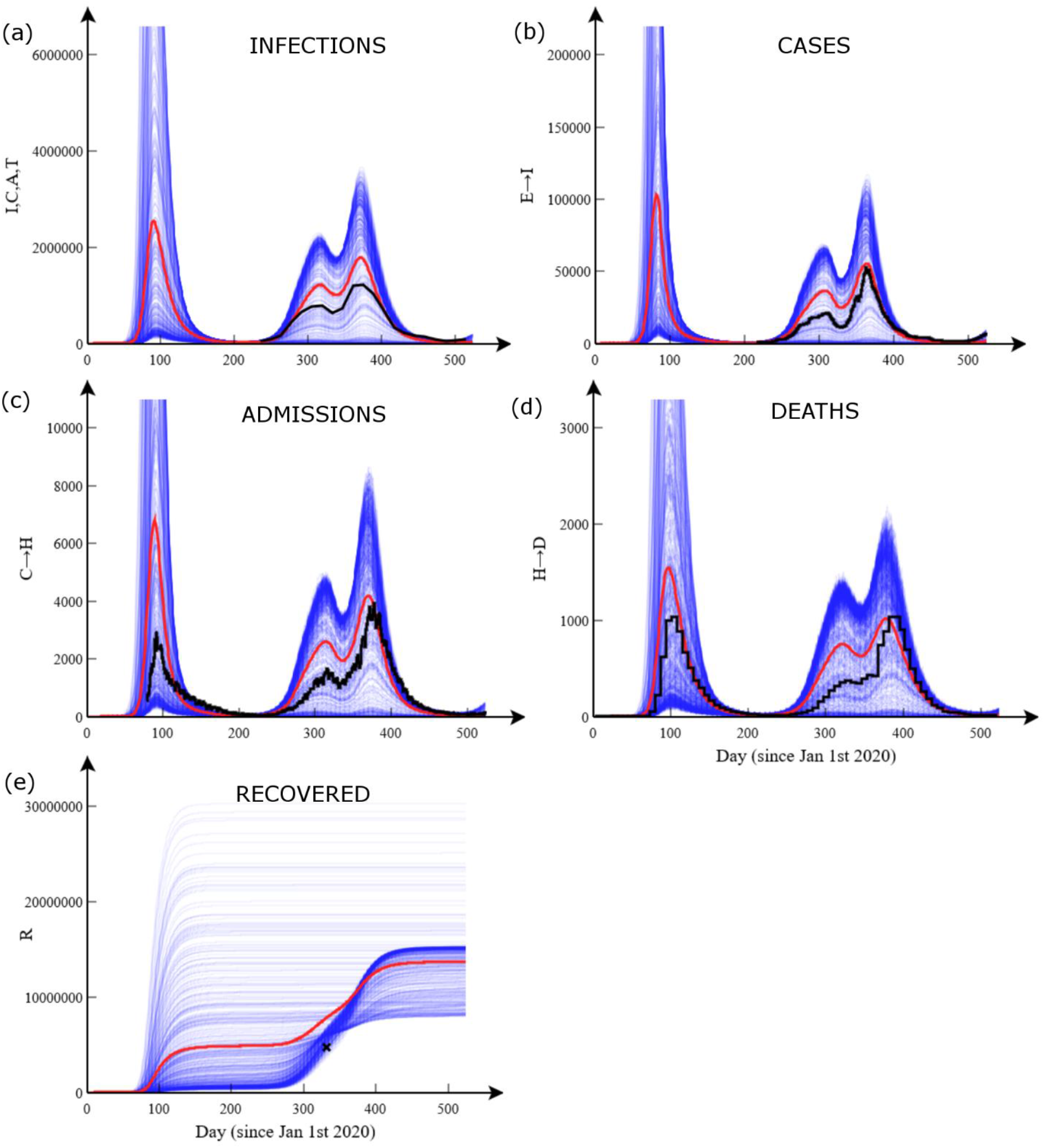
Simulation results and data. These plots show age-aggregated data (black) against 500 simulations (blue) performed using model parameters taken from the analysis posterior means (see Table R1 of the posterior Supplementary Results). The red lines indicate an average across simulations. (a) The total population in the infected I, C, A and T compartments (data from CIS), (b) daily cases, and (c) daily hospital admissions (not including care home patients), (d) daily deaths, (e) total recovered (excludes 0-14 age groups and care home residents, single data point from antibody results in CIS). Note, to aid visualisation of the data the first peak has been cut off.

The probability of hospitalisation (given a case) 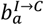 and dying in hospital (given hospitalised) 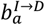 are shown in Figs. 5(b) and 5(c), respectively. These detailed estimates are entirely consistent with the broad understanding that COVID-19 affects the elderly much more severely than the young.

#### Duration of PCR test sensitivity

Estimated residency times in the PCR test-sensitive T compartment are shown in Fig. 5(d). As both I and A compartments are also detectable by PCR test, adding a mean value from this plot of 11 days to *m*_*E*_=*m*_*A*_=4 days leads to an estimated overall test-sensitive period of around 15 days. This is in good agreement with [32], who estimated the time from symptoms to two negative PCR tests at 13 days, which when added to an estimated 2 days presymptomatic period [32] gives exactly our figure. It might be expected the test-sensitive period would increase with age (Fig. 5(d)), but importantly T represents non-hospitalised individuals with significantly milder symptoms than hospitalised cases.

## Discussion

ABC-MBP has been shown to be reliable and efficient at producing informative posterior distributions of many key parameters of a complex epidemiological COVID-19 model that integrates age-stratified transitions and contact rates. Here we discuss the impact of key assumptions on the robustness of the estimates obtained.

Compartmental residency times in the model are based on evidence in the literature (see Appendix N), but a variety of alternative estimates exist. To understand the impact of such uncertainty consider the three key interrelated quantities that determine epidemiological dynamics: the reproduction number, the generation time^23^, and the infection rate. As infection rate is fully determined by observed data, a shift in the generation time (brought about by making alterations to any of the assumed residency times) must be accompanied by a corresponding rescaling of the reproduction number around one to maintain the observed infection rate. Appendix N investigates this and concludes the results are robust to inaccuracies in fixed residency times in the compartmental model, excepting this rescaling in the profile for *R*_*t*_. Results are also insensitive (Appendix N) to rescaling of the estimated external force of infection (Appendix D). Further support for our inferences comes from detailed analysis of the raw age-stratified data, which exhibits similar estimates and trends with age obtained for the various quantities reported here (Appendix P).

Our results suggest contact patterns during the COVID-19 outbreak in England are significantly different to pre-pandemic patterns. Although plausible and consistent with other results (see *e*.*g*. [5]), alternative explanations may also be consistent with the available data. As discussed above, there is the possibility for age-dependent residency times and/or infectiousness, but support for these hypotheses in the literature seems limited for COVID-19 [21]. We therefore now consider age-dependent susceptibility *σ*_*a*_ (whilst fixing the contact matrix to **C**^**0**^), for which the force of infection from Eq.(1) becomes:

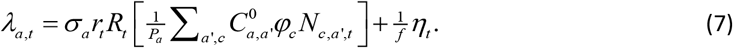

Estimates under this model are shown in Fig. 3(b), and, perhaps unsurprisingly, they exhibit a similar profile to the age contact factors in Fig. 3(a) (note, Appendix Q shows that other inferred parameter values undergo almost no change, which is further evidence of the robustness of these estimates).

So which model is correct, one which implies age-variation in susceptibility, or age-variation in expected contact rate, or perhaps both? One clue can be provided by [22], which suggests that susceptibility in individuals under 20 years of age is actually less than in older individuals (although this study is based on case data collected relatively early on in the pandemic, and is perhaps open to bias). This is contrary to what we find. Indeed, if contact patterns were to be estimated under this assumption, even larger increases in relative contact rates for younger age groups would be found, suggesting our estimates with Eq.(1) are actually conservative. Thus, we suggest observed increases in force of infection from younger age groups are more plausibly explained by changes in contact pattern. Nonetheless, there may be age-dependent effects in contact rate, susceptibility, infectivity and duration of disease states, but the public data considered here is not sufficient to estimate these simultaneously. Importantly, however, the model based on Eq.(1) is more accurate than one not containing such age contact or susceptibility factors, and so provides an important improvement for more accurate prediction, and highlights important avenues for further investigations regarding these confounding effects.

Figure 6 compares national-level age-aggregated data with simulations based on inferred parameters. We find relatively good agreement with simulation averages, but with large stochastic variability between runs. It is important to appreciate, however, that these results are contingent on historic lockdowns and restriction measures encoded in *R*_*t*_. In reality extremes of the distribution of modelled outcomes are unlikely to be realised as government interventions and human behaviour are dynamically linked to epidemic severity and would likely reduce 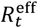 in the case of rapidly increasing case numbers.

Clearly the compartmental model in Fig. 1 does not capture all the features of the data, *e*.*g*. when examining posterior distributions for the state (in the posterior Supplementary Results) we see clear deviations between the model and the profiles for some of the data curves^24^. These discrepancies may be due to a number of factors not considered here, including observed spatial heterogeneity in the outbreak, time variation in the contact matrix (*e*.*g*. caused by opening and closing schools), the introduction of vaccination, and emergence of different COVID-19 variants. In principle these various effects could be studied using the methodology presented in this paper (no doubt requiring significantly more computation and in some cases additional data), and these remain the subject of future work.

## Conclusions

This paper has demonstrated the feasibility of fitting complex compartmental stochastic models containing a large number of parameters (over 150) to a large dataset (almost 9000 individual observations). This was achieved using a new ABC-MBP inference algorithm that is highly parallelisable and generic. Application to publicly available COVID-19 data demonstrated a good model fit based on the inferred parameter values, and yielded several new insights. Firstly, an indication that younger age groups come into contact more frequently than is suggested by a commonly used contact matrix taken from the pre-pandemic literature. Secondly, estimates for age variation in the probability individuals become asymptomatic revealed more asymptomatic young. Thirdly, estimation of the time-varying reproduction number which could inform intervention effectiveness and impacts of COVID-19 variants and vaccination. Finally, a consistently parameterised age-structured model is presented and available to support data-driven public health policy via simulation of near-term projections and of future scenarios.

## Supporting information

Supplementary Results

## Data Availability

All data produced in the present work are contained in the manuscript

## Additional Information

### Data Accessibility

The datasets supporting this article are available in GitHub [here](https://github.com/ScottishCovidResponse/BEEPmbp/tree/master/COVID%2019%20England).

Details on how to run the analyses reported are provided in the supplementary materials (Appendix M) and the code is available on GitHub here.

### Authors’ Contributions

CMP originated the idea for ABC-MBP and performed the cleaning of the raw data, the analysis, and drafted the manuscript. GM helped with epidemiological modelling, the inference algorithm and making improvements to the text. ADW helped with modelling and revising the text. All authors read and approved the manuscript.

## Acknowledgments

We would like to acknowledge Helen Brown, Stella Mazeri, and Margo Chase-Topping for many useful discussions regarding COVID-19 modelling and data acquisition. Also research software engineers Ian Hinder and Robin Williams for their contributions in developing BEEPmbp (the software used to perform the analysis). Also we express our gratitude to DiRAC (Distributed Research utilising Advanced Computing) for allowing us to use their HPC facilities for free during the pandemic.

## Footnotes

1 Such over-dispersion allows the particle number to be reduced substantially. This study used 96 particles, but many more would be required with a realistic observation model.

2 For brevity, we subsequently refer to these as “age groups”, even though one group actually relates specifically to care homes.

3 The I compartment includes both pre-symptomatic and symptomatic classifications of disease progression.

4 This matrix is asymmetric because populations in different age groups are not the same (notably the care home group has a significantly smaller population).

5 Such a constraint is necessary, otherwise rescaling ***v*** becomes confounded with rescaling *r*_*t*_ in Eq.(1).

6 Analysis of spatial data has shown this to be more or less true (results not show).

7 Disentangling these two effects is not possible from the data.

8 With age-structured populations set to be representative of England using ONS data.

9 For example, taking the difference between *Y*_*i*_ and *y*_*i*_ and squaring would result in something akin to a least-squares approach.

10 Its value is set to 2% of the highest peak from the time series *g*_*i*_ to which the measurement belongs. This was a simple way to suppress the effect of noise in measurements with low values.

11 This reflects the fact that daily transitions, like case and death data, are expected to be Poisson distributed, and so larger values are expected to be less susceptible to stochastic variation than smaller values.

12 Some time series have a daily time step whereas some are weekly. *w*_*i*_ helps to adjust for this discrepancy.

13 Sensitivity analysis is performed later to identify what affect changing these parameters has on the results.

14 *P* must be set to an even number.

15 In practice this is achieved by ordering the error function values for each particle, and setting the cut-off to the average of the middle two values on the list.

16 This is a property whereby a particle will explore the potential parameter and state space in a relatively small number of updates. So called “poor mixing” results from correlations in the posterior requiring MCMC to take many updates to generate a effectively new random sample from this space.

17 In stochastic models, simulation with a fixed parameter set can lead to a huge variation in the error function. On the other hand, MBPs ensure that a small change in parameter space is always accompanied by a small shift in the error function. Consequently MBPs can be effectively tuned to keep the system state error function within the ever decreasing cut-off limit.

18 Not all runs were found to converge on precisely the same solution, indicating some level of multimodality in the posterior (they were, however, all close, essentially exhibiting only small deviations in *R*_*0*_).

19 Generally speaking they fit well with some notable exceptions discussed below.

20 The CH column in Fig. 2(b) has a darker red shades than other columns.

21 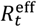 accounts for depletion in the susceptible population as a result of infection-induced immunity.

22 With information coming solely from the 80+ age group through the CIS data.

23 Defined as the interval between infection time in an individual and when that person transmits to another individual.

24 Note, these cannot simply be the result of stochastic fluctuations, as that is already accounted for.

