## Supplementary Materials for "Estimation of age-stratified contact rates during the COVID-19 pandemic using a novel inference algorithm"

### Appendix A: The reproduction number $R_t$

The force of infection in Eq.(1) is parameterised in terms of a time-varying quantity $R_t$ referred to as the "reproduction number". This appendix explains what this quantity is and how it is related to other model parameters.

#### Definitions

We begin with a clarification of terminology:

**Generation** – An epidemic starts with a single infected individual. We refer to this as "generation 1". That individual then goes on to infect other secondary infections which make up generation 2. Those individuals subsequently infect generation 3, and so on and so forth.

**Basic reproduction number $R_0$** – This is often defined to be the expected number of cases directly caused by the individual in generation 1 (*i.e.* in contact with an otherwise completely susceptible population). However, for models that include different demographic classifications, *e.g.* age and/or sex, or complex compartmental structures, careful consideration needs to be given to precisely how $R_0$ is calculated. In particular, just averaging over the demographic possibilities for the initially infected individual is not enough. One must also consider what happens in subsequent generations in the early phase of an epidemic to obtain a meaningful estimate for $R_0$ (it typically takes several generations for the distribution in demographic groups that cause the bulk of disease transmission to manifest). See below for how $R_0$ is actually calculated.

**Time-varying reproduction number $R_t$** – This is defined as the expected number of secondary cases directly generated by a case in the population in which nearly all individuals are susceptible. Consequently, $R_t$ should be interpreted as a quantity proportional to the "effective" rate at which individuals come into contact with each other (this combines the actual rate of contacts with the probability of disease transmission on contact). So, for example, when $R_t$ decreases it indicates that either individuals are meeting less frequently, or disease control measures are blocking transmission somehow (such as mask wearing). Note, disease transmission only actually happens when effective contacts occur between infected and susceptible individuals. It is important to remember that $R_t$ *does not* account for the reduction in the susceptible fraction of the population as the epidemic progresses.

**Time-varying effective reproduction number $R_t^{\text{eff}}$** – The effective reproduction number is the expected number of cases directly caused by an infected individual as a function of time $t$. Note, this *does* take into account the fact that as the epidemic progresses the fraction of susceptible individuals reduces (causing the transmission upon contact of individuals to become less and less frequent). $R_t^{\text{eff}}$ is always less than $R_t$, and when it reduces below one herd immunity is reached (*i.e.* the disease naturally dies out over time).

#### Relationship between reproduction number and transmission rate

Perhaps a more standard way to write the force of infection in Eq.(1) is in terms of a transmission rate parameter:

$$\lambda_{a,t} = \beta_t \sigma_a \tfrac{1}{P_a} \sum_{a',c} C_{a,a'} \varphi_c N_{c,a',t}, \tag{A1}$$

where here we ignore the external force of infection (as it doesn't play a role in the calculation of $R_t$) and $\sigma_a$ is an age-dependent relative susceptibility (note, this is not in Eq.(1) from the paper, but we include it here for completeness). The parameter $\beta_t$ in Eq.(A1) is called the "transmission rate", which is a proportionality constant that relates quantities measuring the rate of contacts between individuals in the population to the actual probability per unit time of an individual becoming infected (so here $\beta_t$ incorporates effects such as mask wearing, social distancing *etc*…). Comparing Eqs.(1) and (A1) we identify the relationship

$$\beta_t = r_t R_t. \tag{A2}$$

This equation simply states that the transmission rate is proportional to the reproduction number through a time-varying factor $r_t$. Below we show how $r_t$ can be calculated from $R_t$ and other model parameters.

#### Calculating $R_t$

We outline here the approach taken by Diekmann *et al.* [4] to calculate the reproduction number $R_t$. First, a set of compartments Ω for which individuals are infected (but not necessarily infectious) is identified. For the model in Fig. 1 this is given by Ω = $\{E_a, I_a, A_a\}$, where $a$ goes over different age groups[1]. For brevity the expressions shown below are for just two age groups $a_1$ and $a_2$, but these results can easily be extended to an arbitrary number (*e.g.* the 18 age groups used in the paper).

A vector[2] $\mathbf{p} = (E_{a_1}, E_{a_2}, I_{a_1}, I_{a_2}, A_{a_1}, A_{a_2})^T$ gives the number of individuals in each of the compartments in Ω. In the deterministic limit, the time evolution in $\mathbf{p}$ is given by

$$\frac{d\mathbf{p}_t}{dt} = (\mathbf{F}_t - \mathbf{\Sigma})\mathbf{p}_t, \tag{A3}$$

where $\mathbf{F}_t$ is a matrix accounting for the rate of individuals *entering* compartments in Ω, and $\mathbf{\Sigma}$ is a constant matrix accounting for transitions *between* and *leaving* compartments within Ω.

From Fig. 1 we see that individuals enter Ω through the exposed $E_{a_1}$ and $E_{a_2}$ compartments, caused by infectious individuals in the $I_{a_1}$, $I_{a_2}$, $A_{a_1}$ and $A_{a_2}$ compartments. Using the force of infection expression in Eq.(A1), we can derived:

---

[1] In principle other compartments such as C, T and H could also be included, but since they themselves are uninfectious (and don't lead to any infectious compartments), they can be treated as though they are uninfected states.

[2] The superscript "T" stands for "transpose", which converts a row vector into a column vector.

$$\mathbf{F}_t = \beta_t \begin{bmatrix} 0 & 0 & \sigma_{a_1}\phi_{a_1,t}C_{a_1,a_1}\varphi_{\mathrm{I}} & \sigma_{a_1}\phi_{a_1,t}C_{a_1,a_2}\varphi_{\mathrm{I}} & \sigma_{a_1}\phi_{a_1,t}C_{a_1,a_1}\varphi_{\mathrm{A}} & \sigma_{a_1}\phi_{a_1,t}C_{a_1,a_2}\varphi_{\mathrm{A}} \\ 0 & 0 & \sigma_{a_2}\phi_{a_2,t}C_{a_2,a_1}\varphi_{\mathrm{I}} & \sigma_{a_2}\phi_{a_2,t}C_{a_2,a_2}\varphi_{\mathrm{I}} & \sigma_{a_2}\phi_{a_2,t}C_{a_2,a_1}\varphi_{\mathrm{A}} & \sigma_{a_2}\phi_{a_2,t}C_{a_2,a_2}\varphi_{\mathrm{A}} \\ 0 & 0 & 0 & 0 & 0 & 0 \\ 0 & 0 & 0 & 0 & 0 & 0 \\ 0 & 0 & 0 & 0 & 0 & 0 \\ 0 & 0 & 0 & 0 & 0 & 0 \end{bmatrix}, \quad \text{(A4)}$$

where $\phi_{a,t}$ gives the time-varying fraction of the susceptible population in age group $a$, $\sigma_a$ gives the relative susceptibility, $\mathbf{C}$ is the age-adjusted contact matrix, and, finally, $\varphi_c$ is the relative infectivity of compartment $c$.

When calculating $R_t$ (as opposed to $R_t^{\mathrm{eff}}$) a key assumption is that the epidemic growth is occurring when the population is almost entirely susceptible. This allows us to replace $\phi_{a,t}$ in Eq.(A4) with $\phi_a$, the fraction of the population in age group $a$[3]. Consequently

$$\mathbf{F}_t = \beta_t \begin{bmatrix} 0 & 0 & \sigma_{a_1}\phi_{a_1}C_{a_1,a_1}\varphi_{\mathrm{I}} & \sigma_{a_1}\phi_{a_1}C_{a_1,a_2}\varphi_{\mathrm{I}} & \sigma_{a_1}\phi_{a_1}C_{a_1,a_1}\varphi_{\mathrm{A}} & \sigma_{a_1}\phi_{a_1}C_{a_1,a_2}\varphi_{\mathrm{A}} \\ 0 & 0 & \sigma_{a_2}\phi_{a_2}C_{a_2,a_1}\varphi_{\mathrm{I}} & \sigma_{a_2}\phi_{a_2}C_{a_2,a_2}\varphi_{\mathrm{I}} & \sigma_{a_2}\phi_{a_2}C_{a_2,a_1}\varphi_{\mathrm{A}} & \sigma_{a_2}\phi_{a_2}C_{a_2,a_2}\varphi_{\mathrm{A}} \\ 0 & 0 & 0 & 0 & 0 & 0 \\ 0 & 0 & 0 & 0 & 0 & 0 \\ 0 & 0 & 0 & 0 & 0 & 0 \\ 0 & 0 & 0 & 0 & 0 & 0 \end{bmatrix}. \quad \text{(A5)}$$

We now turn our attention to the matrix $\mathbf{\Sigma}$ in Eq.(A3), which represents transitions between and leaving the infected compartments in Ω. Based on the compartmental model in Fig. 1, this is given by:

$$\mathbf{\Sigma} = \begin{bmatrix} \frac{1}{m_E} & 0 & 0 & 0 & 0 & 0 \\ 0 & \frac{1}{m_E} & 0 & 0 & 0 & 0 \\ -\frac{1}{m_E}b_{a_1}^{\mathrm{E}\to\mathrm{I}} & 0 & \frac{1}{m_I} & 0 & 0 & 0 \\ 0 & -\frac{1}{m_E}b_{a_2}^{\mathrm{E}\to\mathrm{I}} & 0 & \frac{1}{m_I} & 0 & 0 \\ -\frac{1}{m_E}b_{a_1}^{\mathrm{E}\to\mathrm{A}} & 0 & 0 & 0 & \frac{1}{m_A} & 0 \\ 0 & -\frac{1}{m_E}b_{a_2}^{\mathrm{E}\to\mathrm{A}} & 0 & 0 & 0 & \frac{1}{m_A} \end{bmatrix}, \quad \text{(A6)}$$

where $m_c$ is the mean residency time in compartment $c$ and $b_a^{\mathrm{tr}}$ are branching probabilities leaving the exposed compartment along either the infectious and asymptomatic branches. This matrix was constructed by considering each transition in the model in turn. Denoting the initial and final infected compartments to be $i$ and $j$, matrix element $\Sigma_{ii}$ receives a positive contribution given by the individual rate (because individuals are leaving compartment $i$) and $\Sigma_{ji}$ gets a corresponding negative contribution (because those individuals are entering compartment $j$). Note, if $j$ is not one of the infected compartments in Ω, this second contribution is ignored.

The inverse of the matrix in Eq.(A6) is given by

[3] Note, $\phi_a$ contains a fixed set of quantities available from census data.

$$\boldsymbol{\Sigma}^{-1} = \begin{bmatrix} m_E & 0 & 0 & 0 & 0 & 0 \\ 0 & m_E & 0 & 0 & 0 & 0 \\ m_I b_{a_1}^{\mathrm{E}\to\mathrm{I}} & 0 & m_I & 0 & 0 & 0 \\ 0 & m_I b_{a_2}^{\mathrm{E}\to\mathrm{I}} & 0 & m_I & 0 & 0 \\ m_A b_{a_1}^{\mathrm{E}\to\mathrm{A}} & 0 & 0 & 0 & m_A & 0 \\ 0 & m_A b_{a_2}^{\mathrm{E}\to\mathrm{A}} & 0 & 0 & 0 & m_A \end{bmatrix}. \quad \text{(A7)}$$

This has a simple interpretation: If an individual enters compartment $i$, $\Sigma_{ji}^{-1}$ gives the time, on average, that individual is expected to spend in compartment $j$ (eventually). For example, inspecting the first column in Eq.(A7) we see that if an individual enters compartment $\mathrm{E}_{a_1}$ (at the top) it will stay in $\mathrm{E}_{a_1}$ for time $m_E$, spend no time in $\mathrm{E}_{a_2}$ (individuals do not change age group), time $m_I b_{a_1}^{E\to\mathrm{I}}$ in compartment $\mathrm{I}_{a_1}$ (this average is made up of $m_I$ if the individual branches to the I compartment with probability $b_{a_1}^{E\to\mathrm{I}}$ and zero if it doesn't) and time $m_A b_{a_1}^{E\to\mathrm{A}}$ in compartment $\mathrm{A}_{a_1}$.

We now construct the next generation matrix (NGM) $\mathbf{K}_t$. We denote $\mathbf{w}_g$ to be a vector giving the number of individuals entering the different compartments in Ω in generation $g$. The vector for the next generation is given by

$$\mathbf{w}_{g+1} = \mathbf{K}_t \mathbf{w}_g \quad \text{where} \quad \mathbf{K}_t = \mathbf{F}_t \boldsymbol{\Sigma}^{-1}. \quad \text{(A8)}$$

The reasoning behind this expression is as follows. Suppose we consider an individual in generation $g$ that enters compartment $i$. As described above, during its infected lifetime this individual spends on average $\Sigma_{ji}^{-1}$ in each compartment $j$. But in doing so Eq.(A5) tells us that through its own infectiousness it will generate $F_{kj}$ new infected individuals in each of the compartments $k$. This means that $w_{g+1,k}$ gains a contribution $F_{kj}\Sigma_{ji}^{-1}$ for each of the $w_{g,i}$ individuals in generation $g$ that enter compartment $i$. Summing over all the possible values for $i$ and $j$ gives the relationship in Eq.(A8).

Next, we ask the question what happens when we iterate Eq.(A8) over many generations? It turns out that this equation can easily be solved[4], as shown in Appendix B. In summary, as the generations increase, so $\mathbf{w}_g$ becomes proportional to the eigenvector $\mathbf{e}$ with the largest eigenvalue of the NGM $\mathbf{K}_t$. This eigenvalue provides an estimate for $R_t$ (because, by definition, this is the average number of infections an individual in one generation causes in the next).

#### Calculating $r_t$

The explanation above showed that given a transmission rate $\beta_t$ and other model parameters an estimate for $R_t$ can be made. The way the model is parameterised in the paper, however, this process works the opposite way around. Model parameters are used to inform a spline for $R_t$, and this in turn is used to calculate $r_t$, which then goes on to inform $\beta_t$ through Eq.(A2). We now provide a description of how this is done.

First, we define the quantity

$$\mathbf{F}_t' = \frac{\mathbf{F}_t}{\beta_t}. \quad \text{(A9)}$$

[4] We assume the timescale of the initial phase of the epidemic is slow compared to time variation in other model parameters, *e.g.* $\beta_t$.

From Eq.(A5) we see that $\mathbf{F}_t'$ can be calculated using known model parameters. Next, we define a modified NGM:

$$\mathbf{K}_t' = \mathbf{F}_t' \boldsymbol{\Sigma}^{-1}. \tag{A10}$$

The largest eigenvalue of this matrix gives $R_t$ divided by $\beta_t$. From Eq.(A2) we see that taking the reciprocal of this quantity gives $r_t$.

##### Calculating $\boldsymbol{R_t^{\text{eff}}}$

As mentioned above, the effective reproduction number is the expected number of cases directly caused by an infected individual as a function of time $t$. This has to account for the fact that some susceptible individuals have already been infected, recovered and acquired immunity.

Here, we use Eq.(A4), instead of Eq.(A5), when calculating the modified NGM:

$$\mathbf{K}'_{\text{eff},t} = \begin{bmatrix} 0 & 0 & \sigma_{a_1}\phi_{a_1,t}C_{a_1,a_1}\varphi_{\text{I}} & \sigma_{a_1}\phi_{a_1,t}C_{a_1,a_2}\varphi_{\text{I}} & \sigma_{a_1}\phi_{a_1,t}C_{a_1,a_1}\varphi_{\text{A}} & \sigma_{a_1}\phi_{a_1,t}C_{a_1,a_2}\varphi_{\text{A}} \\ 0 & 0 & \sigma_{a_2}\phi_{a_2,t}C_{a_2,a_1}\varphi_{\text{I}} & \sigma_{a_2}\phi_{a_2,t}C_{a_2,a_2}\varphi_{\text{I}} & \sigma_{a_2}\phi_{a_2,t}C_{a_2,a_1}\varphi_{\text{A}} & \sigma_{a_2}\phi_{a_2,t}C_{a_2,a_2}\varphi_{\text{A}} \\ 0 & 0 & 0 & 0 & 0 & 0 \\ 0 & 0 & 0 & 0 & 0 & 0 \\ 0 & 0 & 0 & 0 & 0 & 0 \\ 0 & 0 & 0 & 0 & 0 & 0 \end{bmatrix} \boldsymbol{\Sigma}^{-1}, \tag{A11}$$

If we define the largest eigenvalue of $\mathbf{K}_t'$ in Eq.(A10) to be $\lambda_t$, and for $\mathbf{K}'_{\text{eff},t}$ in Eq.(A11) to be $\lambda_{\text{eff},t}$, the effective reproduction number can be related to $R_t$ through

$$R_t^{\text{eff}} = \frac{\lambda_{\text{eff},t}}{\lambda_t} R_t. \tag{A12}$$

#### Appendix B: Solving iterative matrix equations

This appendix outlines the solution to the matrix equation in Eq.(A8). Note, we drop the index $t$ because it is assumed the timescale over which disease spreads (*i.e.* the generation time) is fast compared to the timescale over which model parameter changes (such as the transmission rate $\beta_t$).

We refer to $\mathbf{e}$ as an "eigenvector" and $\lambda$ as an "eigenvalue" of a matrix $\mathbf{K}$ if it solves the equation

$$\mathbf{Ke} = \lambda \mathbf{e}. \tag{B1}$$

In general, a square matrix of size $N$ will have $N$ eigenvectors $\mathbf{e}_j$ and eigenvalues $\lambda_j$, ordered such that $\lambda_1$ is the highest and $\lambda_N$ is the lowest. These can collectively be written as

$$\mathbf{KU} = \mathbf{U} \begin{bmatrix} \lambda_1 & 0 & 0 & \cdots \\ 0 & \lambda_2 & 0 & \cdots \\ 0 & 0 & \lambda_3 & \cdots \\ \vdots & \vdots & \vdots & \ddots \end{bmatrix}, \tag{B2}$$

where $\mathbf{U}$=[$\mathbf{e}_1$, $\mathbf{e}_2$, $\mathbf{e}_3$,…] is a matrix made up of the eigenvectors. Multiplying both sides of Eq.(B2) by the inverse matrix $\mathbf{U}^{-1}$ and substituting this expression for $\mathbf{K}$ into Eq.(A8) gives

$$\mathbf{w}_g = \mathbf{K}^{g-1}\mathbf{w}_1$$
$$= \left( \mathbf{U} \begin{bmatrix} \lambda_1 & 0 & 0 & \cdots \\ 0 & \lambda_2 & 0 & \cdots \\ 0 & 0 & \lambda_3 & \cdots \\ \vdots & \vdots & \vdots & \ddots \end{bmatrix} \mathbf{U}^{-1} \right)^{g-1} \mathbf{w}_1 \qquad \text{(B3)}$$
$$= \mathbf{U} \begin{bmatrix} \lambda_1^{g-1} & 0 & 0 & \cdots \\ 0 & \lambda_2^{g-1} & 0 & \cdots \\ 0 & 0 & \lambda_3^{g-1} & \cdots \\ \vdots & \vdots & \vdots & \ddots \end{bmatrix} \mathbf{U}^{-1}\mathbf{w}_1.$$

A key feature of this relationship is that as the generation number $g$ increases, so the diagonal element that corresponds to the largest eigenvalue $\lambda_1$ dominates over all others (which also means that $\mathbf{w}_g$ become proportional to the eigenvector $\mathbf{e}_1$). Because the overall number of individuals in $\mathbf{w}_g$ goes up by a proportion $\lambda_1$ each generation, so $\lambda_1$ provides an approximation to $R_t$.

#### Appendix C: Pre-pandemic contact matrix $\mathbf{C}^0$

This appendix details how the pre-pandemic contact matrix $\mathbf{C}^0$ in Fig. 2(a) is specified.

For contacts between 5 year age bands less than 70 year of age, elements of $\mathbf{C}^0$ are directly copied from data published by the BBC Pandemic study [5]. This study actually provides separate matrices for interactions at home, school, work and other, but for simplicity these separate contributions are added together. Unfortunately, the BBC Pandemic study does not provide information on interactions between individuals less than 10 years old (*i.e.* the 2x2 matrix in the top left-hand corner of $\mathbf{C}^0$ is missing). This part of the matrix has been filled in using data from Prem *et al.*[5] [6] (which is, itself, based on the POLYMOD study [7]).

Next, it is necessary to fill in elements of $\mathbf{C}^0$ for the older age groups and for care home residents. In particular, values along the diagonal and in the upper triangular region of $\mathbf{C}^0$ need to be specified (other values can be derived, as shown below).

First we consider contact rates for individuals in the 70-74 age group with the same or younger age groups. For simplicity, we assume these follow the same pattern as for the 65-69 age group, except all the ages are shifted by 5 years (*i.e.*, we set $C^0_{70-74,70-74} = C^0_{65-69,65-69}$, $C^0_{65-69,70-74} = C^0_{60-64,65-69}$, $C^0_{60-64,70-74} = C^0_{55-59,65-69}$, *etc*…). The unspecified new element $C^0_{0-4,70-74,}$ giving contacts with individuals in the 0-4 age group is set to be the same as $C^0_{5-9,70-74}$. This process is repeated a further two times to generate elements for columns relating to the 75-79 and 80+ age categories.

For the care home category CH, the contact rates are copied directly from the 80+ age group results, but with two modifications: 1) contacts between care home residents are boosted by a factor of

[5] Specifically, the contact rates in the 2x2 matrix for the 0-4 and 5-9 age groups were copied from the contact matrices published by Prem *et al.* [3], but rescaled such that contacts between individuals in the 10-14 age groups matched between the two studies.

three[6], and 2) contacts between care home residents and individuals in age categories ranging from 25 to 64 are also boosted by a factor of three[7].

It must be noted that there is a considerable level of arbitrariness in these choices in constructing the new elements of $\mathbf{C^0}$ for the older age categories and care home residents. Importantly, however, much of this subjectivity is corrected for during inference, because the rows and columns of $\mathbf{C^0}$ are multiplied by age contact factors $\boldsymbol{v}$ estimated directly from the data.

#### Contact symmetry

The matrix in Fig. 2(a) is asymmetric because the population sizes in the different age groups are not the same (the care home population, in particular, is substantially smaller than others, and so the asymmetry is most notable when comparing the care home row and column). However, because contacts must happen symmetrically between individuals, the total number between any two age classes $a$ and $a'$ must be the same, *i.e.*

$$C^0_{a,a'}P_{a'} = C^0_{a',a}P_a, \tag{C1}$$

where $P_a$ is the population in age group $a$. This can be rewritten as

$$C^0_{a,a'} = C^0_{a',a}\frac{P_a}{P_{a'}}. \tag{C2}$$

This means that if the upper triangular region of the matrix $\mathbf{C^0}$ is known, so the lower triangular region can be derived. Furthermore, other elements of the contact matrix $\mathbf{C^0}$ are corrected to ensure that Eq.(C2) is strictly satisfied:

$$C^{0\,\text{corrected}}_{a,a'} = \frac{1}{2}\left(C^0_{a,a'} + C^0_{a',a}\frac{P_a}{P_{a'}}\right). \tag{C3}$$

This corrected version is used as the contact matrix $\mathbf{C^0}$ in our analysis.

---

[6] Because they are typically in much closer contact than 80+ individuals in the community (who largely live in separate private housing).

[7] Care home workers are most likely to lie in this age range and can be expected to have frequent contacts with the care home residents.

#### Appendix D: External force of infection

The movement of infected individuals between countries causes COVID-19 to spread globally. The combined effect of sources of infection coming from outside England is incorporate into the model through the external force of infection $\eta_t$ in Eq.(1). Figure D1 shows an estimated time-variation in this quantity (used for the analysis in this paper). Below we outline how this curve is derived.

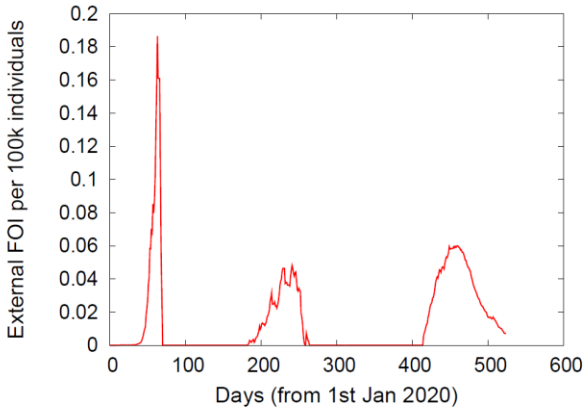


**Figure D1: External force of infection** – The estimated external force of infection (per 100,000 individuals) for England.

We take $f_d(t)$ to be the daily number of passengers moving to and from a destination country $d$ at time $t$. For simplicity, it is assumed the majority of these movements are from air travel. We use data provided by the Civil Aviation Authority [8], which publishes monthly statistics giving the number of passengers travelling to and from different UK airports to countries around the world (aggregated results are shown in Fig. D2(a)). These are converted to daily estimates of travel $f_d(t)$ by fitting a piecewise linear spline, as shown in Fig. D2(b).

When flying internationally, we assume travellers spend on average $\tau$ days in the destination country before returning home (we use the estimate $\tau$=4.6 days taken from [9]). This means that in country $d$ there is a steady-state population of $f_d(t)\times\tau/2$ residents who have travelled from England and *vice-versa*.

Next, we make use of estimated levels of COVID-19 infection in different countries as a function of time. Johns Hopkins University provides time series estimates for the number of daily COVID-19 cases and deaths for most countries in the world [10] (see Fig. D3(a) for results from countries with close links to the UK). Because of significant variation in testing availability across different countries, it was deemed that death data provides a more robust estimate for overall infection levels than case data. We define $n_d$(t) to be the time-varying death rate in country $d$. We estimate the daily infection rate by

$$\frac{n_d(t+\Delta)}{\mathrm{IFR}}, \tag{D1}$$

where Δ is the time difference between when a death is recorded and the time at which that individual was most likely to have transmitted their virus to a resident of England (an estimate of Δ=18 days is obtained by considering the compartmental model in Fig. 1[8]). In Eq.(D1) IFR stands for the "infection fatality rate", *i.e.* the probability of death given infection, which is taken to be 1.2% [11].

We assume that infections occur randomly across a country. Since Eq.(D1) gives the overall rate of infections, the rate in the sub-population of English travellers $f_d(t)\times\tau/2$ temporarily staying in the destination country $d$ is given by

[8] This comprises of $m_H+m_C+½\, m_I$, rounded to the nearest integer.

$$\frac{f_d(t)\tau}{2p_d}\frac{n_d(t+\Delta)}{\text{IFR}}, \tag{D2}$$

where $p_d$ is the population in the destination country [12].

However, we also have to take into account the fact that had those individuals stayed in England, they would have also been infected at a certain rate (note, these infection have already been accounted for in the model and so must be subtracted from our external force of infection estimate). In fact, it is the difference in prevalence between England and the destination country that yields the correct contribution to the external force of infection[9]:

$$\frac{f_d(t)\tau}{2\times\text{IFR}}\left(\frac{n_d(t+\Delta)}{p_d}-\frac{n_{\text{England}}(t+\Delta)}{p_{\text{England}}}\right). \tag{D3}$$

The calculation above focused on English residents who travelled to a destination country and acquired infection in that country. However, there is a further external force of infection coming from residents of the destination country travelling to England and passing on their infection whilst staying in England. It turns out that this contribution is again given by Eq.(D3), and so summing up the two contributions removes the factor 2 in the denominator.

Summing over all destination countries finally provides an estimate for the overall external force of infection:

$$\eta(t)=\max\left\{\frac{\tau}{\text{IFR}}\sum\nolimits_d f_d(t)\left(\frac{n_d(t+\Delta)}{p_d}-\frac{n_{\text{England}}(t+\Delta)}{p_{\text{England}}}\right),0\right\}, \tag{D4}$$

where the max function ensures that this quantity is strictly positive (in cases in which England has a higher prevalence of COVID-19 than countries with which it interacts, it can be considered as the source of external infections for those other countries, and *vice-versa*).

Based on integrating the curve in Fig. D1, we estimate that around 3500 externally generated infections occurred from the beginning of the pandemic up until June 9th 2021. Whilst this is small compared to the total number of infections, the timing and profile of the external force of infection curve is crucial in determining the initiation and severity of the COVID-19 pandemic waves.

It is important to mention that the curve in Fig. D1 probably represents something closer to a lower bound, and the actual force of infection may be somewhat higher. This is because of a number of simplifying assumptions used above: Firstly, other forms of transportation crossing borders were ignored, *e.g.* ferries, the Channel Tunnel and land borders with Scotland and Wales, which would have all provided contributions (unfortunately finding data for these aspects proved challenging). Secondly, we discounted the extra levels of transmission that occurs during transit (it can be imagined that enclosed spaces, like aircraft and terminal buildings, would likely be a significant source of elevated disease transmission). Lastly, it was assumed that travellers behave much like the local population to which they are travelling. It could be, however, that their contact rate with others is significantly higher compared to local residents, by virtue of the fact they are travelling.

---

[9] Note, $n_{\text{England}}$ is the empirically observed death rate (not the inferred death rate from the model) such that $\eta(t)$ is an external input into the model.

To test whether breaking these assumptions has a big impact on results, Appendix N looks at re-running the analysis with double the size of external force of infection. The results remained largely unchanged, which shows that provided the time-varying profile for the external force of infection curve is reasonably correct, results are robust to its magnitude.

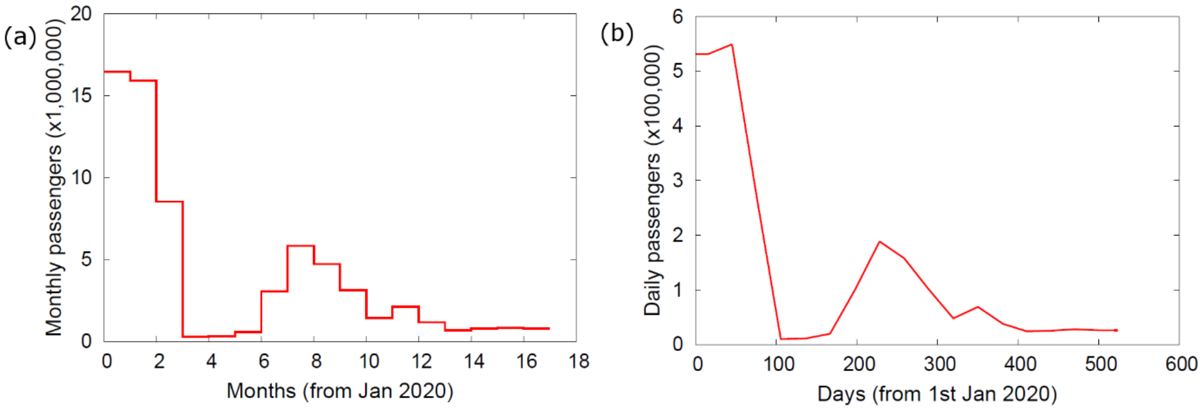


**Figure D2: Flight data**. (a) This shows raw data provided by the Civil Aviation Authority giving the number of monthly passengers travelling to and from England. (b) This is converted to estimated daily passenger numbers by fitting piecewise linear splines (for each destination country). Note, these graphs show aggregated results, but data is available separately for all destination countries.

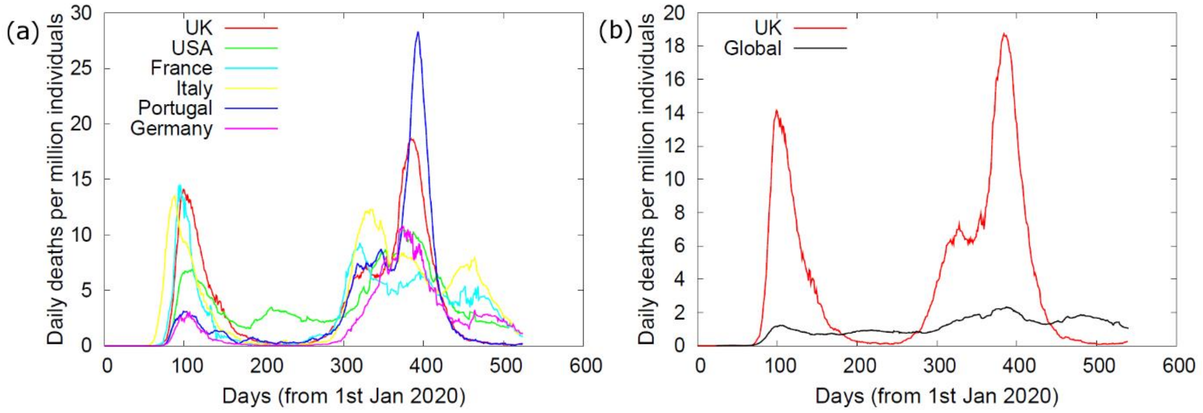


**Figure D3: Death rates**. This shows daily COVID-19 deaths per million individuals, as published by Johns Hopkins University [10]. (a) Results for a selection of countries with close links to the UK (in fact, results from all 141 countries were used in the analysis). (b) The red line shows UK results and the black line shows global results (other than the UK), weighted by air traffic to the destination countries.

### Appendix E: System dynamics

This appendix describes the $\tau$-leaping algorithm used to model system dynamics [13]. This is a discrete-time algorithm with time step $\tau$.

The boxes in Fig. 1 show the different compartments in the model. We denote $p_{a,c,t}$ to be the population in age group $a$, compartment $c$, at time $t$.

Transitions within the model, represented by the arrows in Fig. 1, are indexed by $j$. They move individuals from an initial compartment $i_j$ to a final compartment $f_j$. Within a time period between $t$ and $t+\tau$ the number of transitions of type $j$ in age group $a$ is taken to be $n_{a,j,t}$. Because the residency times in the compartments are taken to be exponentially distributed, so $n_{a,j,t}$ is Poisson distributed.

Simulation from the model is implemented in the following way:

1) **Initialisation** – The time $t$ is set to $t_{\text{start}}$ and the initial population sizes are set $p_{a,c,t_{\text{start}}}$. By default all individuals are assumed to start in the susceptible compartment S. The population sizes for each age group are taken from census data described in the main paper.
2) **Sample transition numbers** – We go through each age group $a$ and transition type $j$ and sample the transition number. For the infection transition I→E:

$$n_{a,\mathrm{I}\to\mathrm{E},t} \sim \mathrm{Poisson}\left(\tau p_{a,S,t}\lambda_{a,t}\right), \tag{E1}$$

where the force of infection $\lambda_{a,t}$ is obtained from Eq.(1).

For non-infection transitions $j$:

$$n_{a,j,t} \sim \mathrm{Poisson}\left(\tau p_{a,i_j,t} b_a^j / m_{i_j,a}\right), \tag{E2}$$

where $b_a^j$ is the branching probability for individuals to move down transition $j$ (as opposed to any other transition leaving the initial compartment $i_j$) for age group $a$, and $m_{i_j,a}$ is the residency time in compartment $i_j$ (*i.e.*, the compartment from which the individuals are leaving).

3) **Update populations** – As a result of the transitions in Eqs.(E1) and (E2), compartmental populations are updated according to:

$$p_{a,c,t+\tau} = p_{a,c,t} + \left[\sum_{j\in\text{enter c}} n_{a,j,t}\right] - \left[\sum_{j\in\text{leave c}} n_{a,j,t}\right], \tag{E3}$$

where the first sum goes over all transitions that enter compartment $c$ (increasing its population), and the second goes over all those that leave $c$ (decreasing its population).

4) **Iterate** – Increment $t$ by $\tau$ and if less than $t_{\text{end}}$ go to step 2.

Note, the $\tau$-leaping algorithm uses a finite time step τ. Making τ closer to zero results in the algorithm dynamics aligning more and more closely to the exact, continuous time Doob-Gillespie algorithm [14]. This, however, comes at a significant computation cost. For the analysis in this paper $\tau$=0.5 days was chosen because it seemed a sensible choice balancing accuracy with computational speed (importantly, it is significantly shorter than the residency times in any of the compartments).

### Appendix F: Data Sources

This appendix provides information on publicly available data sources used in the analysis (along with details on how some of these have been transformed to allow them to be incorporated).

#### i) Demographic data

**Population data** – Population sizes $P_a$ for each age category in England are taken from 2020 estimates provided by the Office for National Statistics (ONS)[10]. These define the initial susceptible population

[10] These figures apply to the entire of the UK but are corrected to represent England by multiplying by the ratio of the estimated total English and UK populations.

for the system. The care home population in England is around 419,000 [15]. Following [16], we assume the age-structure in care homes is given by 65-69: 5%, 70-74: 5%, 75-79: 15%, 80+: 75%. Numbers in these categories are reduced accordingly in the general population.

#### ii) Operational data

**Death data** – The Office for National Statistics (ONS) publishes age-stratified death data [2] giving the weekly number of deaths in England and Wales[11]. The values for England alone are estimated by multiplying by a factor to match the overall deaths in England[12]. The age bands "<1" and "1-4" are combined to give a "0-4" category, and the bands "80-85", "85-89"and "90+" are combined to give a "80+" category. All other 5 yearly intervals are taken directly from the data. Care home deaths are removed from the general population and placed into the separate "CH" category (see below). During inference the death data provides information about weekly transitions from the $\mathrm{H}$ to $\mathrm{D}$ compartments for each age-class[13].

**Case data** – We define a "case" as an individual that has had at least one positive PCR test result. Case data, stratified into 5 year bands, was downloaded from the Gov.uk website [1]. A one week rolling average is used to correct for any week effect. The estimated number of care home cases is subtracted from the general population (see below). Case data is used only for the second wave of COVID-19 infection (from 8th August 2020), when the UK testing capability became less restricted (for a justification of this see Appendix H).

During inference case data were shifted back 4 days to inform the number of daily transitions from $\mathrm{E}$ to $\mathrm{I}$; this shift comes from adding a pre-symptomatic period of 2 days [17] to an estimated 2 days for symptoms to become sufficiently severe to induce a person to get tested. That cases inform the $\mathrm{E}$ to $\mathrm{I}$ transition sets an operational interpretation of the "asymptomatic" compartment as "non-case" for the second wave. Whilst clearly not synonymous with a clinical definition, those not tested in the second wave were, on average, much more likely to have had milder symptoms than those who did.

**Hospital admissions** – Two sources of information are used: the overall number of admissions since near the start of the pandemic (provided by the Gov.uk dashboard [18]) and age-stratified daily hospital admissions from 12th October 2020 (provided by Public Health England [19]). The latter provides data in the following age categories: "Age 0-5", "Age 6-17", "Age 18-54", "Age 55-64", "Age 65-74", "Age 75-84" and "Age 85+". These are transformed into the 5 year age bands using the method described in Appendix G. This data informs the daily number of transitions from the $\mathrm{C}$ to $\mathrm{H}$ compartments.

---

[11] This is by date of occurrence in which COVID-19 was mentioned on the death certificate.

[12] Specifically, up to 6th Aug 2021 there were 132369 deaths in England and 140518 in England and Wales giving a factor 0.942.

[13] When the weekly death rate becomes very low (less than 5 deaths per week), several weeks data are amalgamated until this limit is exceeded. This proved an effective way of reducing Poisson noise in the data.

**Care home data** – ONS provides two data sources regarding care home deaths. Firstly, [2] provides information about the location of deaths. These locations are divided into: "Hospital" (69.1% of all deaths), "Care Home"(22.9%), "Home" (5.8%), "Hospice"(1.6%), "Other communal establishment"(0.3%) and "Elsewhere" (0.4%). The results for "Hospital" and "Care Home" are shown by the blue and green lines in Fig. F1.

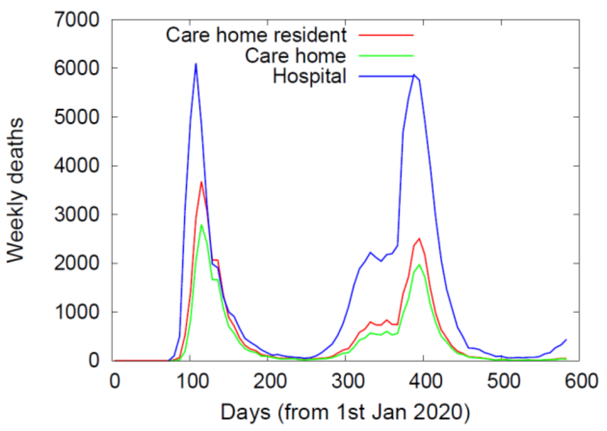


**Figure F1: Death data.** The weekly deaths in England and Wales for care home residents (red), irrespective of whether that death is in a care home or hospital, within care homes (green) and within hospitals (blue).

Secondly, [20] provides the weekly number of deaths from care home residents (irrespective of whether those deaths occur in care homes or hospitals). This is shown by the red line in Fig. F1. By comparing the areas under the red and green curves we calculate that 25% of care home patients actually die in hospital. The red curve is used as the weekly death rate for care home residents, *i.e.* for the "CH" category in our model[14]. Since care homes make up their own category, it is necessary to remove their contributions from the other data sources. We describe this process is the sections below.

#### iii) Survey data

The Coronavirus Infection Survey (CIS) [3] is the largest regular survey of coronavirus infections and antibodies in the UK, providing vital information to help in the UK's response to the pandemic. Within CIS households are randomly selected from the population and invited to take part (note, people living in care homes, other communal establishments and hospitals are not included). CIS carries out two types of test as described below:

**CIS PCR test data** – Nose and throat PCR tests are taken to establish if individuals are currently infected (this is expected to pick up asymptomatic carriers of the disease, so providing crucial information not obtainable from other forms of data). The data published by CIS provides an estimate for the percentage of the population that are PCR positive at fortnightly intervals. This data is age-stratified into the following age bands: "Age 2 to School Year 6", "School Year 7 to School Year 11", "School Year 12 to Age 24", "Age 25-34", "Age 35-49", "Age 50-69", "Age 70+". These age bands are transformed into the 5 yearly intervals required by our analysis using the method described in Appendix G. This informs the total population in the $\mathrm{I}$, $\mathrm{A}$, $\mathrm{C}$ and $\mathrm{T}$ compartments.

**CIS seroprevalence data** – Blood samples are taken from ~20% of the study's participants (of those aged 16 and above), and these are tested for the presence of COVID-19 antibodies [21]. This data is complicated by the fact that, for the most part, these measurements have been taken whilst the vaccination programme has been underway. Since the serological test is unable to distinguish between a vaccinated individual or someone who has become infected with COVID-19 we use only the very earliest set of measurements. In particular, we focus on data taken on two dates: On 10th December 2020 (before seroconversion of the first vaccinated individuals) data is reported in the following age bands: "Age 16-24", "Age 25-34", "Age 35-49", "Age 50-59", "Age 60-64", "Age 65-69", "Age 70-74", "Age 75-79" , "Age 80+". Because the five highest age groups coincide with the age bands used in our study, we use these results directly. On 7th January 2021 estimates for yearly age

[14] This curve is multiplied by a factor 0.942 to transform it from data for England and Wales to just England (this factor comes from the ratio of total deaths in England to those in England and Wales).

bands are published [22], and these are used to generate data for the younger age groups[15] (up to 59 years old). Although this data does come from after the start of the vaccination programme, it can still reliably be used because very few individuals in the younger age groups had been vaccinated by this date (the initial focus for vaccination was on the vulnerable elderly and individuals in care homes). Seroprevalence data informs the total population in the recovered R compartment[16].

For simplicity we assumed that both the PCR and antibody tests are perfect, as estimates for their sensitivity and specificity are both high [23, 24].

#### Adjusting overall death data to give hospital death data

We now describe how COVID-19 deaths in various age groups published by the ONS [2] are converted to give only hospital deaths for the general population (*i.e.* by removing care home residents already accounted for, as well as individuals who die at other locations).

Consider first removal of care home residents. Assuming COVID-19 deaths of individuals in care homes occur in the same proportions as the population at large, the fraction of deaths in different age groups is given by 65-69: 0.6%, 70-74: 1.0%, 75-79: 4.2%, 80+: 94%. Care home deaths are subtracted from the other age groups using these proportions (this ensures that care home deaths are not counted twice).

We now correct for the fact that deaths occur in locations other than hospitals and care homes. Using the percentages from above, we see that 5.8+1.6+0.3+0.4 = 8.1% deaths occur at other locations. Consequently, we multiply the final deaths data by a factor 69.1/(69.1+8.1)=0.90 to make it representative of hospital deaths only.

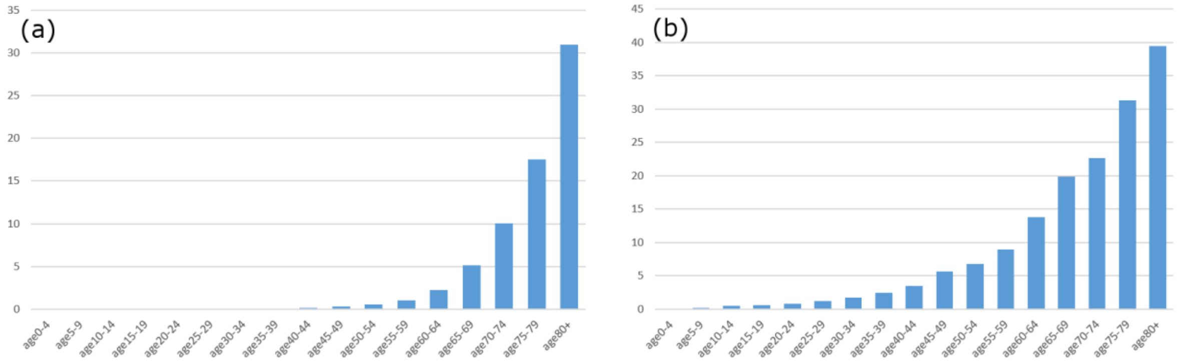


**Figure F2:** Age-stratification for: (a) the case fatality rate (CFR), estimated by dividing the total number of deaths by the number of cases[17], and (b) the hospitalised fatality rate (HFR), estimated by dividing the total number of deaths by the number of hospital admissions[18] and allowing for a factor 0.69 to account for the fraction of deaths occurring in hospitals.

#### Removing care home contribution to case data

No public information is available specifically for case numbers in care home resident. It was necessary, therefore, to make an informed guess as to the number of cases to remove from the

---

[15] These are adjusted back to 10th Dec 2020 by multiplying by a factor given by the total number of COVID-19 hospital admissions up until 10th Dec 2020 divided by the total number until 7th January 2021 (see Fig. P2).

[16] Seroconversion takes typically a couple of weeks, so individuals in other compartments would not test positive for antibodies, despite the fact they are infected.

[17] This was calculated for the second wave between 28th August 2020 to 1st July 2021.

[18] This was calculated for the second wave between 16th October 2020 to 1st July 2021.

general population. Figure F2(a) shows the overall case fatality rate (CFR) for the population. Although care home residents cover a broad range of ages, because they can be expected to be more frail than the population at large, we consider them (from an epidemiological point of view) to behave the same as the “80+” group in the general population. Using the CFR of 31% for the “80+” group along with the death curve in the “CH” group (see the red curve in Fig. F1), shifted by 16 days to account for the difference in time between being a case and dying, we estimate the number of cases to remove from the general population as a function of time. The age distribution of this removal is done in proportion to the age groups in the care homes themselves.

#### Removing care home contribution to hospital admission data

In a similar fashion, the contribution of care home residents to hospital admissions is removed. Above it was estimated that 25% of care home residents die in hospital. Figure F2(b) shows the hospitalised fatality rate (HFR). As before, we use the value of 40% from the “80+” group as representative of care home residents. Finally, we use the shifted death curve for the “CH” classification, multiplied by a factor 25%/40%, to estimate the number of care home admissions, which are then removed from the data for the general population.

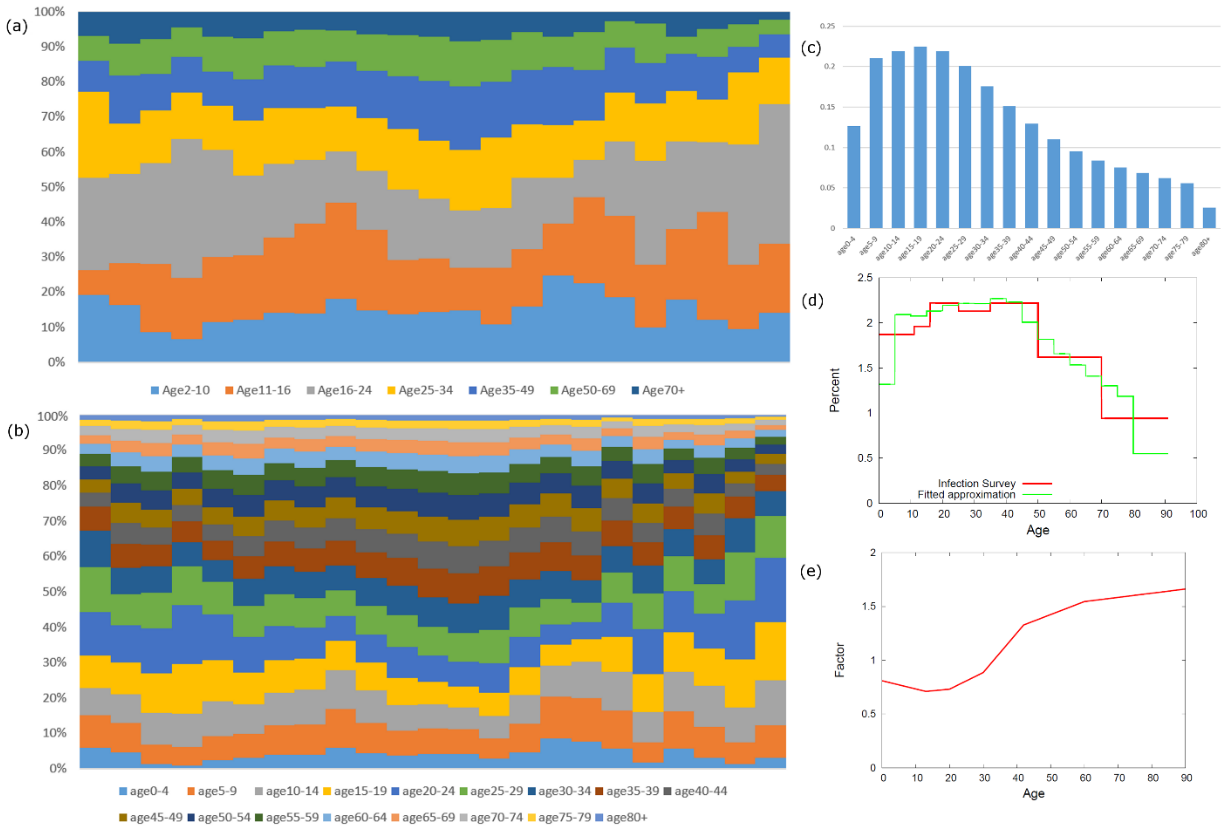


**Figure G1**: Proportions of COVID-19 PCR positive individuals in different age bands changes in time for: (a) the age bands used in published data and (b) transformed 5 year age bands used for the analysis in this paper. These plots are made up of columns representing fortnightly intervals. The left edge corresponds to the 23rd August 2020 and the right edge 27th June 2020. (c) A representative distribution for the percentage of PCR positive individuals. (d) The transformation from the original set of age bands (red) to the new age bands (green). (e) An age-varying factor multiplying the representative distribution to obtain the modified approximation.

#### Appendix G: Transforming data into 5 year age bands

Unfortunately, not all time series summary data (published by the various organisations) are in the 5 year age bands required by the data analysis used in this paper. This appendix describes how the raw data is transformed into the required format.

An example is provided in Fig. G1(a), which shows how the proportion of PCR positive individuals in different age bands changes as a function of time (data provided by the Coronavirus Infection Survey (CIS) [3]). To transform this to the required age bands (as shown in Fig. G1(b)) a representative distribution is needed[19]. Fortunately, CIS provides [2] data stratified by yearly age intervals for a limited period of the study's duration. Averaging over data provided between 23rd May 2021 and 3rd July 2021 yields the representative distribution in Fig G1(c).

Transformation from Fig. G1(a) to Fig. G1(b) consists of taking each column in turn and calculating how the representative distribution should be "distorted" to make it fit this data as closely as possible. This fitted distribution is then used to generate the corresponding column in Fig. G1(b).

Let's look at an example. The red curve in Fig. G1(d) shows results for a single time point from the raw data. We define a linear spline *f* (defined at yearly age intervals, as shown in Fig. G1(e)) that multiplies the representative distribution[20] to give the new one. We define an error function that determines how close the distorted distribution agrees with the data in the existing age bands, as well as ensuring some level of smoothness on *f*:

$$EF = \sum_{b_{old}} \left[ \left( \sum_{a \in b_{old}} d_{b_{new} \in a} f_a p_a \right) - \left( \sum_{a \in b_{old}} r_{b_{old}} p_a \right) \right]^2 + \kappa \sum_{j=2}^{J-1} \left( f_{j-1} + f_{j+1} - 2 f_j \right)^2, \qquad \text{(G1)}$$

where $b_{old}$ goes over the existing age bands, $r_{b_{old}}$ is the data measurement for band $b_{old}$, and $a$ sums over individual ages within $b_{old}$. The quantity $d_{b_{new} \in a}$ gives the value for the representative distribution in the new age band corresponding to age $a$. For a given age $a$, $f_a$ is the value of the spline and $p_a$ is the population size (obtained from ONS [25]). The parameter $\kappa$ sets the smoothness of the spline[21] and $j$ sums over the intermediate break points of the spline (which have values $f_j$).

The values for $f_j$ that minimise the error function in Eq. (G1) were found using a gradient descent algorithm. The results in the new age bands are calculate using

$$r_{b_{new}} = \frac{\sum_{a \in b_{new}} d_{b_{new} \in a} f_a p_a}{\sum_{a \in b_{new}} p_a}. \qquad \text{(G2)}$$

This final result is represented by the green curve in Fig. G1(d). This fitting procedure is repeated for all the columns in Fig. G1(a) to generate the transformed data in Fig. G1(b).

---

[19] Note, this is just a single instance, not a time series.

[20] This has break points at the mid-points of the existing age categories as well as at zero and 90 (the oldest age group considered).

[21] This was set to 0.001, which, by visual inspection, provided a moderate level of smoothing but was not too restrictive.

### Appendix H: Raw data analysis

Figure H1 plots curves showing how different data sources vary as a function of time (note, other than cases, these curves have been scaled to see how similar the profiles are to each other).

Overall, the profiles agree well, demonstrating that each measure can, in principle, be used as a reliable indicator to track the progress of the epidemic. We also see the expected ordering based on progression of the disease: infections (from CIS), followed by cases, then hospital admissions, and, finally, deaths[22].

There are, however, some discrepancies between these curves. In particular, referring to the annotations in Fig. H1:

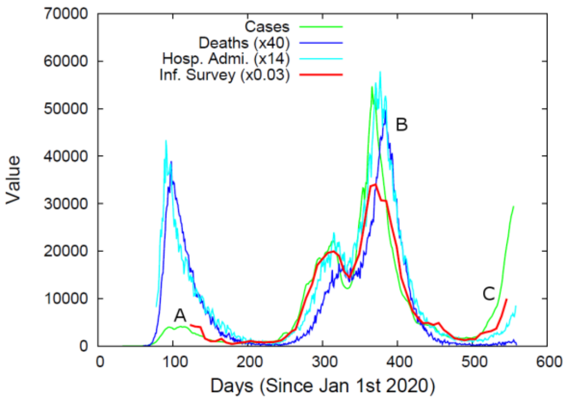


**Figure H1: Publically available data**. This shows daily results for cases (green, Gov.uk [1]), deaths (blue, ONS [2]), hospital admissions (cyan, Gov.uk [13]) and estimated infection levels within the general population (red, CIS [3]). Curves other than cases have been scaled to allow for comparison.

A) The case data (green curve) during the first wave is clearly much lower than would be expected from the other curves. This is primarily due to a lack of testing capacity during the early period of the pandemic. For this reason, only case data from the second wave is used in the analysis (from 8th August 2020 onwards). Furthermore, results from CIS were relatively unreliable early on due to a small number of participants. Data is only used from 23rd August 2020 onwards.
B) The peak in the cases, hospital admissions, and deaths rise above the level shown from CIS. This is largely due to significant numbers of infections and deaths occurring in care homes during this period which are unaccounted for in CIS (CIS only detects infection levels in private residences, not care homes).
C) We see a substantial reduction in deaths compared to infections. This is a direct result of the vaccination program, which has been particularly effective at reducing deaths in the elderly.

### Appendix I: Weighting observation in the error function.

This appendix outlines how the weights $w_i$ on different observations $i$ in Eq.(6) are specified. The first thing to mention is that we expect measurements $y_i$ that have a higher value to be more accurate (*i.e.* less prone to statistical fluctuation) than those which have a lower value. For example, if we consider a Poisson distributed variable, the uncertainly of an observation goes up with the square root of its size. Following this principle, we start by setting weights according to:

$$w_i = \sqrt{y_i} \qquad \text{(J1)}$$

In practical terms this means that when fitting data to the model, more emphasis is placed on getting the peaks of time series right and less emphasis is placed elsewhere, *e.g.* in the tails of distributions.

The expression in Eq.(J1), however, is not quite right. To understand why, we have to remember that the data being fitted to the model is in the form of sets of time series measurements on different

[22] This is particularly noticeable at the peak of the second wave.

system quantities (*e.g.* see the black lines in Fig. R1 in the posterior Supplementary Results). Some time series relate to weekly deaths in certain age groups, and others give the estimated infected population as a function of time *etc*... Now the first problem is that the scales on these time series may be completely different, *e.g.* population numbers will likely be far larger than death rates. Consequently, naïve application of Eq.(J1) would result in far more weight being placed on the former over the latter. The second problem is that some time series contain many more observations than others, *e.g.* cases are measured daily, but data from only a single time point is available for the antibody test results. A simple way to overcome both of the difficulties is the following:

$$w_i = \frac{\sqrt{y_i}}{\sum_j \sqrt{y_i}}, \tag{J2}$$

where $j$ sums over all measurements that share the same time series as $i$. This ensures that each time series gets the same overall weighting of one.

As a result of the weighting in Eq.(J2), when the ABC-MBP algorithm iterates through generations the overall error function is distributed roughly evenly across the different sources of data.

The small constants $\varepsilon_{g_i}$ in Eq.(6) are chosen to have 2% the value of the highest point within a given time series $g_i$. This choice is somewhat arbitrary, but results are found to be relatively insensitive to this particular value.

### Appendix J: Prior specifications

The prior $\pi(\theta)$ captures the state of knowledge regarding model parameter values before data $y$ is considered. Prior specifications for the analysis in the paper are shown in Table R1 of the posterior Supplementary Results. Below we discuss why these selections have been make:

**Mean residency times** $m_{c,a}$ – Because, for the most part, these can't be estimated from the data they are set to fixed values taken from the literature (see Appendix K) with the exception of the residency time in the PCR test sensitive compartment T. $m_{T,a}$ are given uniform prior distributions with wide, realistically plausible bounds (between 4 and 20 days). Because CIS only provides seroprevalence data for ages 16 and above, the four age groups up to 19 are all assumed to have the same residency time $m_{T,0\text{-}19}$.

**Branching probabilities** $b_a^{trans}$– These are mostly given uninformative Dirichlet priors[23], with the exception of $b_a^{I\to\mathrm{C}}$ and $b_a^{H\to\mathrm{D}}$, which are biased toward being small in the younger age groups (in reality these quantities are very small, as demonstrated by inference). Because the only source of information for care home residents comes from death data, we make the prior assumption that, from an epidemiological viewpoint, individuals in the CH classification behave the same as in the "80+" category (*i.e.* we set $b_{CH}^{E\to\mathrm{A}}$ to be equal to $b_{80+}^{E\to\mathrm{A}}$, $b_{CH}^{I\to\mathrm{C}}$ to be equal to $b_{80+}^{I\to\mathrm{C}}$, and $b_{CH}^{H\to\mathrm{D}}$ to be equal to $b_{80+}^{H\to\mathrm{D}}$).

**Age contact factors** $v_a$ – These are restricted to have a weighted average of 1 (see Eq.(2)). Correspondingly, a so-called "modified" Dirichlet prior[24] is applied. This means that $v_a(P_a/P)$ have a

---

[23] These are used because branching probabilities summed over all transitions leaving a compartment are required to strictly add to one.

[24] A standard Dirichlet applies to quantities required to strictly add to one, *e.g.* probabilities. This modified version applies to cases when a weighted sum must add to 1, as is the case in Eq.(2).

Dirichlet prior Dir($\alpha_a$), where $P_a/P$ is the population fraction in the age group $a$. The alpha values are set according to

$$\alpha_a = \left(\frac{A-1}{\sigma^2} - 1\right)\frac{P_a}{P}, \qquad \text{(J3)}$$

where $A$ is the number of age groups and $\sigma$ is a hyperparameter that sets the strength of the prior (the notation MDir($\sigma$) in Table R1 of the supplementary results shows that $\sigma$=0.5 for this particular analysis). This choice leads to the prior for $v_a$ having a mean of one and a variance

$$\sigma^2 \frac{\frac{P}{P_a} - 1}{A-1}. \qquad \text{(J4)}$$

In the case in which all groups have the same population size, this simplifies to a variance of $\sigma^2$ (or equivalently a standard deviation of $\sigma$). Correspondingly, the choice $\sigma$=0.5 represents a relatively uninformative prior choice (allowing for a potential large ~50% variation in age contact factor).

**Spline for $R_t$** – Parameters $R_d$, which give the value of the reproduction number $R_t$ at spline points indexed by $d$ (up to a maximum of $D$), are given priors with wide uniform distributions. Additionally, a smoothing prior is applied:

$$\pi^{spline}(\theta) = \sum_{d=1}^{D-1} \mathrm{LN}\left(R_{d+1} \mid R_d, \kappa_d^2\right), \qquad (5)$$

where LN($x|m$,σ[2]) represents the log-normal probability of $x$ given a mean $m$ and variance $\sigma^2$. This proved useful at suppressing unphysical oscillations in the reproduction number[25]. The strength of smoothing is determined by the parameter $\kappa_d$. For most points $d$ this was set to $\kappa_d$ =0.2 (corresponding to a prior belief that a 20% change in reproduction number over a two-week interval is reasonable), but it was set to $\kappa_d$ =0.5 at the three lockdown time points [26] (allowing for a potential 50% change, reflecting a prior belief that lockdowns can have a more substantial effect).

#### Appendix K: Fixed parameter estimates

This appendix explains how various parameters in the model are fixed (*i.e.* those which cannot be estimated from the available data) based on figures published in the literature:

| Param. | Value | Description | Source |
|---|---|---|---|
| $m_E$ | 3.8 days | E mean residency time. | The average inoculation period has been estimated by McAloon *et al.* [27] as 5.8 days. Subtracting off an estimated presymptomatic period of 2 days [17] gives a latent period of 3.8 days. |
| $m_I$ | 4 days | I mean residency time. | Whilst 4 days for the infectious period is considerably shorter than published estimates (*e.g.* Byrne *et al.* [17] gives an average infectious period for symptomatic patients of 13.4 days) it is necessarily small to provide the model with a realistic generation time. A meta-analysis by the Royal Society [28] gave estimates for the |

[25] Despite being unrealistic, a system with $R_t$ oscillating between high and low with a high frequency behaves almost exactly the same as if the value of $R_t$ is constant.

| | | | |
|---|---|---|---|
| | | | generation time in the range 4-5 days (with some degree of uncertainty) and serial intervals in the range 4-7 days (which included many more studies). The choice of $m_I$ and $m_A$ given here corresponds to a generation time of 5.8 days. |
| $m_A$ | 4 days | A mean residency time. | For simplicity, this is chosen to be the same as $m_I$ (asymptomatic individuals do not isolate, so they would perhaps be expected to have a longer infectious period, but on the other hand they experience a less severe disease and so would be expected to recover faster). |
| $m_C$ | 3.1 days | C mean residency time. | Pellis *et al.* [29] used Public Health England (PHE) data to estimate the time from the onset of symptoms to hospital admission as 5.1 days. Adding on the presymptomatic period of 2 days and subtracting off the infectious period of 4 days finally gives 3.1 days. |
| $m_H$ | 13 days | H mean residency time. | Byrne *et al.* [17] gives a mean duration from symptom onset to hospital discharge or death of 18.1 days and a presymtomatic period of 2 days. Adding these two figures and subtracting off the infectious period $m_I$ gives 13 days. |
| $\varphi_I$ | 1 | Relative infectivity of I. | By definition, this is set to one (it represents the reference to which the infectiousness of other compartments is measured). |
| $\varphi_A$ | 0.55 | Relative infectivity of A. | This value is taken from D. McEvoy *et al.* [30]. |

#### Appendix L: Model-based proposals

This appendix provides information on how model-based proposals (MBPs) [31, 32] are used to change the state of particles. MBPs follow a procedure that mirrors how simulation of the system is performed, so we first direct the reader to Appendix E for an explanation of this.

We take $\theta^s$ and $\xi^s$ to be the current parameter set and system state for a given particle. Time $t$ is discretised into time steps of size $\tau$. Within $\xi^s$ we denote $p^s_{a,c,t}$ to specify the time variation in the population of compartment $c$ and age group $a$, and $n^s_{a,j,t}$ to denote the number of transitions of type $j$ between times $t$ and $t+\tau$.

From the model the number of transitions $n^s_{a,j,t}$ is taken to be Poisson distributed with a mean value given by:

$$\omega^s_{a,j,t} = \left. \begin{array}{c} \tau p^s_{a,S,t} \lambda^s_{a,t} \\ \tau p^s_{a,i_j,t} b^{s,j}_a / m^s_{i_j,a} \end{array} \right\} \begin{array}{c} \text{If } j \text{ is infection I} \to \text{E} \\ \text{Otherwise} \end{array} . \quad \text{(L1)}$$

The superscript $s$ on the right-hand side indicates that these are parameters and quantities taken from current particle state $\theta^s$ and $\xi^s$. In Eq.(L1) the force of infection $\lambda^s_{a,t}$ is obtained from Eq.(1), $b^{s,j}_a$

is the branching probability for individuals to move down transition $j$ (as opposed to any other transition leaving the initial compartment $i_j$) for age group $a$, and $m^s_{i_j,a}$ is the residency time in compartment $i_j$ (*i.e.*, the compartment from which the individuals are leaving).

Two types of MBP are used to make changes to $\theta^s$ and $\xi^s$:

#### Type I: Joint changes to θ and ξ

A new parameter set $\theta'$ is proposed from $\theta^s$ (see below for different ways in which this can be achieved). The proposed state $\xi'$ is generated in the following way:

1) **Initialisation** – The time $t$ is set to $t_{\text{start}}$. The initial population for the proposed state is set[26] $p'_{a,c,t_{\text{start}}}$.
2) **Sample transition numbers** – We go through each age group $a$ and transition $j$ in turn and calculate $\omega'_{a,j,t}$, the expected mean number of transitions in the proposed state (this is the same as Eq.(L1) but with the superscript $s$ replaced with a prime).
   The transition numbers in the proposed state $\xi'$ are sampled according to:

$$\left.\begin{array}{l} n'_{a,j,t} = n^s_{a,j,t} + X \quad \text{where } X \sim \text{Poisson}(\omega'_{a,j,t} - \omega^s_{a,j,t}) \\ n'_{a,j,t} \sim \text{Binomial}(n^s_{a,j,t}, \omega'_{a,j,t} / \omega^s_{a,j,t}) \end{array}\right\} \begin{array}{l} \text{If } \omega'_{a,j,t} > \omega^s_{a,j,t} \\ \text{Otherwise} \end{array}, \tag{L2}$$

   where Binomial($n$,$p$) samples from the binomial distribution with $n$ trials each accepted with probability $p$.

3) **Update populations** – Compartmental populations in $\xi'$ are updated according to:

$$p'_{a,c,t+\tau} = p'_{a,c,t} + \left[\sum\nolimits_{j \in \text{enter c}} n'_{a,j,t}\right] - \left[\sum\nolimits_{j \in \text{leave c}} n'_{a,j,t}\right], \tag{L3}$$

   where the first sum goes over all transitions that enter compartment $c$ (increasing its population), and the second goes all those that leave $c$ (decreasing its population).

4) **Iterate** – Increment $t$ by $\tau$ and if less than $t_{\text{end}}$ go back to step 2.

Having generated the proposed particle state $\theta'$ and $\xi'$ the error function from Eq.(6) is calculated. If this is greater than the cut-off value, *i.e.* $\text{EF}(y|\xi') \geq \text{EF}_{\text{cutoff}}$, the proposal is immediately rejected. If not, it is accepted (*i.e.*, we set $\theta^s=\theta'$ and $\xi^s = \xi'$) with Metropolis-Hastings probability

$$\max\left\{\frac{\pi(\theta')}{\pi(\theta^s)}, 1\right\}. \tag{L4}$$

Note, this equation does not contain the latent process likelihood and so MBPs can be classed as "likelihood-free".

#### Type II: Changes to ξ with fixed θ

These proposals aim to make changes to the state $\xi^s$ for a fixed set of parameters $\theta^s$.

As they are iterated in time they actually intersperse "simulation" steps, of the type given in Eqs.(E1) and (E2), with "modification" steps, of the type given in Eq.(L2).

---

[26] By default, all individuals are assumed to start in the susceptible compartment S. The population sizes for each age group are taken from the census data described in the main paper.

To capture this first a vector $\mathrm{V}_t$ is defined (see below for how this is done in practice), where $\mathrm{V}_t$='sim' if a simulation step is performed at time $t$ and $\mathrm{V}_t$='mod' if a modification is performed instead.

The proposed state $\xi'$ is generated in the following way:

1) **Initialisation** – The time $t$ is set to $t_{\text{start}}$. The initial population for the proposed state is set $p'_{a,c,t_{\text{start}}}$.
2) **Sample transition numbers** – We go through each age group $a$ and transition $j$ in turn and calculate $\omega'_{a,j,t}$, the expected mean number of transitions in the proposed state (this is the same as Eq.(L1) but with the superscript $s$ replaced with a prime).
   If $\mathrm{V}_t$='sim', the transition numbers in the proposed state $\xi'$ are sampled according to:

$$n'_{a,j,t} \sim \text{Poisson}(\omega'_{a,j,t}), \tag{L5}$$

   or if $\mathrm{V}_t$='mod':

$$\left.\begin{array}{c} n'_{a,j,t} = n^s_{a,j,t} + X \quad \text{where } X \sim \text{Poisson}(\omega'_{a,j,t} - \omega^s_{a,j,t}) \\ n'_{a,j,t} \sim \text{Binomial}(n^s_{a,j,t}, \omega'_{a,j,t} / \omega^s_{a,j,t}) \end{array}\right\} \begin{array}{c} \text{If } \omega'_{a,j,t} > \omega^s_{a,j,t} \\ \text{Otherwise} \end{array} . \tag{L6}$$

3) **Update populations** – Compartmental population in $\xi'$ are updated according to:

$$p'_{a,c,t+\tau} = p'_{a,c,t} + \left[\sum_{j \in \text{enter c}} n'_{a,j,t}\right] - \left[\sum_{j \in \text{leave c}} n'_{a,j,t}\right], \tag{L7}$$

   where the first sum goes over all transitions that enter compartment $c$ (increasing its population), and the second goes all those that leave $c$ (decreasing its population).

4) **Iterate** – Increment $t$ by $\tau$ and if less than $t_{\text{end}}$ go back to step 2.

Having generated the proposed particle state $\xi'$ the error function from Eq.(6) is calculated. If this is greater than the cut-off value, *i.e.* $\text{EF}(y|\xi') \geq \text{EF}_{\text{cutoff}}$, the proposal is rejected, otherwise $\xi^s = \xi'$.

#### A particle "update"

When the ABC-MBP algorithm is run each particle within each generation undergoes an "update". This consists of a series of MCMC proposals that allow the particle to explore parameter and state space subject to the error function being confined to below a cut-off. Below we outline the set of proposals used:

**Univariate type I MBPs** – Each parameter $k$ in the model is taken in turn and undergoes $N$ type I MBPs (see above). In each of these proposals, the parameter set $\theta'$ is generated by changing only parameter $k$ and sampling its value from a normal distribution centred at the parameter's current value:

$$\theta'_k \sim \text{Normal}(\theta^s_k, \sigma^2_k). \tag{L8}$$

The standard deviation $\sigma_k$ is dynamically tuned to give an acceptance probability of around 33%. The number of proposals $N$ is tuned by

$$N = \frac{\sigma^2_k}{\Sigma_{kk}}, \tag{L9}$$

where $\Sigma_{kk}$ is the estimated variance of parameter $k$ across all particles. This ensures more proposals are made when only relatively small jumps in parameter space are possible (consequently, more and more proposals are required as the number of generations increases).

**Multivariate type I MBPs** – For the most part posterior correlations between model parameters were found to be relatively small (hence the univariate proposals from above were sufficient). One exception to this is for parameters controlling the spline for $R_t$. Here two additional types of proposals were used to help promote mixing:

*Neighbouring spline points* – Each spline point $q$ is selected in turn (apart from the end point) and the proposed parameter set $\theta'$ is generated by first sampling a scaling factor $a_q$ from a normal distribution and then multiplying the spline point $R_q$ by a factor $e^{a_q}$ and dividing the neighbouring spline point $R_{q+1}$ by that same factor, *i.e.*

$$a_q \sim \text{Normal}(0,\sigma_q^2), \quad R'_q = R_q e^{a_q}, \quad R'_{q+1} = R_{q+1} e^{-a_q}. \tag{L10}$$

The standard deviation $\sigma_q$ is dynamically tuned to give an acceptance probability of around 33%.

*Sinusoidal variation* – Separate proposals are made for wavelengths ranging from $\lambda$=1 up to $\lambda$=11 (distances are here measured using the spline index $q$). For each proposal the parameter set $\theta'$ is generated by adding a sinusoidal variation to the existing values on the spline:

$$R'_q = R_q + d_\lambda \sin\left(\tfrac{2\pi}{\lambda} q + \phi\right), \tag{L11}$$

where $\phi$ is a randomly sampled phase (between 0 and 2π). The magnitude $d_\lambda$ is, again, dynamically tuned to give an acceptance probability of around 33%.

**Type II MBPs** – The aim of these is to propose changes to the particle state $\xi^s$ for a fixed set of parameters $\theta^s$. These changes are accepted conditional on the error function being below a cut-off. In the early generations of the algorithm this is relatively easy, because the proposed state can be simply generated through simulation (here the cut-off is relatively large, and so the simulated state will usually still have an error function below this threshold, resulting in a high acceptance probability). Direct simulation of the state is the same as performing a type II MBPs with $V_t$='sim' for all $t$.

As the generations increase, however, such proposals become increasingly likely to be rejected as the error function cut-off becomes smaller and smaller. The algorithm dynamically tackles this situation in the following way. At the end of each generation it looks at the rejection rate of the type II MBPs (across all particles). If it exceeds 30%, the single global simulation proposal is split in two new type II proposals applied to subsequent generations: 1) Simulate in the first half followed by modification in the second half (*i.e.*, $V_t$='sim' for $t<t_{\text{mid}}$ and $V_t$='mod' for $t \geq t_{\text{mid}}$) and 2) modification in the first half followed by simulation (*i.e.,* $V_t$='mod' for $t<t_{\text{mid}}$ and $V_t$='sim' for $t \geq t_{\text{mid}}$), where $t_{\text{mid}}$ is the mid-point between the system start and end time. These new proposals have a higher acceptance probability than the original one that simulated the entire system. As further generation pass, however, these new proposals many also have a rejection rate exceeding 30%. At that point, they are, themselves, split in two according to the midpoint of their simulation time and these are used in subsequent generations. Consequently, as generations increase, so more and more type 2 MBPs are used which simulate over smaller and smaller time periods (with other time points set to modification).

To help improve mixing, proposals that make up an “update” are randomly ordered (*i.e.* such that univariate type I, multivariate type I and type II MBPs are all interspersed).

#### Appendix M: BEEPmbp software package

“Bayesian Estimation of Epidemic Parameters using Model-Based Proposals” (BEEPmbp) is an open source software tool for fitting epidemiological models to data. It allows for arbitrary specification of the compartmental model and supports potential spatial and/or demographic stratification of the population. To perform inference BEEPmbp accepts a variety of population-based data (time-series giving rates of transitions, populations in different compartments and marginal distributions). Priors on model parameters can be specified from a large range of possibilities.

BEEPmbp implements the ABC-MBP algorithm outlined in this paper, as well as alternative approaches such as particle MCMC, ABC-SMC and MC[3]. A manuscript is currently in preparation that will provide a comprehensive comparison between these various approaches (*e.g.* investigating relative computational speed and accuracy of posteriors obtained on a number of benchmark systems), highlighting the advantages of ABC-MBP.

The code for BEEPmbp, along with a manual, is available on GitHub here. It is designed to run on the command line in Linux and implements efficient parallelisation for use on high performance computing (HPC) facilities[27]. Inference using BEEPmbp is run by specifying three things: 1) an initialisation file that provides all the details of the model, the file names of data tables to be loaded, prior specifications, and, finally, details describing how outputs should be plotted, 2) the data files themselves, and 3) specification of the inference algorithm (along with quantities used to run that algorithm).

For the analysis in this paper the initialisation file is named “England_AS.toml”, which can be found here along with the data files in the directory “Data_England_AS”. This analysis can be replicated by cloning the GitHub repository and executing the following command:

```
mpirun –n 256 ./beepmbp inputfile="COVID 19 England/England_AS.toml" mode="abcmbp"
nparticle=16 nrun=16 ngeneration=350
```

This specifies that the ABC-MBP inference algorithm is to be used and that it should be run 16 times with 16 particles in each run for a total of 350 generations. Here 256 refers to the number of cores used for the computation (which would change depending on the available computing facilities). This choice corresponds to a minimum execution time, because each particle on each run is executed on a separate CPU core[28].

#### Appendix N: Validation and sensitivity analysis

This appendix first demonstrates that the inference algorithm is able to provide accurate estimates for model parameters based on realistic simulated data, and then goes on to look at the sensitivity of the inferred results to changes in the model’s fixed parameter values[29].

---

[27] This makes use of Message Passing Interface (MPI).

[28] Information is transferred between cores using MPI after each generation.

[29] In particular, for two sets of key parameters: the external force of infection and the average residency times for the model compartments.

#### Validation

The model was simulated using posterior mean estimates for parameter values taken from Table R1 in the posterior Supplementary Results. Substantial stochastic variability was found across different simulations, so a selected realisation was chosen that closely resembled the real data[30]. Data with the same information as those from the real COVID-19 dataset (*e.g.* simulated cases, admissions, deaths, *etc*…) were constructed from the simulated system state. These data were censored in the same way as the real data (*e.g.* case data was available for only the second wave and antibody test results consisted of just a single time point).

Figure N1 shows the results of inference carried out on the simulated dataset (inference is performed in exactly the same way as for the analysis on the real data, but for brevity additional outputs such as those given in the posterior Supplementary Results are not shown here).

We obtain excellent results for the estimated time variation in the reproduction number $R_t$, as shown in Fig. N1(a). The posterior mean red curve almost exactly follows the "true" black curve used to generate the data. This means that inference is able to do a good job of tracking the true time variation in reproduction number, based on the types and quantities of data available for COVID-19 in England. We observed a similar picture across other inferred quantities in Fig. N1(b)-(f), where there is a very good agreement between the horizontal black lines represent the true values and the red columns with error bars that give the posterior estimates.

Interestingly, the precision of the posterior means is found to be somewhat higher than would be expected from the size of the posterior error bars. This, to a large extent, is because the ABC-MBP provides a posterior estimate, not the true posterior itself. If the algorithm were left for more generations, these error bars would be expected to reduce in size. However, the algorithm would take longer and longer to run, and so, as is usual, there exists a balance between accuracy and computational practicality.

Some systematic biases can be observed in Fig. N1, although these effects are relatively small (< 5%). For example, the posterior mean for the asymptomatic branching probabilities in Fig. N1(c) all underestimated their true value. These systematic biases can arise as an artefact of using a finite generation number, or from the imperfect, finite data being used.

#### Sensitivity analyses

First, we look at the external force of infection, as derived in Appendix D. A number of simplifying assumptions went into this derivation, and so an important question to address is how sensitive are the results in the paper to the precise values for this external force of infection? To answer this we doubled $\eta(t)$ and repeated the inference procedure to see what effect this change had. The results are shown in Fig. N2, which exhibits little difference compared to the results in the paper. The biggest disparity is a slight reduction in the estimated basic reproduction number $R_0$[31] (*e.g.* in the early stages reducing from a posterior mean of around 4.0 to 3.7) with the value for $R_t$ later changing less than ~2% [32]. We conclude that the results presented in the paper are robust against inaccuracy in the magnitude of the external force of infection curve.

Next, we look at the effect of changing the fixed residency times in the compartmental model. Modifying these alters the fundamental system dynamics because they result in a shift in the

---

[30] It has two similarly sized peaks and a comparable number of COVID-19 deaths to the real data.

[31] This makes sense, because the extra external infections don't have to exponentially grow at quite such a high rate to account for the observed increase in hospital admissions and deaths.

[32] Essentially indistinguishable for stochastic noise in the posterior estimates themselves.

generation time. To provide an example parameters in the model are changed from $m_E$=3.8, $m_I$=$m_A$=4.0 and $m_C$=3.1 to $m_E$=3.0, $m_I$=$m_A$=2.0 and $m_C$=5.1, which reduces the generation time from 5.8 days to 4 days. Inference was performed using this new parameter set, as shown in Fig. N3. As would be expected, because the generation time has gone down, so the initial values for the reproduction number no longer needs to be so high for the system to match the observed hospital admission and death data (we see that it has reduced from a posterior mean of 4.0 to 3.0). Other inferred parameter values essentially remain unchanged. It can be concluded, therefore, that the results in this paper are largely robust to inaccuracies in the fixed residency times in the compartmental model (barring $R_t$, which becomes rescaled to account for the resulting shift in the generation time).

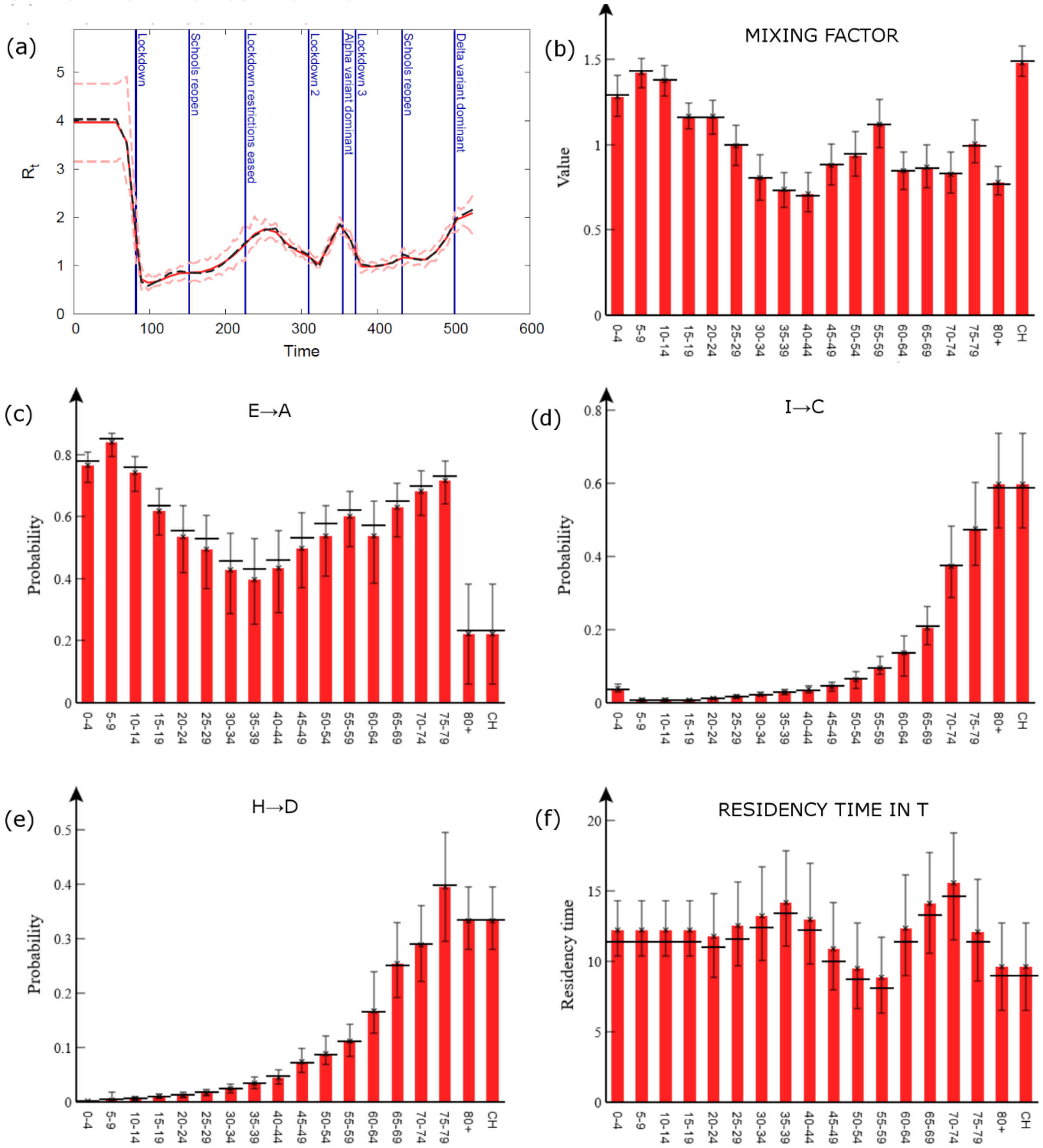


**Figure N1: Validation.** This shows the results of simulating from the model to generate hypothetical data (similar to the real COVID-19 dataset) and then performing inference on that data. (a) The reproduction number $R_t$ (the red solid line gives the posterior mean, the dashed lines denote 95% credible intervals, and the black, dashed line shows the spline used for the simulation). For (b) the age contact factor, (c) the asymptomatic branching probability, (d) the hospitalised branching probability, (e) the death branching probability, and (f) the residency time in the T compartment, the red columns with error bars give the posterior means with 95% credible intervals, and the horizontal black lines show the true parameter values. Good agreement between the black lines and red bars indicates successful inference.

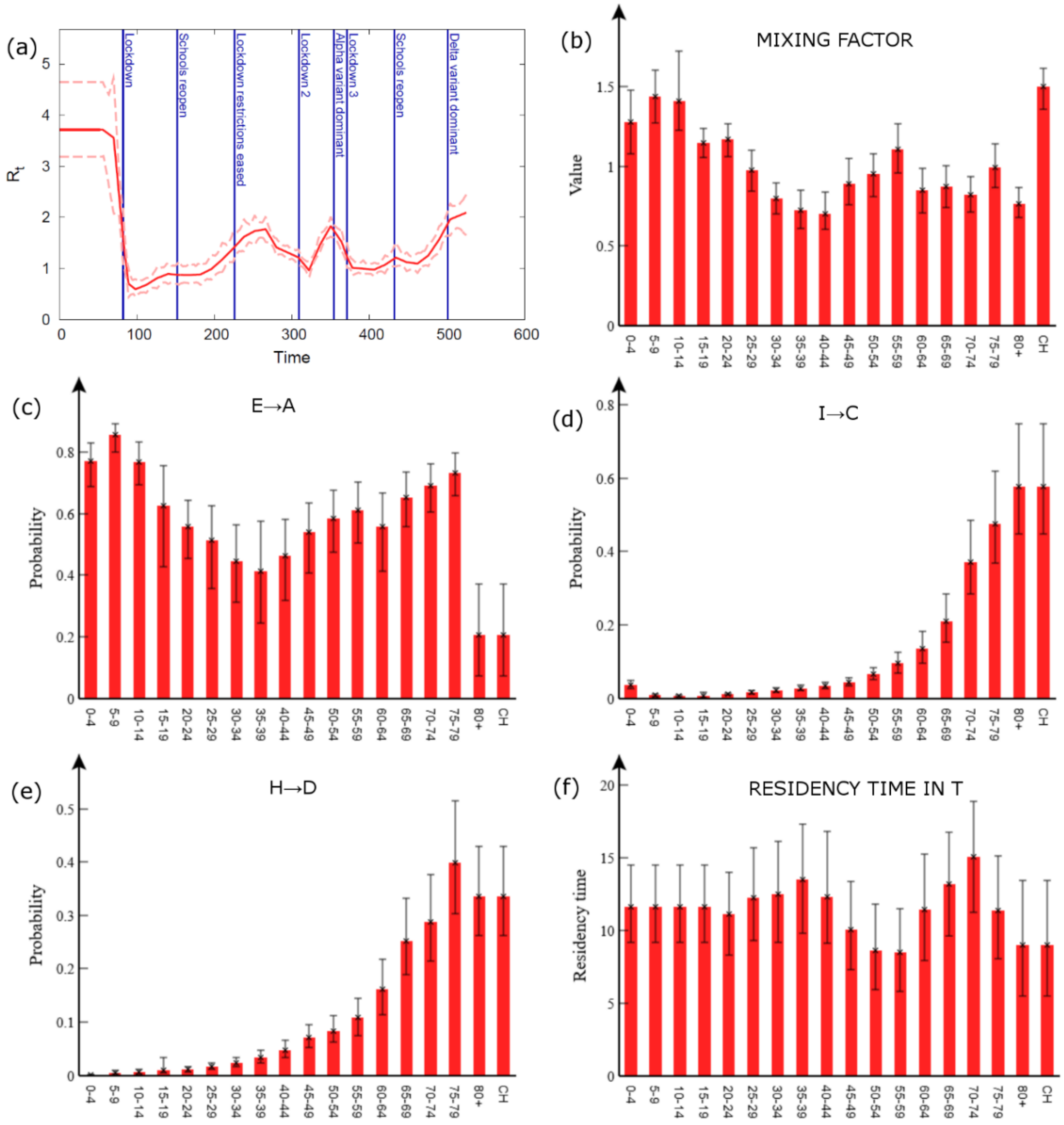


**Figure N2: Doubling external force of infection.** This shows the results of inference assuming the external force of infection is double that used in the paper. (a) The reproduction number $R_t$ (the red solid line gives the posterior mean and the dashed lines denote 95% credible intervals). For (b) the age contact factor, (c) the asymptomatic branching probability, (d) the hospitalised branching probability, (e) the death branching probability, and (f) the residency time in the T compartment, the red columns with error bars give the posterior means with 95% credible intervals.

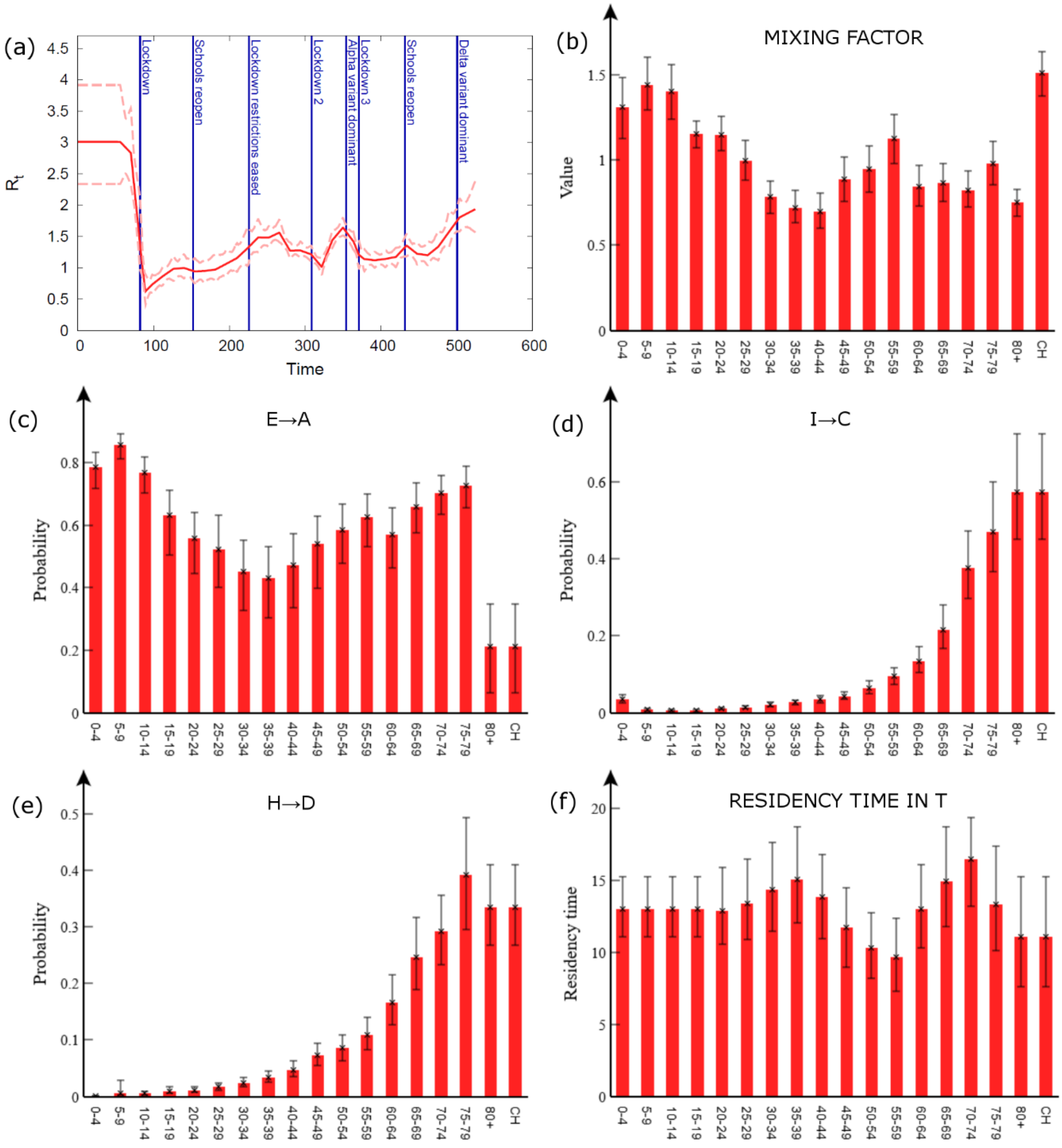


**Figure N3: Changing the generation time.** This shows the results of inference when the generation time is 4 days instead of 5.8 days (as used in the paper). (a) The reproduction number $R_t$ (the red solid line gives the posterior mean and the dashed lines denote 95% credible intervals). For (b) the age contact factor, (c) the asymptomatic branching probability, (d) the hospitalised branching probability, (e) the death branching probability, and (f) the residency time in the T compartment, the red columns with error bars give the posterior means with 95% credible intervals.

### Appendix O: Time variation in COVID-19 variants

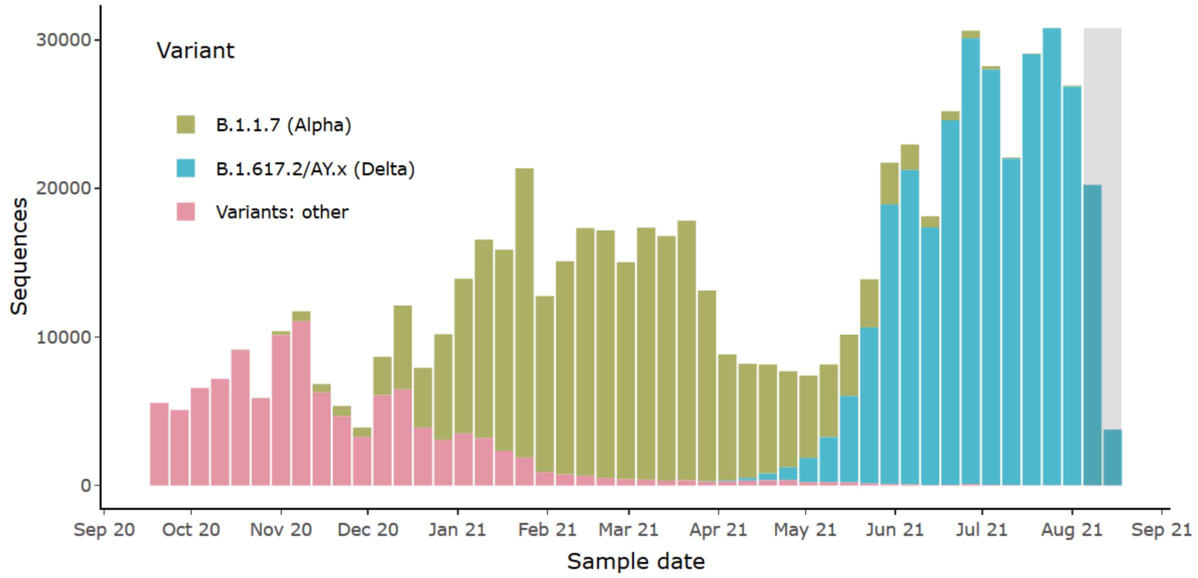


**Figure O1: COVID-19 Variant data**. This figure, taken from the COG UK website [33], shows how the number of samples of different variants has changed as a function of time in the UK. By visual inspection we see that the alpha variant became dominant around 20th December 2020, and the delta variant takes over from 16th May 2021. These correspond to the vertical date lines in the Fig. 4 in the paper.

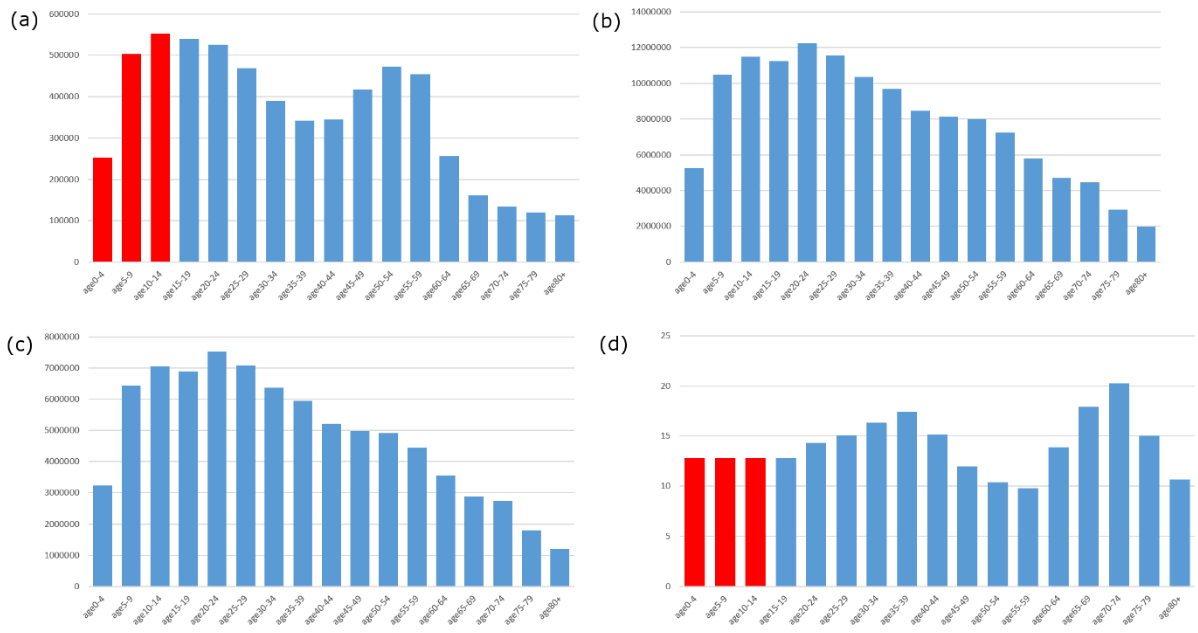


**Figure P1: Data analysis on the Coronavirus Infection Survey in England**. For different age groups: (a) The estimated total number of COVID-19 infected individuals until 29th Nov 2020 based on antibody seroprevalence data (the red bars are imputed, see main text). (b) The PCR positive population integrated in time between 23rd Aug 2020 and 12th June 2021. (c) The estimated PCR positive population integrated in time between 1st Jan 2020 and 29th Nov 2020. (d) The estimated time (in days) over which an individual is PCR positive (red bars assumed to take the same value as the 15-19 age group).

### Appendix P: Direct analysis of data

This appendix provides a complementary analysis of the raw data from Appendix F to demonstrate that the estimates of key epidemiological parameters in the paper obtained by ABC-MBP inference are consistent with the data. Note, this analysis relies on some additional simplifying assumptions (most notably ignoring depletion of the susceptible population due to infection acquired immunity and assuming the timescale of the entire epidemic is much longer than for individual disease progression), and so we expect the full analysis in the paper to provide more accurate estimates[33].

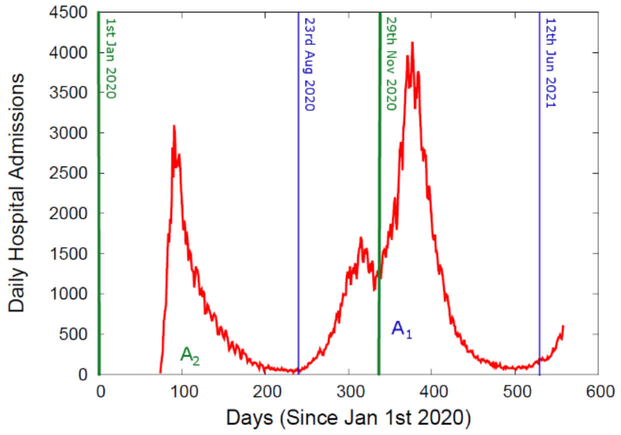


**Figure P2: Hospital admissions**. The red curve shows the total daily COVID-19 hospital admissions for England. The vertical lines denote dates referenced in the text. $A_1$ is the area under the red curve between the two blue lines and $A_2$ is the area between the two green lines.

Based on seroprevalence results from the Coronavirus Infection Survey (CIS) [21] the estimated number of individuals that test antibody positive on 10th Dec 2020 is shown by the blue columns in Fig. P1(a) (note, blood samples were only taken for individuals 16 and over, so no data is available for younger age groups). Because seroconversion takes around 11 days [34], the result in Fig. P1(a) provide an estimate for the overall infected population from the beginning of the pandemic until 29th Nov 2020 (assuming weak waning immunity and perfect antibody test results).

Through random sampling of the population with PCR tests [3], CIS publishes fortnightly estimates giving the total population of PCR positive individuals in England. Data is available and reliable between 23rd Aug 2020 and 12th Jun 2021. Integrating over this time period for each age group gives the distribution shown in Fig. P1(b).

For the next step in the analysis we need to scale Fig. P1(b) such that it becomes representative of the time period from the start of the epidemic (using 1st Jan 2020 as an arbitrary reference point) until 29th Nov 2020 (so coinciding with the antibody CIS results from above). This scaling is achieved by making use of daily COVID-19 hospital admissions data, as shown in Fig. P2, as a proxy for the overall infection level as a function of time (Fig. H1 indicates this is a reasonable assumption because the hospital admissions curve has a profile which closely follows that of the case rate[34]).

The dates 23rd Aug 2020 and 12th Jun 2021 correspond to days 235 and 528 on the $x$-axis in Fig. P2. First we calculate the area $A_1$ under the curve between these time points, then we calculate the area $A_2$ from day 0 until day 333 (29th Nov 2020). Taking the ratio $A_2/A_1$=0.614 provides the factor used to scale Fig. P1(b) to give Fig. P1(c). Hence Fig. P1(c) is representative of the time integrated population of PCR positive individuals in England up to 29th Nov 2020.

---

[33] Additionally, the methodology in the paper provides estimates for $R_t$ as well as having the flexibility to allow for future investigation of time variation in, *e.g.*, age contact factors and vaccination levels.
[34] For simplicity the approximate shift of a few days between the two curves is ignored.

Note, this method of using the hospital admissions curve in Fig. P2 to shift time ranges to coincide with the antibody test results is also used for the case, age-stratified admission and death data below.

We assume that individuals in age group $a$ are PCR positive for time $T_a$. This means that the columns in Fig. P1(a) multiplied by $T_a$ should approximately match the columns in Fig. P1(c) (here we are ignoring "end effects", but these are expected to be relatively small[35]). Conversely, we can use the ratio of the two columns to estimate $T_a$, and this is shown in Fig. P1(d). Given there is no data with which to estimate the three lower age groups, we make the simplifying assumption that the $T_a$ for these groups are equal to the 15-19 age group. This allows us to make an informed guess as to the number of infected individuals in the three lower age groups in Fig. P1(a), as shown by the imputed red bars.

If we compare the graph for PCR time sensitivity in Fig. P1(d) with that from Fig. 5(d) in the main paper (shifted up by 3.8 days to account for the time spent in the infected $\mathrm{I}$ or $\mathrm{A}$ compartments) we find reassuringly good agreement[36].

Next, we look at estimating distributions for branching probabilities. Figure P3(a) shows age-stratified estimates for the total number of cases until 29th Nov 2020 (this is based on data giving the number of individuals with at least one positive PCR test between 8th Aug 2020 and 9th June 2021 [1] scaled by a factor 0.614 using the method from above). Taking the ratio of the cases in Fig. P3(a) with the total infected population in Fig. P1(a) gives an estimate for case branching probability $b_a^{E\to\mathrm{I}}$. Taking one minus this gives $b_a^{E\to\mathrm{A}}$, as shown in Fig. P3(b). We find this to be in excellent agreement with the distribution from the main paper in Fig. 5(a).

Similarly dividing the total hospital admissions [19] in Fig. P3(c) by the total number of cases in Fig. P3(a) yields an estimate for the hospitalisation branching probability $b_a^{I\to\mathrm{C}}$ in Fig. P3(d). Again, this agrees well with Fig. 5(b). Finally, dividing hospitalised death data [2] in Fig. P3(e) with admission data in Fig. P3(c) gives $b_a^{H\to\mathrm{D}}$ in Fig. P3(f), which corresponds well with Fig. 5(c).

[35] By "end effects" we mean discrepancies near to 29th Nov 2020. For example, individuals near to this date do not contributed fully to the integrated sums. However, because the timescale of individual disease progression is small compared to that of the entire epidemic, these end effects can largely be ignored.

[36] Note, this ignores the relatively small ~2% of infected individuals which become hospitalised.

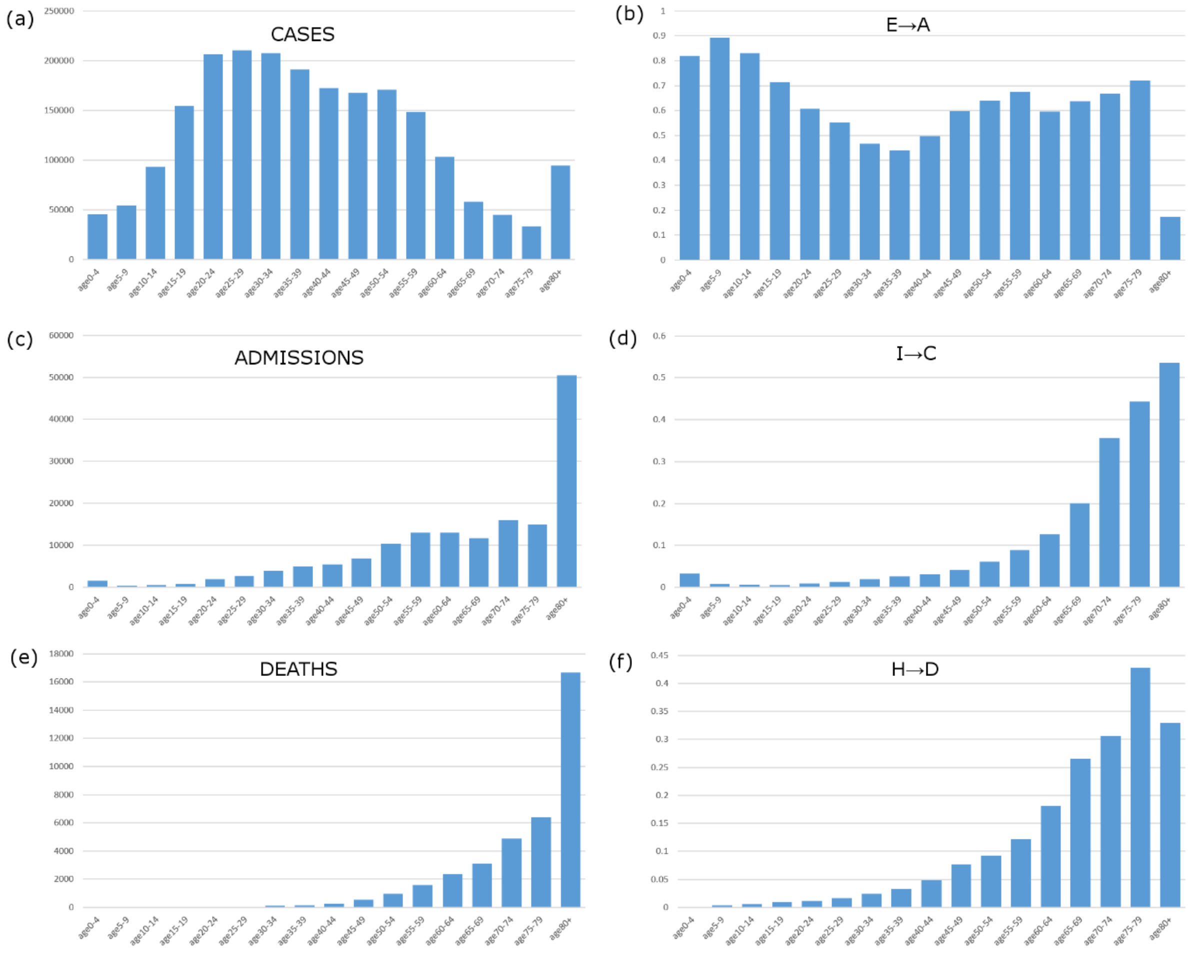


**Figure P3: Branching probability estimates derived directly from the data**. These figures represent estimates for the time period between 1st Jan 2020 and 29th Nov 2020, excluding data for care home residents. (a) The total number of cases. (b) The probability of being asymptomatic. (c) The total number of COVID-19 hospital admissions. (d) The probability of becoming hospitalised given a case. (e) The total number of hospitalised deaths. (f) The probability of death given hospitalised.

Next we look at estimating the age contact factors in Fig. 4(a). As discussed in Appendix A, the next generation matrix (NGM) transforms the age distribution in the infected population in one generation to the next (*e.g*. see Eq. (A8)). The eigenvalue of the NGM gives an estimate for the reproduction number. The corresponding eigenvector gives the expected long-term age distribution of infected individuals[37]. Figure P4(a) shows this eigenvector for the simplest model (using the pre-pandemic contact matrix $\mathbf{C}^0$ and assuming no variation in susceptibility), for which the force of infection is given by

$$\lambda_{a,t} = \beta_t \tfrac{1}{P_a} \sum_{a',c} C^0_{a,a'} \varphi_c N_{c,a',t}. \qquad \text{(R1)}$$

For comparison, Fig. P4(b) shows the normalised[38] total infected population (directly derived from data in Fig. P1(a)). If $\mathbf{C}^0$ correctly captures contacts during the COVID-19 pandemic we would expect

---

[37] This is the distribution the epidemic will converge towards over successive generations. Depletion of the susceptible population due to infection acquired immunity is ignored.

[38] Such that all values add to one, as with the eigenvector.

these two distributions to be the same. The fact that they are different (Fig. P4(b) is clearly higher for younger age groups and has a characteristic bump for older adults) indicates that additional age variation must be added to the model.

As discussed in the paper, alternative interpretations for this can be considered. If we assume that age contact factors $v_a$ modify the pre-pandemic contact rate:

$$\lambda_{a,t} = \beta_t \sigma_a \tfrac{1}{P_a} \sum\nolimits_{a',c} v_a C^0_{a,a'} v_{a'} \varphi_c N_{c,a',t}, \tag{R2}$$

then $v_a$ can be set (ensuring the condition in Eq.(2)) such that Fig. P4(b) is identical to the eigenvector with the largest eigenvalue[39]. This choice of $v_a$ is shown in Fig. P4(c).

On the other hand, if we assume a relative susceptibility $\sigma_a$ for different age groups

$$\lambda_{a,t} = \beta_t \sigma_a \tfrac{1}{P_a} \sum\nolimits_{a',c} C^0_{a,a'} \varphi_c N_{c,a',t}, \tag{R3}$$

then $\sigma_a$ can similarly be chosen to give the correct eigenvector, as shown in Fig. P4(d). Importantly, we find good agreement between the distributions for the age contact factors in Fig. 3(a) and Fig. P4(c) and relative susceptibility in Fig. 3(b) and Fig. P4(d).

As discussed in the main text other possibilities exist, including having an age-dependent infectivity $\varphi_{c,a}$, having an age-dependent residency time in the infective states or even having an age-dependent residency time in the exposed state. Our results indicate that these would be worth exploring once more informative data becomes available.

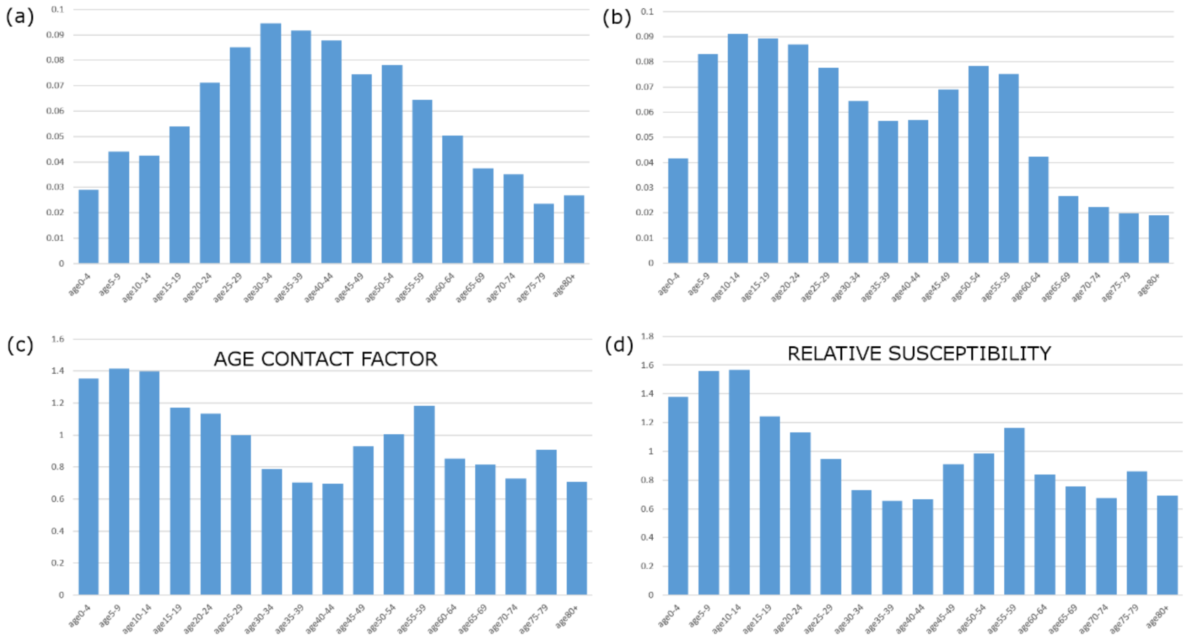


**Figure P4: Derivation of age dependant factors**. (a) The eigenvector of the next generation matrix based on using the pre-pandemic contact matric $\mathbf{C^0}$ (see Eq.(R1)**Error! Reference source not found.**). (b) A normalised distribution giving the total COVID-19 infected population (from the beginning of the pandemic until 29th Nov 2020). (c) Estimated elements of the age contact factor ***v***. (d) Estimated relative susceptibility for different age groups.

[39] This is solved numerically using a gradient decent method.

#### Appendix Q: Variation in susceptibility

Instead of assuming an age-dependent departure from the pre-pandemic contact matrix $\mathbf{C^0}$, this appendix looks at an alternative model in which there is age-stratification in the susceptibility of individuals. In other words, instead of Eq.(1) the force of infection is given by:

$$\lambda_{a,t} = \sigma_a r_t R_t \left[ \tfrac{1}{P_a} \sum_{a',c} C^0_{a,a'} \varphi_c N_{c,a',t} \right] + \tfrac{1}{f} \eta_t . \tag{R4}$$

where $\sigma_a$ is the relative susceptibility for age group $a$. As with the age contact factors, the values for $\sigma_a$ are constrained to have a population weighted average of one:

$$\sum_a \sigma_a \tfrac{P_a}{P} = 1, \tag{R5}$$

Inference was re-run using this new model and the results are shown in Fig. Q1. Of particular importance is the age-stratified distribution for the relative susceptibility $\sigma_a$ shown in Fig. Q1(b). Unsurprisingly, it exhibits much the same variation with age as the age contact factors in Fig. 3(a).

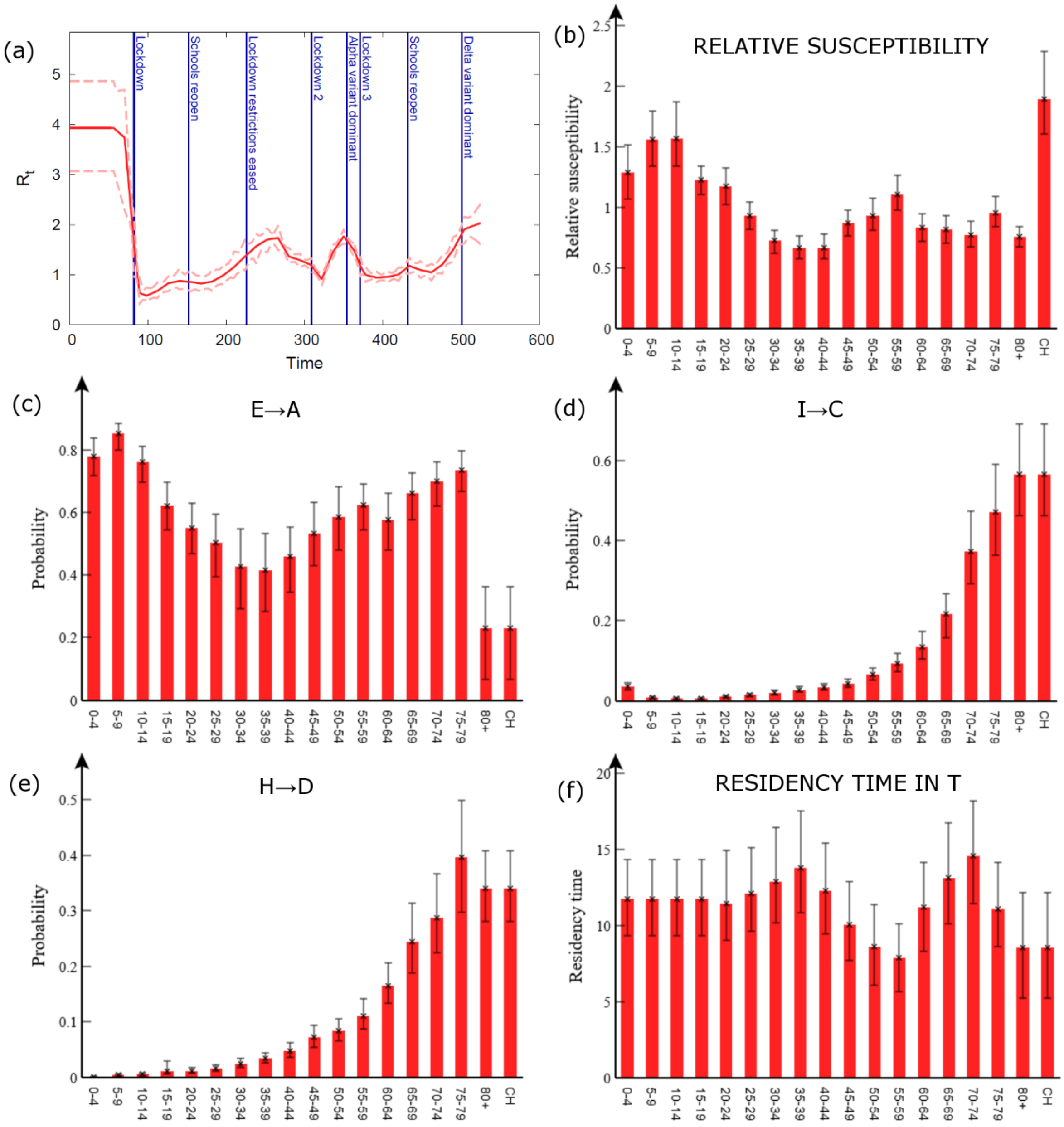


**Figure Q1: Age-stratified susceptibility.** This shows the results of inference using a model with a pre-pandemic contact matrix $\mathbf{C^0}$ and an age-stratified susceptibility (see Eq.(7)**Error! Reference source not found.**). (a) The reproduction number $R_t$ (the red solid line gives the posterior mean and the dashed lines denote 95% credible intervals). For (b) the age contact factor, (c) the asymptomatic branching probability, (d) the hospitalised branching probability, (e) the death branching probability, and (f) the residency time in the T compartment, the red columns with error bars give the posterior means with 95% credible intervals.
