## Supplementary Results for "Estimation of age-stratified contact rates during the COVID-19 pandemic using a novel inference algorithm"

### Posterior Supplementary Results for the paper:

### Summary

This document gives further details regarding the posterior distributions generated by the ABC-MBP inference algorithm using COVID-19 data from England, as described in the section "Data Sources". Section 1 provides a table giving the priors and posterior distributions for model parameters, along with a table for the estimated age-adjusted contact matrix **C**. Section 2 gives graphs showing the posterior fit to the data. Finally, section 3 shows how posterior parameter estimates change with generation number when ABC-MBP is run.

### 1) Parameter estimates

| Param. | Prior | Posterior Mean | Posterior 95% CI | Description |
|---|---|---|---|---|
| $m_E$ | Fixed(3.8) | 3.8 | 3.8 — 3.8 | E mean residency time. |
| $m_I$ | Fixed(4) | 4.0 | 4.0 — 4.0 | I mean residency time. |
| $m_A$ | Fixed(4) | 4.0 | 4.0 — 4.0 | A mean residency time. |
| $m_C$ | Fixed(3.1) | 3.1 | 3.1 — 3.1 | C mean residency time. |
| $m_H$ | Fixed(13) | 13.0 | 13.0 — 13.0 | H residency time. |
| $m_{T,0\text{-}19}$ | Uniform(4,20) | 11.4 | 9.34 — 13.7 | T residency time 0-19. |
| $m_{T,20\text{-}24}$ | Uniform(4,20) | 11.0 | 8.26 — 14.1 | T residency time 20-24. |
| $m_{T,25\text{-}29}$ | Uniform(4,20) | 11.6 | 8.82 — 14.7 | T residency time 25-29. |
| $m_{T,30\text{-}34}$ | Uniform(4,20) | 12.4 | 9.26 — 17.1 | T residency time 30-34. |
| $m_{T,35\text{-}39}$ | Uniform(4,20) | 13.4 | 10.3 — 17.2 | T residency time 35-39. |
| $m_{T,40\text{-}44}$ | Uniform(4,20) | 12.2 | 9.58 — 15.6 | T residency time 40-44. |
| $m_{T,45\text{-}49}$ | Uniform(4,20) | 10.0 | 7.64 — 12.5 | T residency time 45-49. |
| $m_{T,50\text{-}54}$ | Uniform(4,20) | 8.7 | 6.0 — 11.9 | T residency time 50-54. |
| $m_{T,55\text{-}59}$ | Uniform(4,20) | 8.1 | 5.8 — 10.6 | T residency time 55-59. |
| $m_{T,60\text{-}64}$ | Uniform(4,20) | 11.4 | 8.6 — 14.5 | T residency time 60-64. |
| $m_{T,65\text{-}69}$ | Uniform(4,20) | 13.3 | 10.3 — 16.7 | T residency time 65-69. |
| $m_{T,70\text{-}74}$ | Uniform(4,20) | 14.6 | 11.1 — 18.7 | T residency time 70-74. |
| $m_{T,75\text{-}79}$ | Uniform(4,20) | 11.4 | 8.4 — 14.6 | T residency time 75-79. |
| $m_{T,80+}$ | Uniform(4,20) | 9.0 | 5.7 — 12.4 | T residency time 80+. |
| $m_{T,CH}$ | Set to $m_{T,80+}$ | | | T residency time CH. |
| $\varphi_I$ | Fixed(1) | 1.0 --- 1.0 | 1.0 — 1.0 | Relative infectivity of I. |
| $\varphi_A$ | Fixed(0.55) | 0.55 | 0.55 — 0.55 | Relative infectivity of A. |
| $b_{0-4}^{E\to A}$ | Dir(1) | 0.779 | 0.708 — 0.831 | Asymptomatic BP 0-4. |
| $b_{5-9}^{E\to A}$ | Dir(1) | 0.853 | 0.813 — 0.891 | Asymptomatic BP 5-9. |
| $b_{10-14}^{E\to A}$ | Dir(1) | 0.758 | 0.688 — 0.817 | Asymptomatic BP 10-14. |
| $b_{15-19}^{E\to A}$ | Dir(1) | 0.636 | 0.553 — 0.722 | Asymptomatic BP 15-19. |
| $b_{20-24}^{E\to A}$ | Dir(1) | 0.555 | 0.418 — 0.638 | Asymptomatic BP 20-24. |
| $b_{25-29}^{E\to A}$ | Dir(1) | 0.529 | 0.429 — 0.613 | Asymptomatic BP 25-29. |
| $b_{30-34}^{E\to A}$ | Dir(1) | 0.458 | 0.309 — 0.592 | Asymptomatic BP 30-34. |
| $b_{35-39}^{E\to A}$ | Dir(1) | 0.430 | 0.271 — 0.550 | Asymptomatic BP 35-39. |
| $b_{40-44}^{E\to A}$ | Dir(1) | 0.461 | 0.328 — 0.600 | Asymptomatic BP 40-44. |
| $b_{45-49}^{E\to A}$ | Dir(1) | 0.533 | 0.400 — 0.637 | Asymptomatic BP 45-49. |
| $b_{50-54}^{E\to A}$ | Dir(1) | 0.577 | 0.468 — 0.682 | Asymptomatic BP 50-54. |
| $b_{55-59}^{E\to A}$ | Dir(1) | 0.620 | 0.533 — 0.691 | Asymptomatic BP 55-59. |
| $b_{60-64}^{E\to A}$ | Dir(1) | 0.572 | 0.472 — 0.661 | Asymptomatic BP 60-64. |
| $b_{65-69}^{E\to A}$ | Dir(1) | 0.649 | 0.553 — 0.727 | Asymptomatic BP 65-69. |
| $b_{70-74}^{E\to A}$ | Dir(1) | 0.699 | 0.605 — 0.763 | Asymptomatic BP 70-74. |
| $b_{75-79}^{E\to A}$ | Dir(1) | 0.731 | 0.663 — 0.787 | Asymptomatic BP 75-79. |
| $b_{80+}^{E\to A}$ | Dir(1) | 0.231 | 0.109 — 0.341 | Asymptomatic BP 80+. |
| $b_{CH}^{E\to A}$ | Set to $b_{80+}^{E\to A}$ | | | Asymptomatic BP CH. |
| $b_{0-4}^{I\to C}$ | Dir(0.3) | 0.036 | 0.020 — 0.047 | Hospitalised BP 0-4. |
| $b_{5-9}^{I\to C}$ | Dir(0.3) | 0.007 | 0.005 — 0.011 | Hospitalised BP 5-9. |
| $b_{10-14}^{I\to C}$ | Dir(0.3) | 0.006 | 0.004 — 0.008 | Hospitalised BP 10-14. |
| $b_{15-19}^{I\to C}$ | Dir(0.3) | 0.006 | 0.004 — 0.008 | Hospitalised BP 15-19. |
| $b_{20-24}^{I\to C}$ | Dir(0.3) | 0.010 | 0.005 — 0.013 | Hospitalised BP 20-24. |
| $b_{25-29}^{I\to C}$ | Dir(0.3) | 0.015 | 0.011 — 0.019 | Hospitalised BP 25-29. |
| $b_{30-34}^{I\to C}$ | Dir(0.3) | 0.021 | 0.016 — 0.027 | Hospitalised BP 30-34. |
| $b_{35-39}^{I\to C}$ | Dir(0.3) | 0.027 | 0.020 — 0.035 | Hospitalised BP 35-39. |

| | | | | |
|---|---|---|---|---|
| $b_{40-44}^{I\to C}$ | Dir(0.3) | 0.034 | 0.026 — 0.045 | Hospitalised BP 40-44. |
| $b_{45-49}^{I\to C}$ | Dir(0.3) | 0.044 | 0.033 — 0.056 | Hospitalised BP 45-49. |
| $b_{50-54}^{I\to C}$ | Dir(0.3) | 0.065 | 0.050 — 0.083 | Hospitalised BP 50-54. |
| $b_{55-59}^{I\to C}$ | Dir(0.3) | 0.095 | 0.079 — 0.120 | Hospitalised BP 55-59. |
| $b_{60-64}^{I\to C}$ | Dir(0.3) | 0.136 | 0.105 — 0.175 | Hospitalised BP 60-64. |
| $b_{65-69}^{I\to C}$ | Dir(0.4) | 0.208 | 0.160 — 0.273 | Hospitalised BP 65-69. |
| $b_{70-74}^{I\to C}$ | Dir(0.6) | 0.374 | 0.282 — 0.481 | Hospitalised BP 70-74. |
| $b_{75-79}^{I\to C}$ | Dir(0.8) | 0.472 | 0.360 — 0.590 | Hospitalised BP 75-79. |
| $b_{80+}^{I\to C}$ | Dir(1) | 0.586 | 0.464 — 0.741 | Hospitalised BP 80+. |
| $b_{CH}^{I\to C}$ | Set to $b_{80+}^{I\to C}$ | | | Care home BP CH. |
| $b_{0-4}^{H\to D}$ | Dir(0.3) | 0.0008 | 0.0002 — 0.0062 | Death BP 0-4. |
| $b_{5-9}^{H\to D}$ | Dir(0.3) | 0.0035 | 0.0019 — 0.0058 | Death BP 5-9. |
| $b_{10-14}^{H\to D}$ | Dir(0.3) | 0.0054 | 0.0029 — 0.0089 | Death BP 10-14. |
| $b_{15-19}^{H\to D}$ | Dir(0.3) | 0.0085 | 0.0049 — 0.0138 | Death BP 15-19. |
| $b_{20-24}^{H\to D}$ | Dir(0.3) | 0.012 | 0.007 — 0.026 | Death BP 20-24. |
| $b_{25-29}^{H\to D}$ | Dir(0.3) | 0.017 | 0.011 — 0.024 | Death BP 25-29. |
| $b_{30-34}^{H\to D}$ | Dir(0.3) | 0.024 | 0.017 — 0.034 | Death BP 30-34. |
| $b_{35-39}^{H\to D}$ | Dir(0.3) | 0.033 | 0.024 — 0.046 | Death BP 35-39. |
| $b_{40-44}^{H\to D}$ | Dir(0.3) | 0.046 | 0.033 — 0.061 | Death BP 40-44. |
| $b_{45-49}^{H\to D}$ | Dir(0.3) | 0.071 | 0.053 — 0.093 | Death BP 45-49. |
| $b_{50-54}^{H\to D}$ | Dir(0.3) | 0.086 | 0.066 — 0.113 | Death BP 50-54. |
| $b_{55-59}^{H\to D}$ | Dir(0.3) | 0.110 | 0.083 — 0.144 | Death BP 55-59. |
| $b_{60-64}^{H\to D}$ | Dir(0.3) | 0.165 | 0.124 — 0.213 | Death BP 60-64. |
| $b_{65-69}^{H\to D}$ | Dir(0.4) | 0.251 | 0.188 — 0.311 | Death BP 65-69. |
| $b_{70-74}^{H\to D}$ | Dir(0.6) | 0.290 | 0.222 — 0.381 | Death BP 70-74. |
| $b_{75-79}^{H\to D}$ | Dir(0.8) | 0.398 | 0.308 — 0.493 | Death BP 75-79. |
| $b_{80+}^{H\to D}$ | Dir(1) | 0.334 | 0.262 — 0.417 | Death BP 80+. |
| $b_{CH}^{H\to D}$ | Set to $b_{80+}^{H\to D}$ | | | Death BP CH. |
| $R_0$ | Uniform(0.4,5) | 4.03 | 3.13 — 4.94 | Rep. num. until 26/02/2020. |
| $R_1$ | Uniform(0.4,5) | 3.52 | 2.66 — 4.75 | Rep. number 11/03/2020. |
| $R_2$ | Uniform(0.4,5) | 1.67 | 1.13 — 2.37 | Rep. number 23/03/2020. |
| $R_3$ | Uniform(0.4,5) | 0.67 | 0.43 — 1.01 | Rep. number 30/03/2020. |
| $R_4$ | Uniform(0.4,2.5) | 0.58 | 0.43 — 0.73 | Rep. number 08/04/2020. |
| $R_5$ | Uniform(0.4,2.5) | 0.69 | 0.54 — 0.85 | Rep. number 22/04/2020. |
| $R_6$ | Uniform(0.4,2.5) | 0.84 | 0.64 — 1.04 | Rep. number 06/05/2020. |
| $R_7$ | Uniform(0.4,2.5) | 0.89 | 0.72 — 1.07 | Rep. number 20/05/2020. |
| $R_8$ | Uniform(0.4,2.5) | 0.85 | 0.68 — 1.06 | Rep. number 03/06/2020. |
| $R_9$ | Uniform(0.4,2.5) | 0.85 | 0.67 — 1.07 | Rep. number 17/06/2020. |
| $R_{10}$ | Uniform(0.4,2.5) | 0.89 | 0.65 — 1.11 | Rep. number 01/07/2020. |
| $R_{11}$ | Uniform(0.4,2.5) | 1.03 | 0.79 — 1.27 | Rep. number 15/07/2020. |
| $R_{12}$ | Uniform(0.4,2.5) | 1.23 | 0.95 — 1.53 | Rep. number 29/07/2020. |
| $R_{13}$ | Uniform(0.4,2.5) | 1.45 | 1.14 — 1.83 | Rep. number 12/08/2020. |
| $R_{14}$ | Uniform(0.4,2.5) | 1.62 | 1.38 — 1.90 | Rep. number 26/08/2020. |
| $R_{15}$ | Uniform(0.4,2.5) | 1.74 | 1.48 — 2.06 | Rep. number 09/09/2020. |
| $R_{16}$ | Uniform(0.4,2.5) | 1.77 | 1.52 — 2.00 | Rep. number 23/09/2020. |
| $R_{17}$ | Uniform(0.4,2.5) | 1.40 | 1.24 — 1.57 | Rep. number 07/10/2020. |
| $R_{18}$ | Uniform(0.4,2.5) | 1.34 | 1.19 — 1.55 | Rep. number 21/10/2020. |
| $R_{19}$ | Uniform(0.4,2.5) | 1.23 | 1.03 — 1.38 | Rep. number 04/11/2020. |
| $R_{20}$ | Uniform(0.4,2.5) | 0.97 | 0.82 — 1.11 | Rep. number 18/11/2020. |
| $R_{21}$ | Uniform(0.4,2.5) | 1.46 | 1.27 — 1.66 | Rep. number 02/12/2020. |

| | | | | |
|---|---|---|---|---|
| $R_{22}$ | Uniform(0.4,2.5) | 1.86 | 1.69 — 2.03 | Rep. number 16/12/2020. |
| $R_{23}$ | Uniform(0.4,2.5) | 1.54 | 1.31 — 1.76 | Rep. number 30/12/2020. |
| $R_{24}$ | Uniform(0.4,2.5) | 1.22 | 1.02 — 1.46 | Rep. number 06/01/2021. |
| $R_{25}$ | Uniform(0.4,2.5) | 1.03 | 0.88 — 1.21 | Rep. number 13/01/2021. |
| $R_{26}$ | Uniform(0.4,2.5) | 0.99 | 0.85 — 1.13 | Rep. number 27/01/2021. |
| $R_{27}$ | Uniform(0.4,2.5) | 1.01 | 0.88 — 1.16 | Rep. number 10/02/2021. |
| $R_{28}$ | Uniform(0.4,2.5) | 1.06 | 0.93 — 1.21 | Rep. number 24/02/2021. |
| $R_{29}$ | Uniform(0.4,2.5) | 1.23 | 1.04 — 1.43 | Rep. number 10/03/2021. |
| $R_{30}$ | Uniform(0.4,2.5) | 1.14 | 0.95 — 1.33 | Rep. number 24/03/2021. |
| $R_{31}$ | Uniform(0.4,2.5) | 1.10 | 0.93 — 1.27 | Rep. number 07/04/2021. |
| $R_{32}$ | Uniform(0.4,2.5) | 1.28 | 1.07 — 1.55 | Rep. number 21/04/2021. |
| $R_{33}$ | Uniform(0.4,2.5) | 1.57 | 1.33 — 1.83 | Rep. number 05/05/2021. |
| $R_{34}$ | Uniform(0.4,2.5) | 1.99 | 1.71 — 2.32 | Rep. number 19/05/2021. |
| $R_{35}$ | Uniform(0.4,2.5) | 2.16 | 1.76 — 2.45 | Rep. number 09/06/2021. |
| $v_{0\text{-}4}$ | MDir(0.5) | 1.29 | 1.09 — 1.47 | Age contact factor 0-4. |
| $v_{5\text{-}9}$ | MDir(0.5) | 1.43 | 1.28 — 1.57 | Age contact factor 5-9. |
| $v_{10\text{-}14}$ | MDir(0.5) | 1.38 | 1.23 — 1.53 | Age contact factor 10-14. |
| $v_{15\text{-}19}$ | MDir(0.5) | 1.16 | 1.08 — 1.23 | Age contact factor 15-19. |
| $v_{20\text{-}24}$ | MDir(0.5) | 1.16 | 1.05 — 1.28 | Age contact factor 20-24. |
| $v_{25\text{-}29}$ | MDir(0.5) | 1.00 | 0.90 — 1.12 | Age contact factor 25-29. |
| $v_{30\text{-}34}$ | MDir(0.5) | 0.80 | 0.70 — 0.94 | Age contact factor 30-34. |
| $v_{35\text{-}39}$ | MDir(0.5) | 0.73 | 0.63 — 0.84 | Age contact factor 35-39. |
| $v_{40\text{-}44}$ | MDir(0.5) | 0.70 | 0.60 — 0.83 | Age contact factor 40-44. |
| $v_{45\text{-}49}$ | MDir(0.5) | 0.88 | 0.77— 1.01 | Age contact factor 45-49. |
| $v_{50\text{-}54}$ | MDir(0.5) | 0.94 | 0.81 — 1.10 | Age contact factor 50-54. |
| $v_{50\text{-}59}$ | MDir(0.5) | 1.12 | 0.99 — 1.25 | Age contact factor 55-59. |
| $v_{60\text{-}64}$ | MDir(0.5) | 0.84 | 0.76 — 0.97 | Age contact factor 60-64. |
| $v_{65\text{-}69}$ | MDir(0.5) | 0.86 | 0.77 — 0.98 | Age contact factor 65-69. |
| $v_{70\text{-}74}$ | MDir(0.5) | 0.83 | 0.72 — 0.95 | Age contact factor 70-74. |
| $v_{75\text{-}79}$ | MDir(0.5) | 0.99 | 0.87 — 1.13 | Age contact factor 75-79. |
| $v_{80+}$ | MDir(0.5) | 0.77 | 0.68 — 0.86 | Age contact factor 80+. |
| $v_{CH}$ | MDir(0.5) | 1.54 | 1.40 — 1.67 | Age contact factor CH. |

**Table R1: Inferred parameter estimates**. This table provides inferred distributions (giving posterior means and 95% credible intervals) for all model parameters. Residency times are given in days. “Rep. number” is here short for “reproduction number” (note, this isn’t the effective reproduction number) and “BP” stands for branching probability. The branching probabilities leaving a given compartment must add up to one, and so by definition $b_a^{E\to \mathrm{I}} = 1 - b_a^{E\to \mathrm{A}}$ and $b_a^{I\to \mathrm{T}} = 1 - b_a^{I\to \mathrm{C}}$. A Dirichlet prior is used on these branching probabilities and “Dir(α)” specifies this for a given value of α. A modified Dirichlet prior denoted “MDir($\sigma$)” is placed on the age contact factors (see Appendix J for details).

| **Age** | **0-4** | **5-9** | **10-14** | **15-19** | **20-24** | **25-29** | **30-34** | **35-39** | **40-44** |
|---|---|---|---|---|---|---|---|---|---|
| **0-4** | 1.48 | 1.12 | 0.13 | 0.11 | 0.15 | 0.29 | 0.54 | 0.56 | 0.28 |
| **5-9** | 1.22 | 4.46 | 0.51 | 0.29 | 0.30 | 0.35 | 0.42 | 0.73 | 0.84 |
| **10-14** | 0.14 | 0.50 | 4.69 | 1.45 | 0.30 | 0.31 | 0.34 | 0.33 | 0.60 |
| **15-19** | 0.10 | 0.26 | 1.32 | 5.33 | 1.06 | 0.36 | 0.30 | 0.26 | 0.32 |
| **20-24** | 0.16 | 0.30 | 0.31 | 1.19 | 3.79 | 1.42 | 0.70 | 0.47 | 0.44 |
| **25-29** | 0.34 | 0.38 | 0.35 | 0.44 | 1.55 | 2.22 | 1.16 | 0.70 | 0.59 |
| **30-34** | 0.64 | 0.46 | 0.38 | 0.37 | 0.77 | 1.17 | 1.62 | 0.77 | 0.58 |
| **35-39** | 0.65 | 0.77 | 0.36 | 0.31 | 0.50 | 0.69 | 0.75 | 1.28 | 0.68 |
| **40-44** | 0.30 | 0.83 | 0.61 | 0.35 | 0.44 | 0.54 | 0.53 | 0.63 | 1.15 |
| **45-49** | 0.18 | 0.66 | 0.88 | 0.66 | 0.57 | 0.58 | 0.52 | 0.54 | 0.67 |
| **50-54** | 0.13 | 0.42 | 0.74 | 0.74 | 0.77 | 0.68 | 0.57 | 0.56 | 0.64 |
| **55-59** | 0.18 | 0.35 | 0.33 | 0.50 | 0.72 | 0.76 | 0.58 | 0.51 | 0.55 |
| **60-64** | 0.13 | 0.34 | 0.16 | 0.20 | 0.35 | 0.38 | 0.34 | 0.30 | 0.28 |
| **65-69** | 0.14 | 0.17 | 0.11 | 0.09 | 0.22 | 0.25 | 0.24 | 0.24 | 0.23 |
| **70-74** | 0.14 | 0.16 | 0.15 | 0.08 | 0.08 | 0.16 | 0.18 | 0.21 | 0.23 |
| **75-79** | 0.12 | 0.14 | 0.13 | 0.11 | 0.06 | 0.05 | 0.10 | 0.14 | 0.18 |
| **80+** | 0.11 | 0.13 | 0.13 | 0.11 | 0.10 | 0.05 | 0.04 | 0.09 | 0.14 |
| **CH** | 0.05 | 0.05 | 0.05 | 0.04 | 0.04 | 0.09 | 0.04 | 0.04 | 0.11 |

| **Age** | **45-49** | **50-54** | **55-59** | **60-64** | **65-69** | **70-74** | **75-79** | **80+** | **CH** |
|---|---|---|---|---|---|---|---|---|---|
| **0-4** | 0.16 | 0.11 | 0.16 | 0.13 | 0.16 | 0.16 | 0.19 | 0.14 | 0.35 |
| **5-9** | 0.64 | 0.37 | 0.32 | 0.36 | 0.21 | 0.20 | 0.24 | 0.18 | 0.45 |
| **10-14** | 0.83 | 0.65 | 0.30 | 0.16 | 0.13 | 0.19 | 0.23 | 0.17 | 0.42 |
| **15-19** | 0.56 | 0.59 | 0.41 | 0.19 | 0.10 | 0.09 | 0.18 | 0.13 | 0.33 |
| **20-24** | 0.54 | 0.68 | 0.66 | 0.37 | 0.27 | 0.10 | 0.11 | 0.13 | 0.33 |
| **25-29** | 0.60 | 0.66 | 0.75 | 0.44 | 0.33 | 0.21 | 0.10 | 0.07 | 0.80 |
| **30-34** | 0.54 | 0.56 | 0.59 | 0.40 | 0.32 | 0.24 | 0.20 | 0.06 | 0.39 |
| **35-39** | 0.55 | 0.53 | 0.50 | 0.34 | 0.32 | 0.27 | 0.26 | 0.13 | 0.38 |
| **40-44** | 0.63 | 0.57 | 0.50 | 0.30 | 0.28 | 0.29 | 0.31 | 0.19 | 0.94 |
| **45-49** | 1.03 | 0.75 | 0.63 | 0.34 | 0.31 | 0.34 | 0.45 | 0.30 | 1.83 |
| **50-54** | 0.80 | 1.18 | 0.95 | 0.54 | 0.42 | 0.32 | 0.44 | 0.36 | 2.43 |
| **55-59** | 0.66 | 0.93 | 1.17 | 0.77 | 0.53 | 0.48 | 0.47 | 0.40 | 3.23 |
| **60-64** | 0.31 | 0.45 | 0.66 | 0.76 | 0.51 | 0.35 | 0.42 | 0.25 | 2.17 |
| **65-69** | 0.24 | 0.30 | 0.39 | 0.44 | 0.83 | 0.49 | 0.45 | 0.33 | 0.66 |
| **70-74** | 0.27 | 0.23 | 0.36 | 0.31 | 0.49 | 0.78 | 0.56 | 0.32 | 0.78 |
| **75-79** | 0.24 | 0.22 | 0.24 | 0.25 | 0.31 | 0.39 | 0.94 | 0.50 | 0.94 |
| **80+** | 0.21 | 0.23 | 0.27 | 0.20 | 0.30 | 0.29 | 0.65 | 0.70 | 0.92 |
| **CH** | 0.21 | 0.26 | 0.36 | 0.28 | 0.10 | 0.12 | 0.20 | 0.15 | 5.20 |

**Table R2: Inferred age-adjusted contact matrix C.** For an individual in age group given by the column heading, this table provides an posterior mean estimate proportional to the average daily contacts they make with individuals in the same and different age groups (shown down each row). Note, whilst **C** is an estimated constant matrix, the overall rate of effective contacts is modulated by the reproduction number $R_t$, as shown in Eq.(3).

### 2) State graphs

Below are a series of graphs that show the fit between the inferred posterior states and the data:

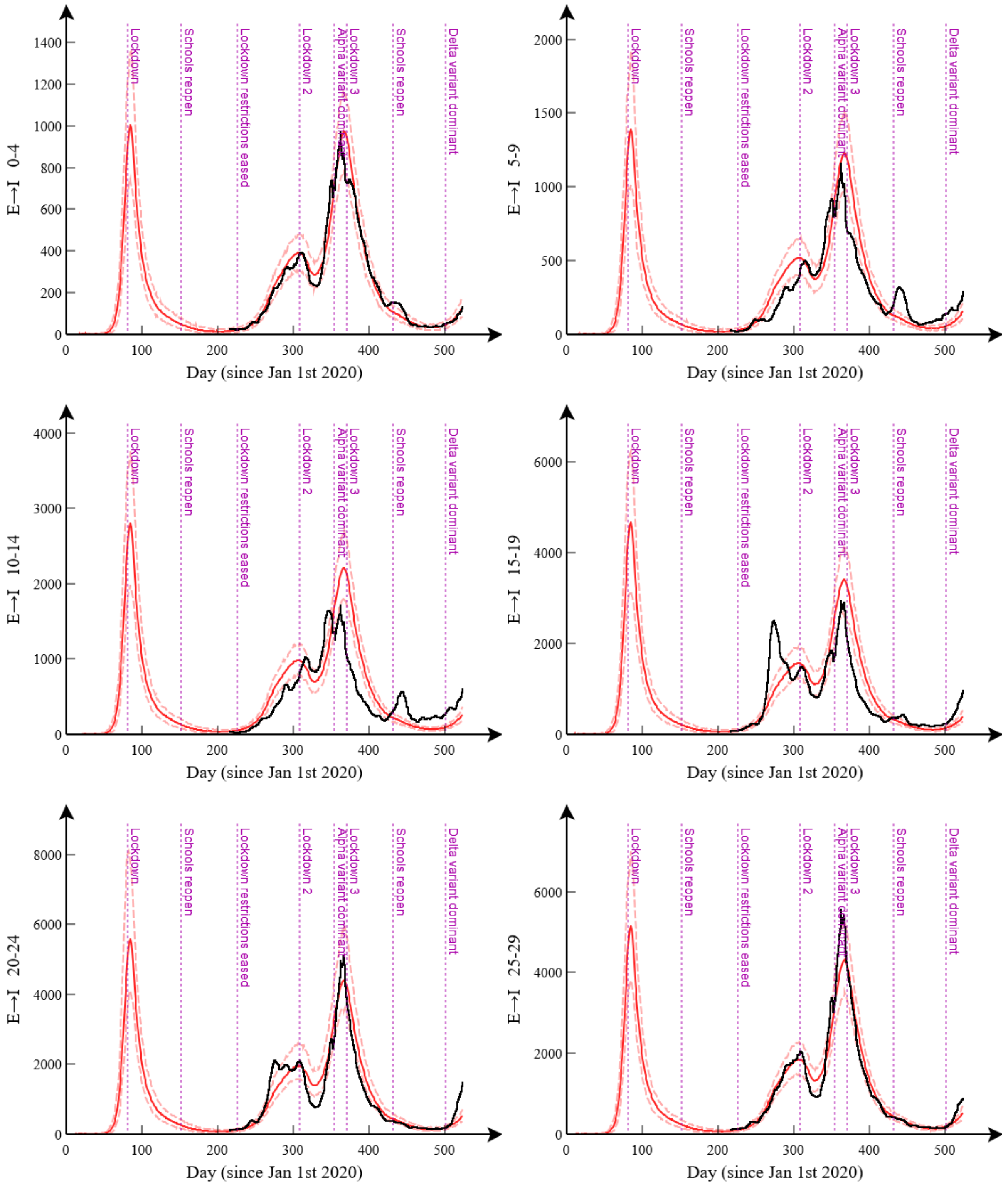


**Figure R1(a): Case data ages 0-29**. This shows the posterior distribution for the daily number of exposed to infectious transitions (red solid line gives the posterior mean and the dashed lines denote 95% credible intervals) and the case data (black line), shifted 4 days to account for the time between becoming infectious and getting tested. Each graph represents a different age group.

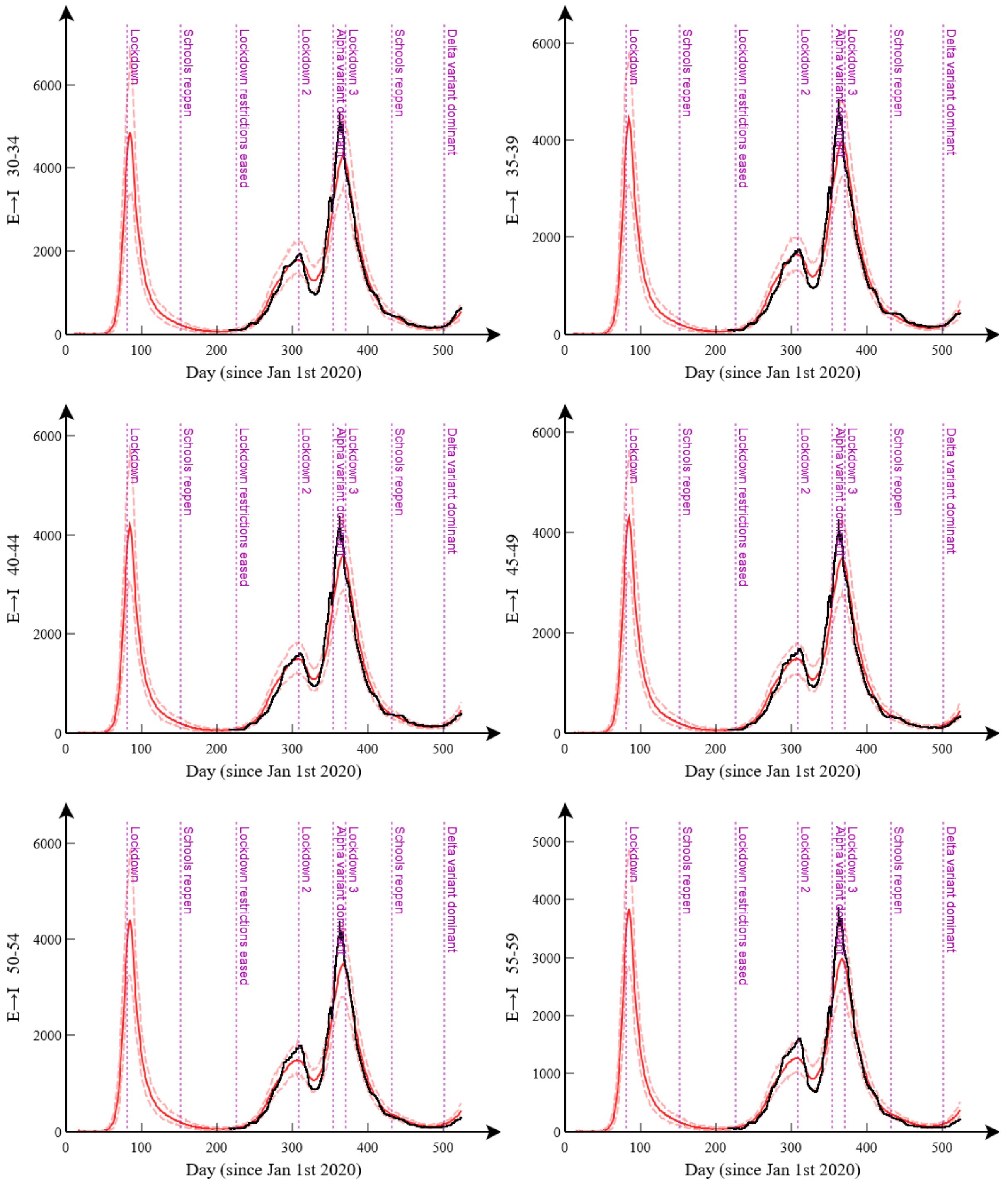


**Figure R1(b): Case data ages 30-59**. This shows the posterior distribution for the daily number of exposed to infectious transitions (red solid line gives the posterior mean and the dashed lines denote 95% credible intervals) and the case data (black line), shifted 4 days to account for the time between becoming infectious and getting tested. Each graph represents a different age group.

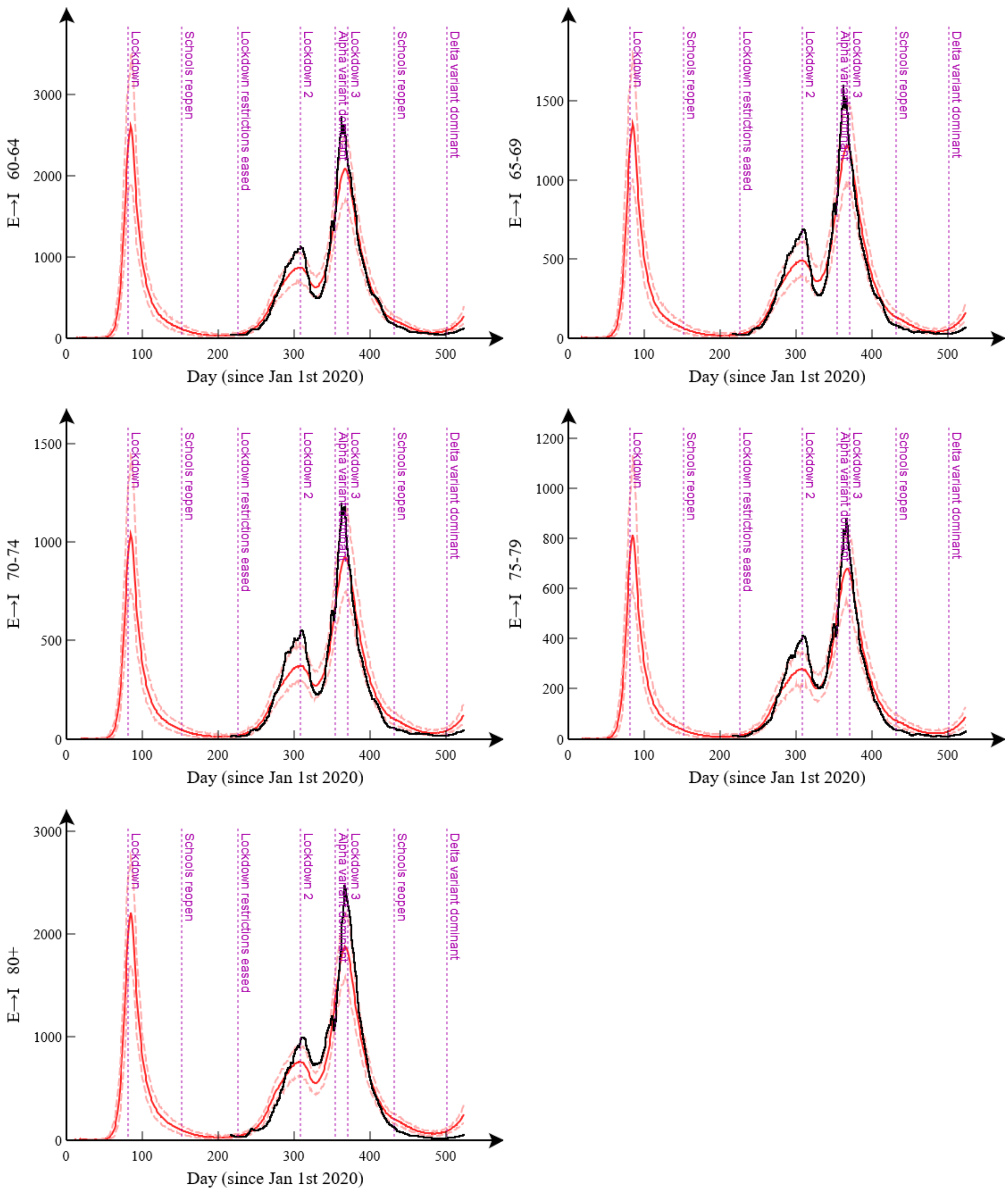


**Figure R1(c): Case data ages 60-80+**. This shows the posterior distribution for the daily number of exposed to infectious transitions (red solid line gives the posterior mean and the dashed lines denote 95% credible intervals) and the case data (black line), shifted 4 days to account for the time between becoming infectious and getting tested. Each graph represents a different age group.

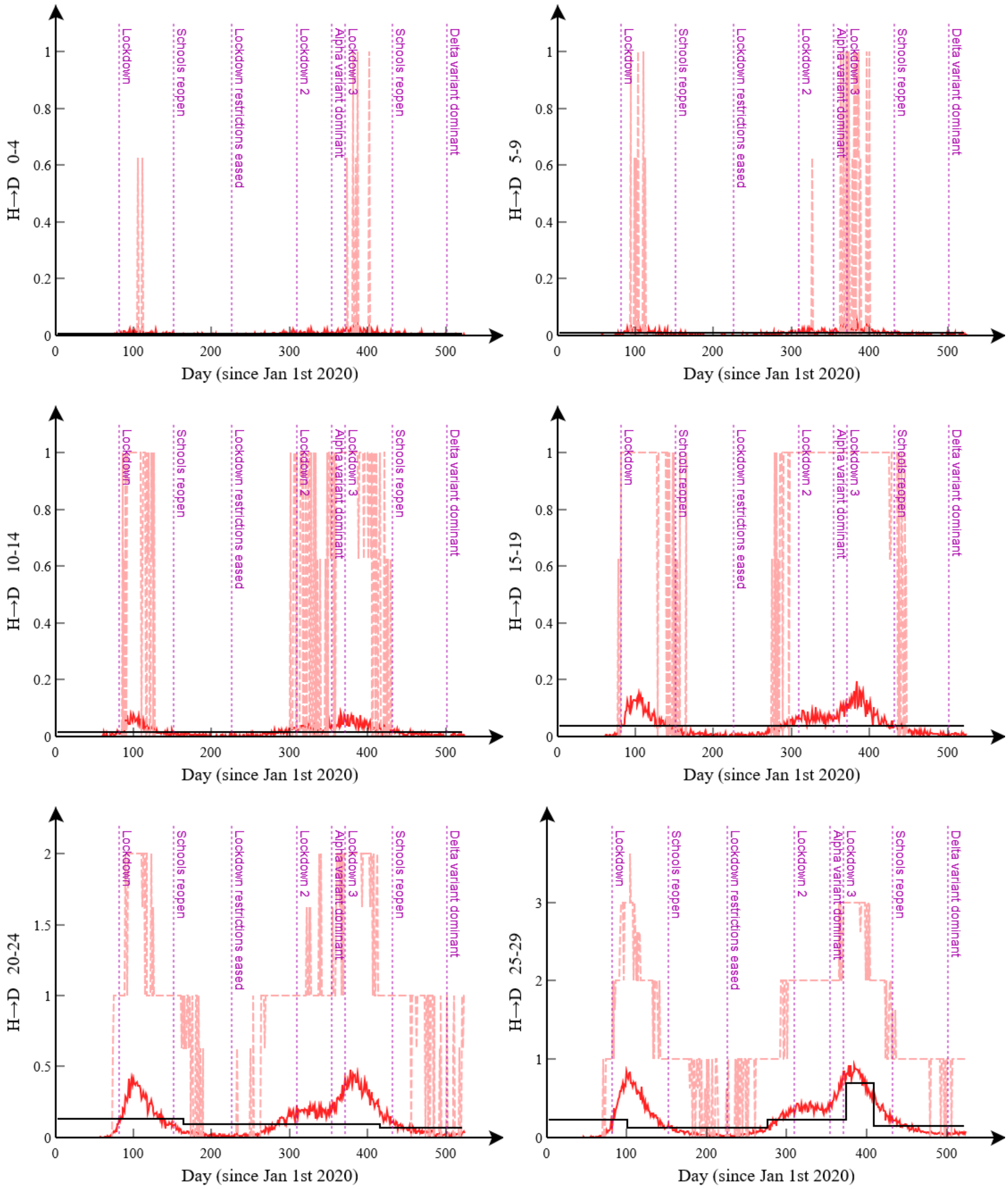


**Figure R1(d): Death data ages 0-29**. This shows the posterior distribution for the inferred daily number of deaths (red solid line gives the posterior mean and the dashed lines denote 95% credible intervals) and the actual data (black line). Each graph represents a different age group. Note, when the death rate is low the data are amalgamated into longer time windows (to avoid Poisson noise). Note, noise in the credible intervals when the transition numbers are low is a finite particle number effect.

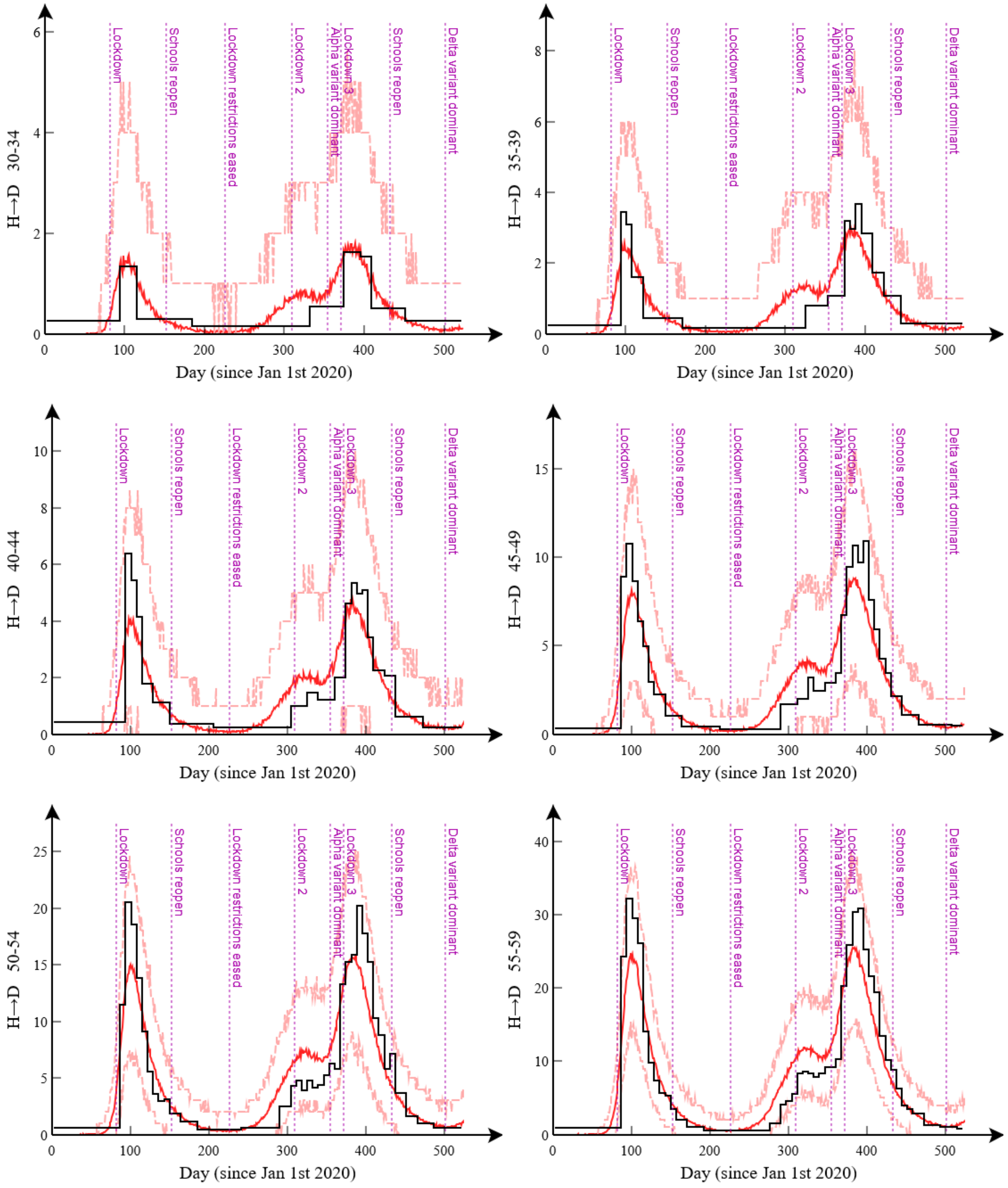


**Figure R1(e): Death data ages 30-59**. This shows the posterior distribution for the inferred daily number of deaths (red solid line gives the posterior mean and the dashed lines denote 95% credible intervals) and the actual data (black line). Each graph represents a different age group. Note, when the death rate is low the data are amalgamated into longer time windows (to avoid Poisson noise).

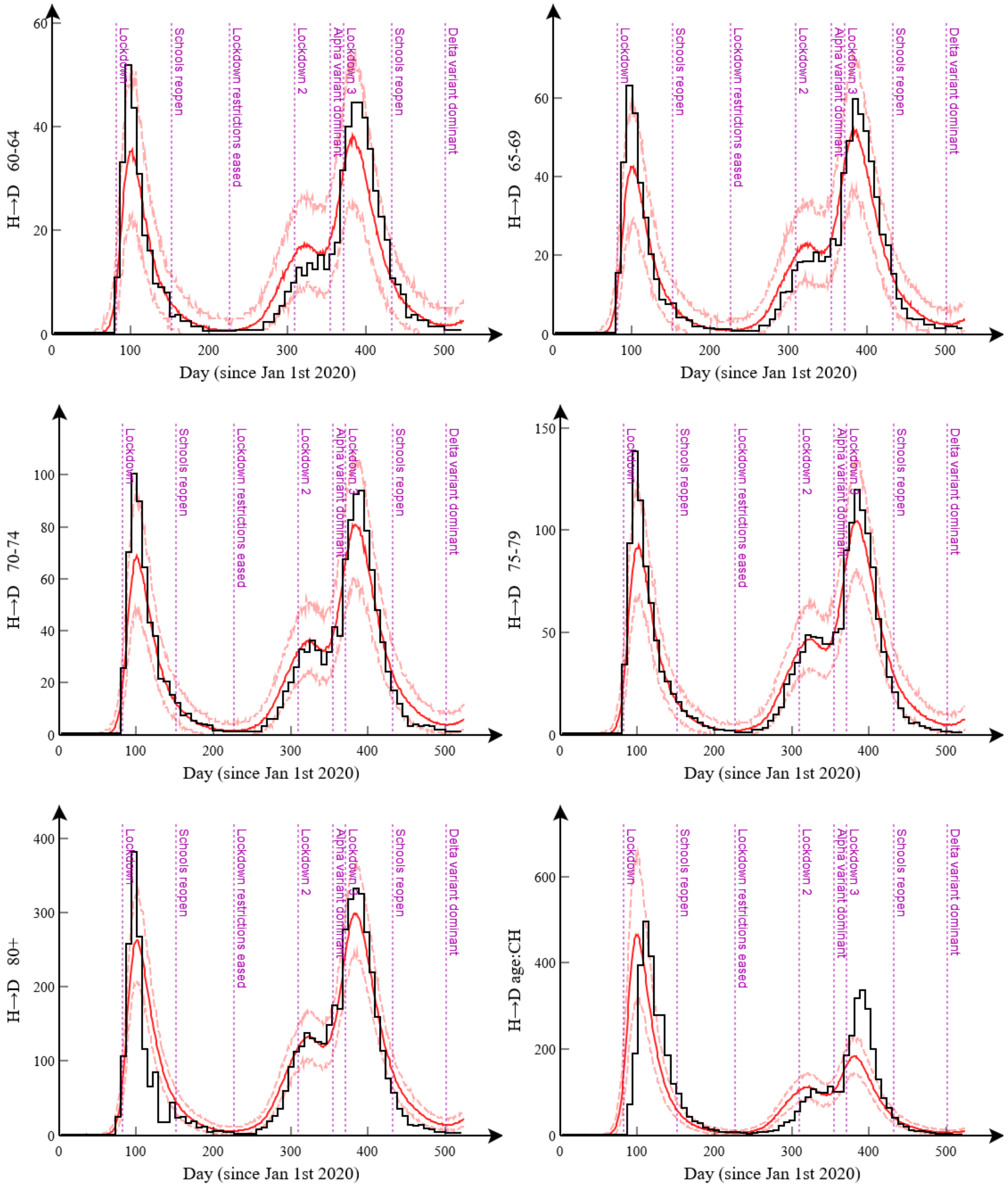


**Figure R1(f): Death data ages 60-CH**. This shows the posterior distribution for the inferred daily number of deaths (red solid line gives the posterior mean and the dashed lines denote 95% credible intervals) and the actual data (black line). Each graph represents a different age group. Note, when the death rate is low the data are amalgamated into longer time windows (to avoid Poisson noise).

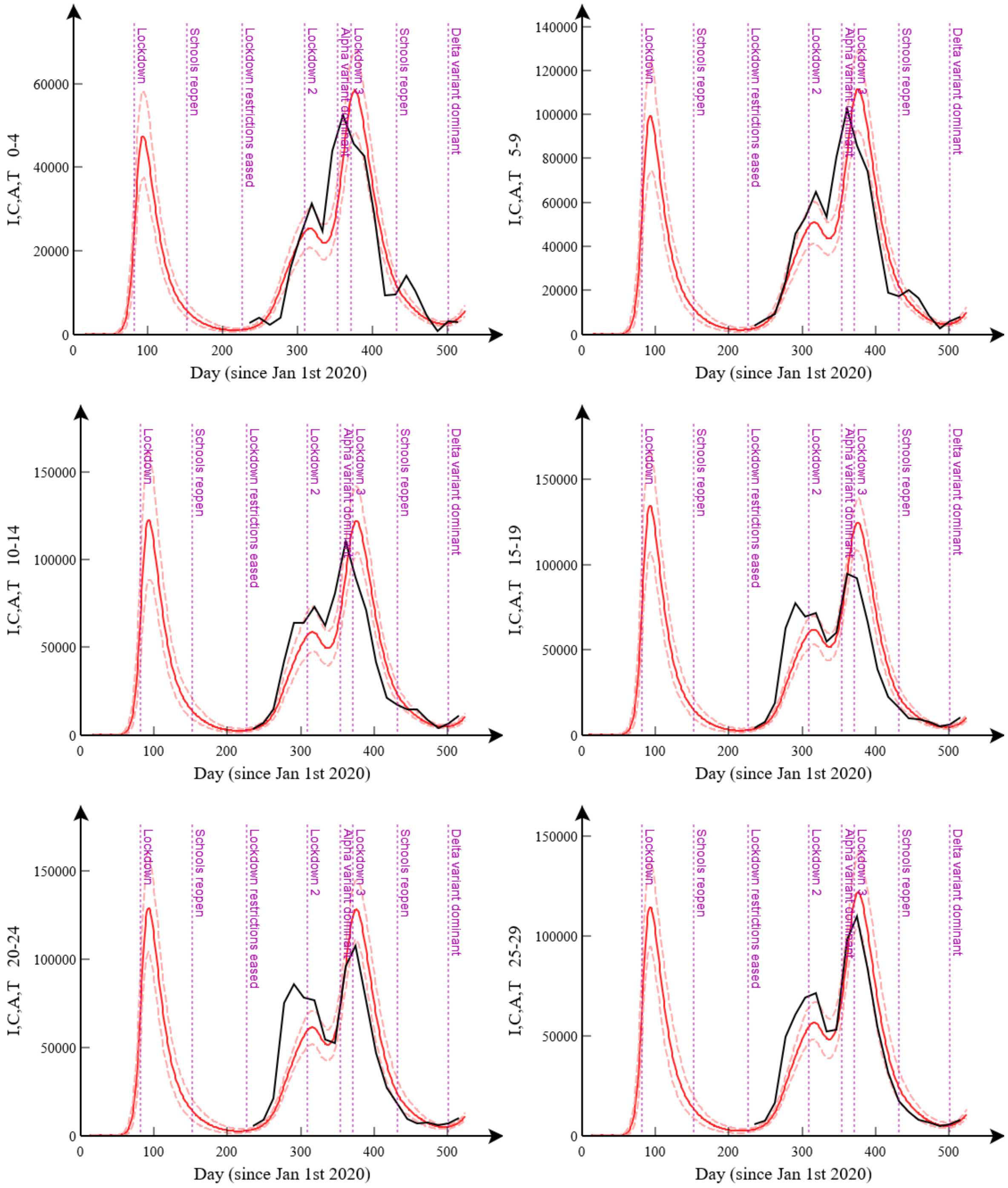


**Figure R1(g): PCR positive infected individuals ages 0-29**. This shows the posterior distribution for the inferred daily number of PCR positive individuals (red solid line gives the posterior mean and the dashed lines denote 95% credible intervals), *i.e.* the sum of the populations in the I, C, A and T compartments, and the data from the COVID-19 Infection Survey (black line). Each graph represents a different age group.

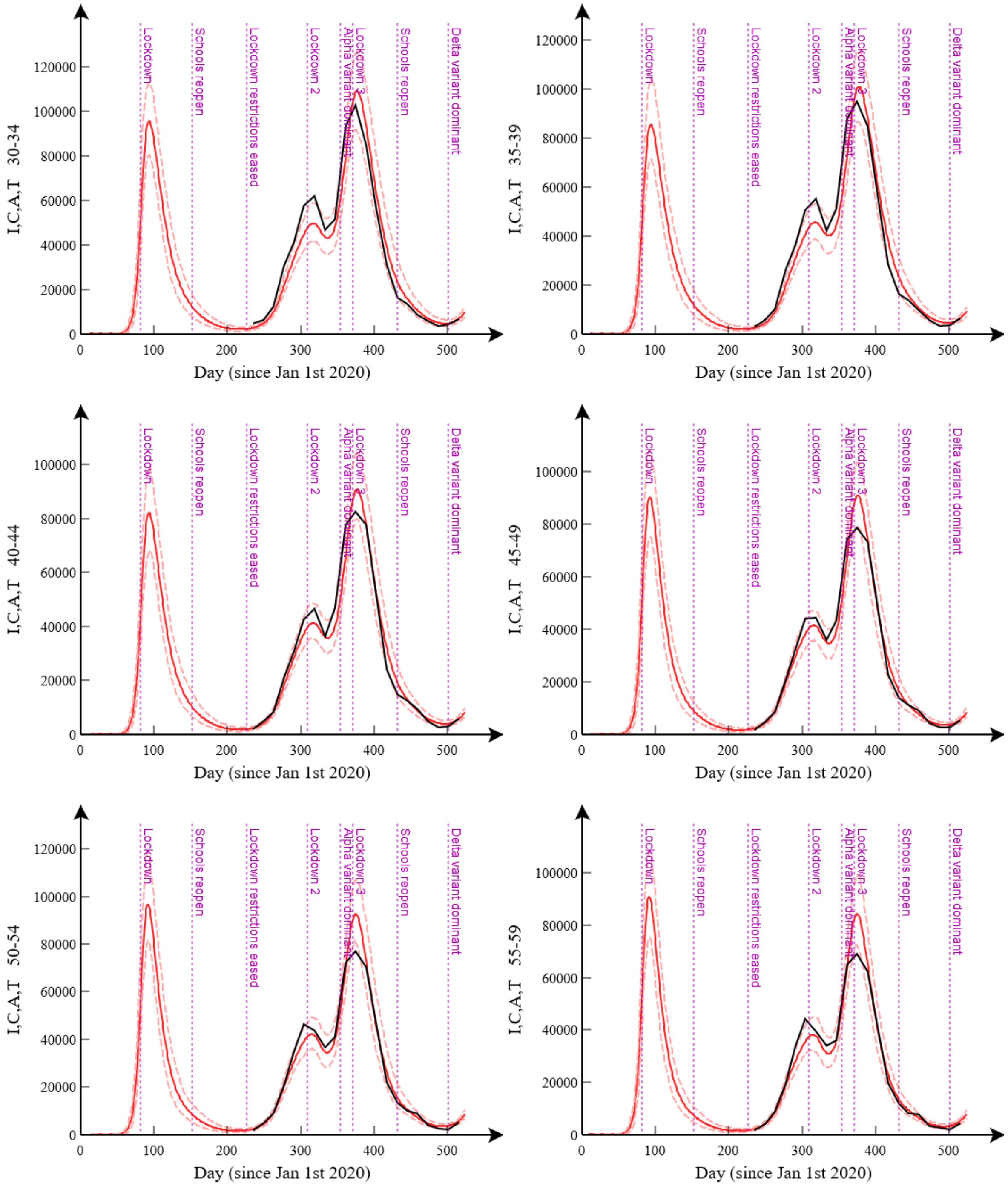


**Figure R1(h): PCR positive infected individuals ages 30-59**. This shows the posterior distribution for the inferred daily number of PCR positive individuals (red solid line gives the posterior mean and the dashed lines denote 95% credible intervals), *i.e.* the sum of the populations in the I, C, A and T compartments, and the data from the COVID-19 Infection Survey (black line). Each graph represents a different age group.

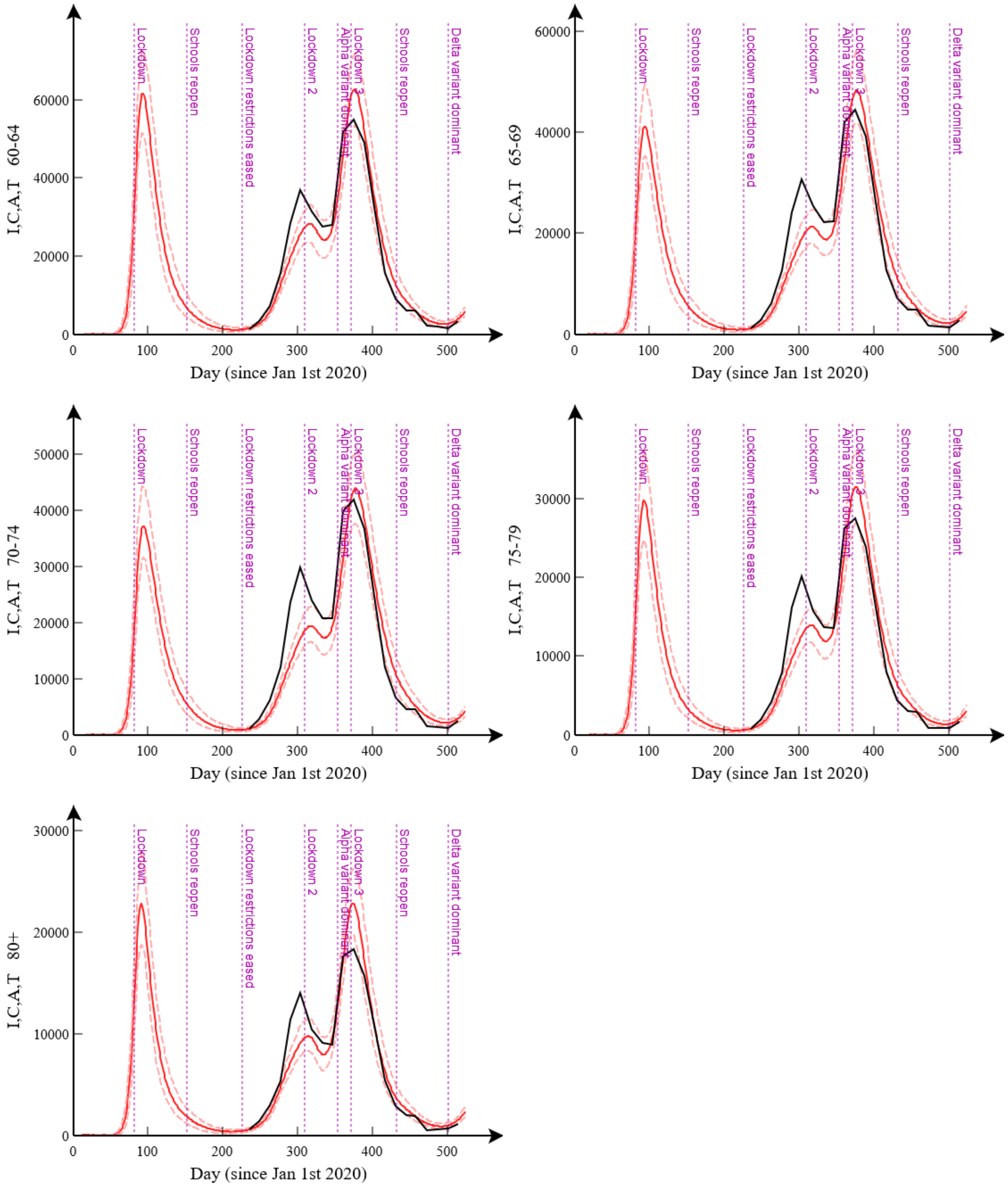


**Figure R1(i): PCR positive infected individuals ages 60-80+**. This shows the posterior distribution for the inferred daily number of PCR positive individuals (red solid line gives the posterior mean and the dashed lines denote 95% credible intervals), *i.e.* the sum of the populations in the I, C, A and T compartments, and the data from the COVID-19 Infection Survey (black line). Each graph represents a different age group.

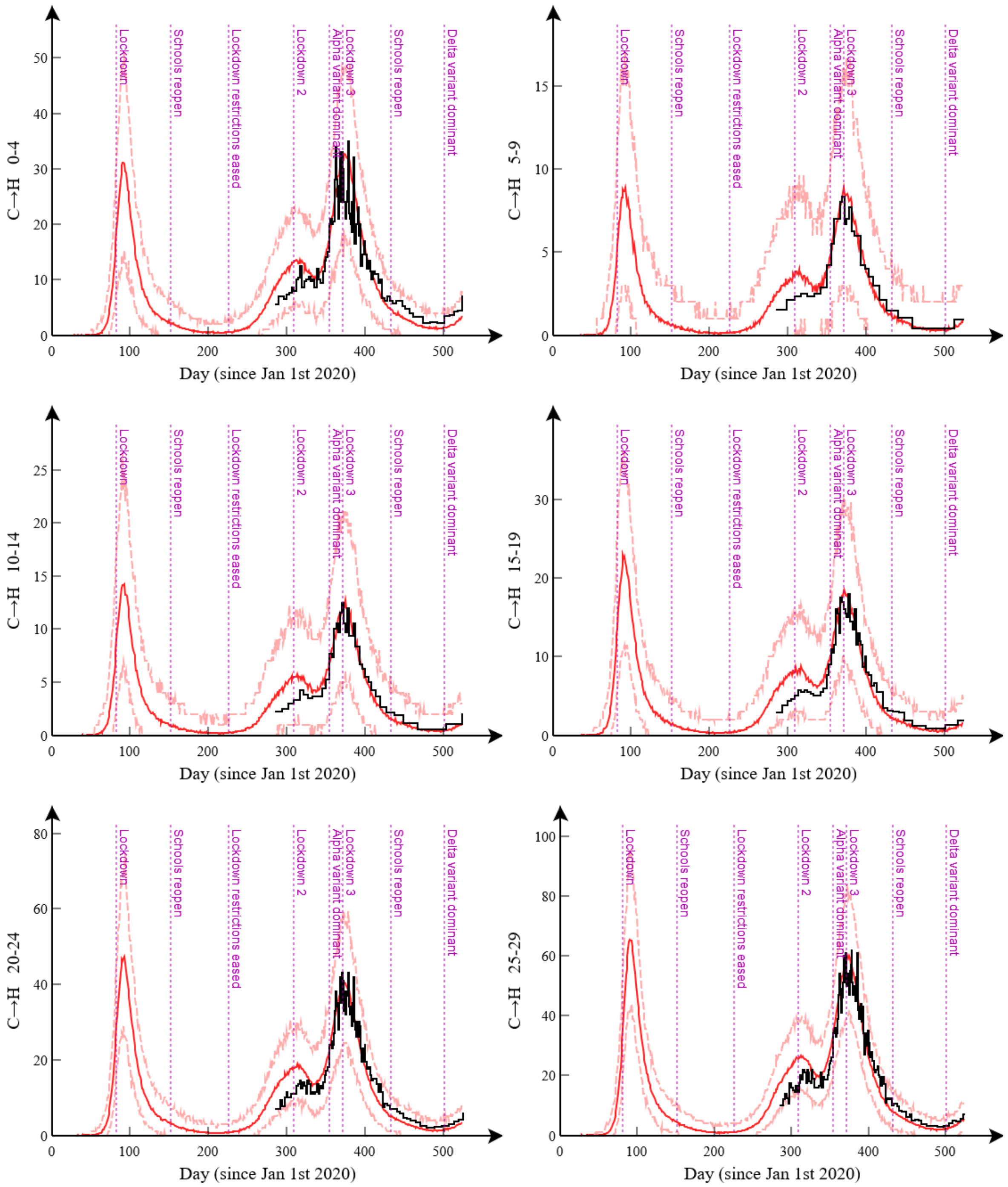


**Figure R1(j): Hospital admissions ages 0-29**. This shows the posterior distribution for the inferred daily number if transitions from the C to H compartments (red solid line gives the posterior mean and the dashed lines denote 95% credible intervals), and corresponding hospital admissions data (black line). Each graph represents a different age group.

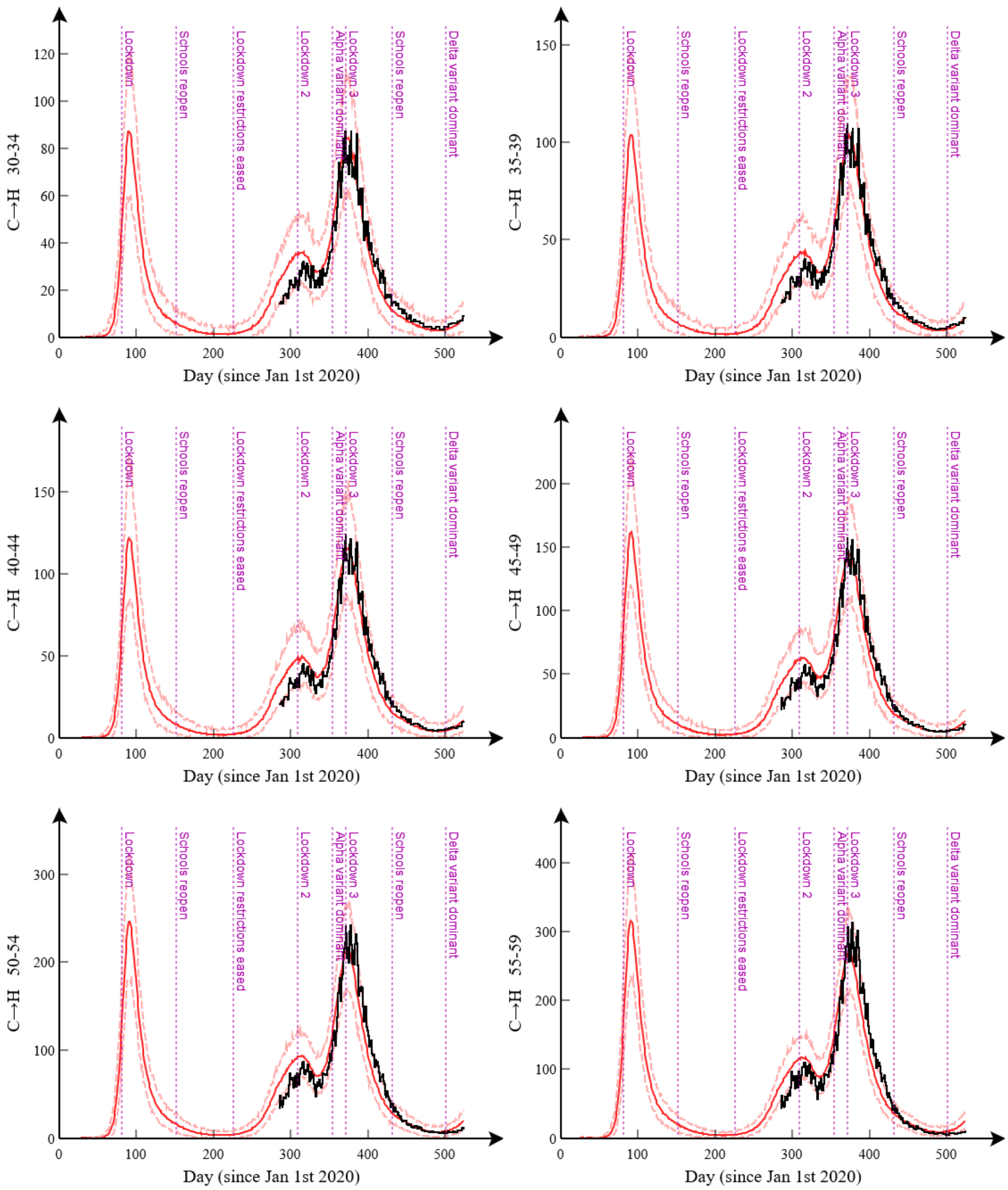


**Figure R1(k): Hospital admissions ages 30-59**. This shows the posterior distribution for the inferred daily number if transitions from the C to H compartments (red solid line gives the posterior mean and the dashed lines denote 95% credible intervals), and corresponding hospital admissions data (black line). Each graph represents a different age group.

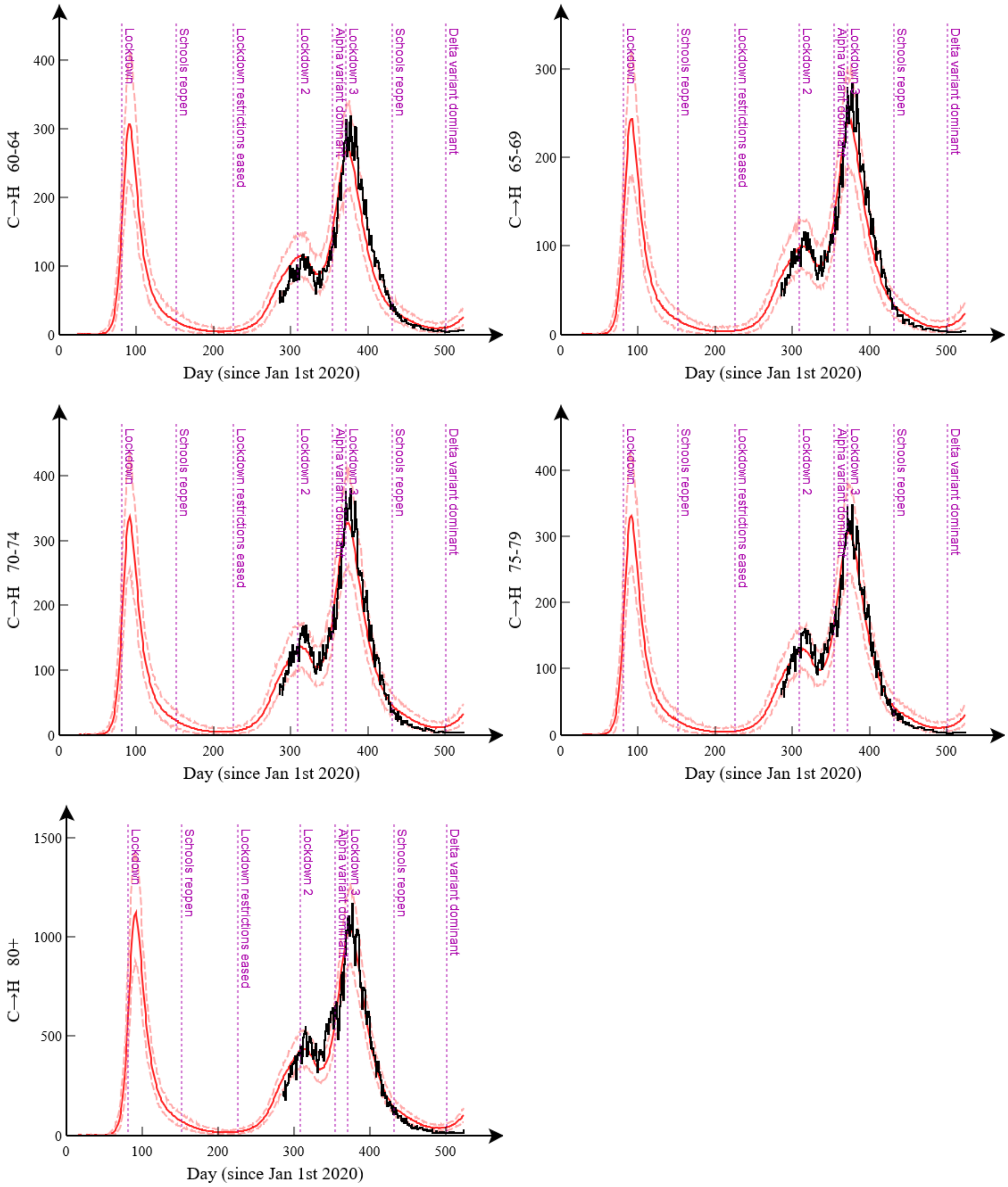


**Figure R1(l): Hospital admissions ages 60-80+**. This shows the posterior distribution for the inferred daily number if transitions from the C to H compartments (red solid line gives the posterior mean and the dashed lines denote 95% credible intervals), and corresponding hospital admissions data (black line). Each graph represents a different age group.

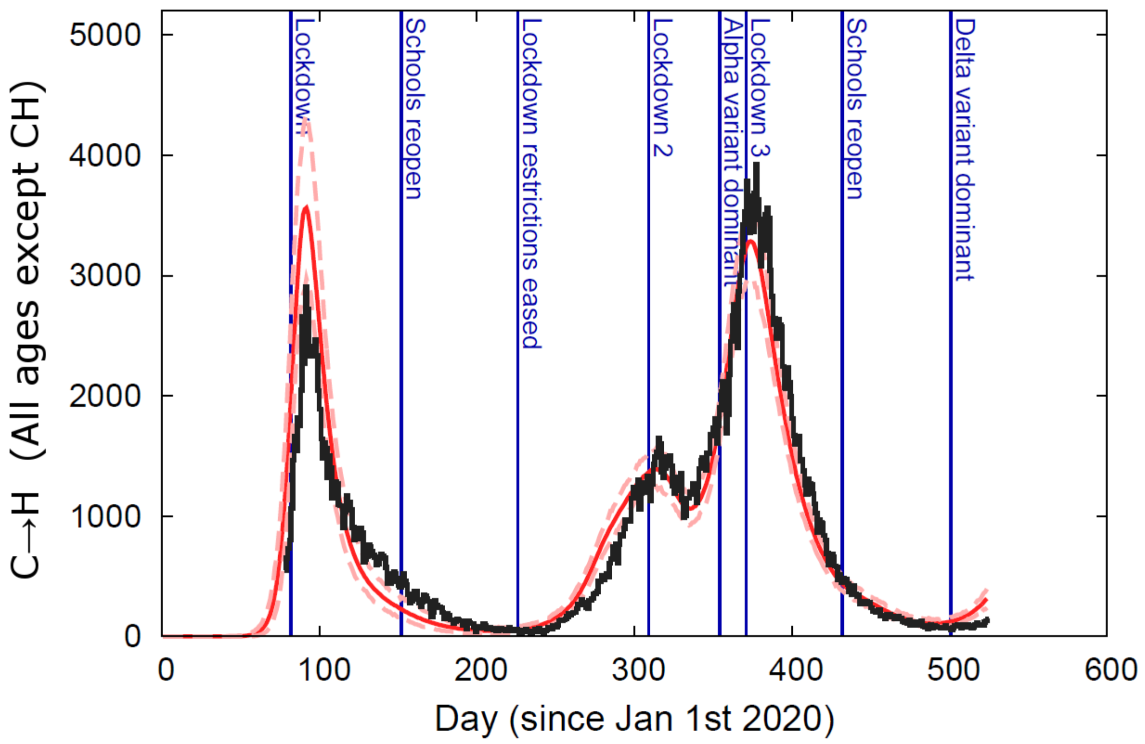


**Figure R1(m): Hospital admissions**. This shows the posterior distribution for the inferred daily number if transitions from the C to H compartments (red solid line gives the posterior mean and the dashed lines denote 95% credible intervals), for all ages except for the care home CH category, and corresponding hospital admissions data (black line).

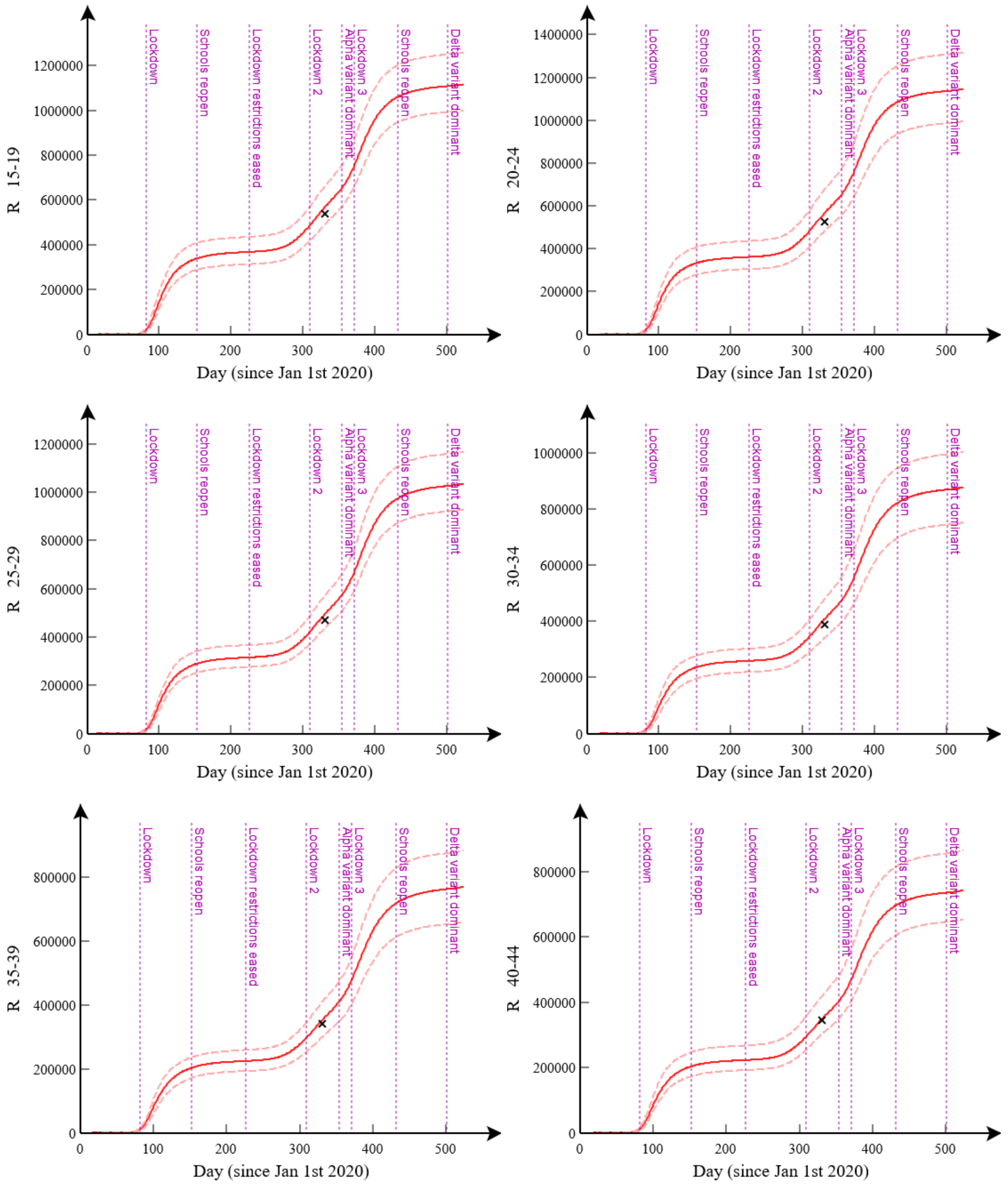


**Figure R1(n): Antibody seropositive 15-44**. This shows the posterior distribution for the inferred number of individuals in the R compartment (red solid line gives the posterior mean and the dashed lines denote 95% credible intervals), and corresponding antibody seropositive data (black cross), which was taken from the very initial phase of the COVID-19 infection survey (importantly before vaccination had begun). Each graph represents a different age group.

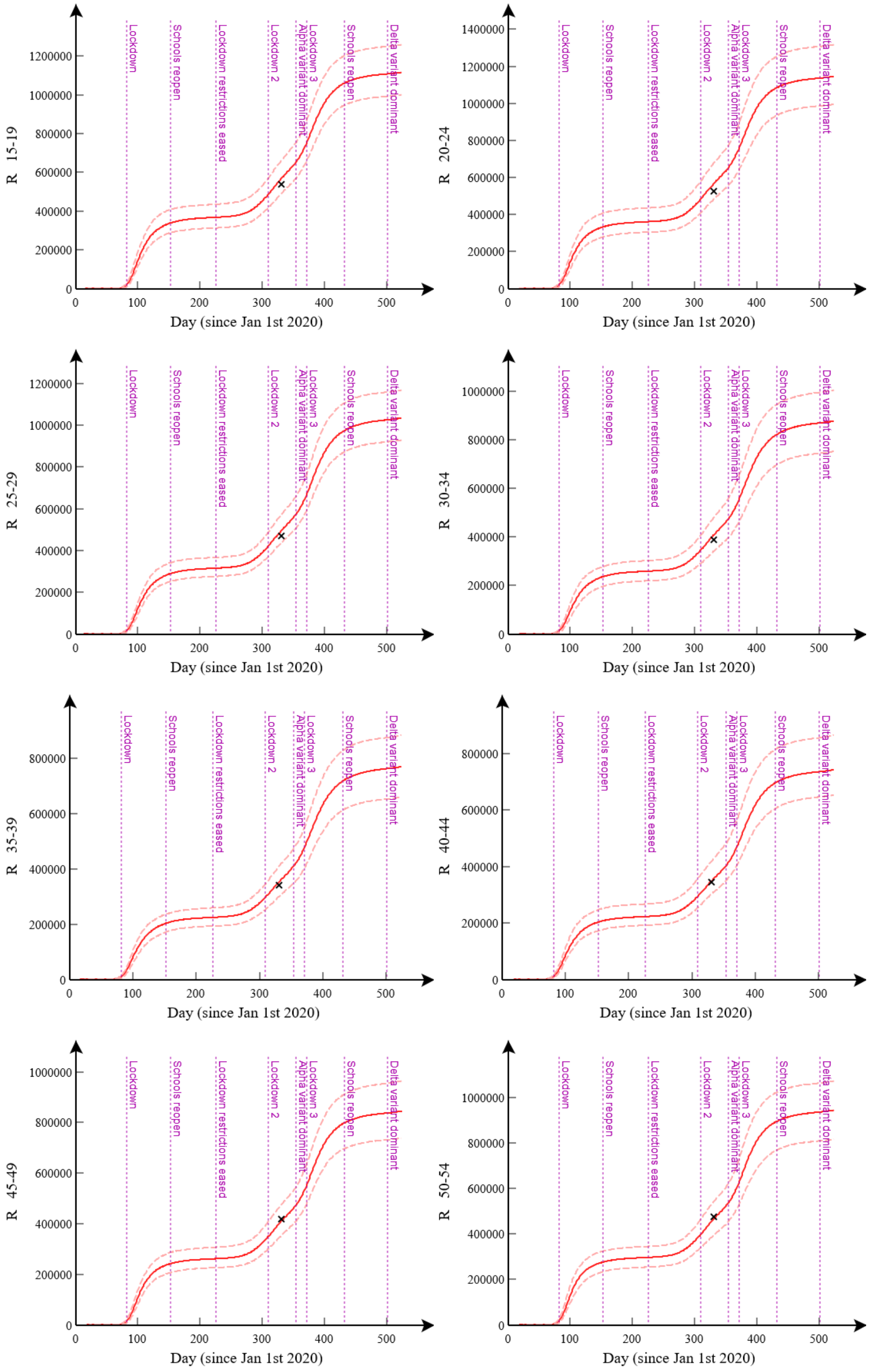


**Figure R1(o): Antibody seropositive 15-54**. This shows the posterior distribution for the inferred number of individuals in the R compartment (red solid line gives the posterior mean and the dashed lines denote 95% credible intervals), and corresponding antibody seropositive data (black cross), which was taken from the very initial phase of the COVID-19 infection survey (importantly before vaccination had begun). Each graph represents a different age group.

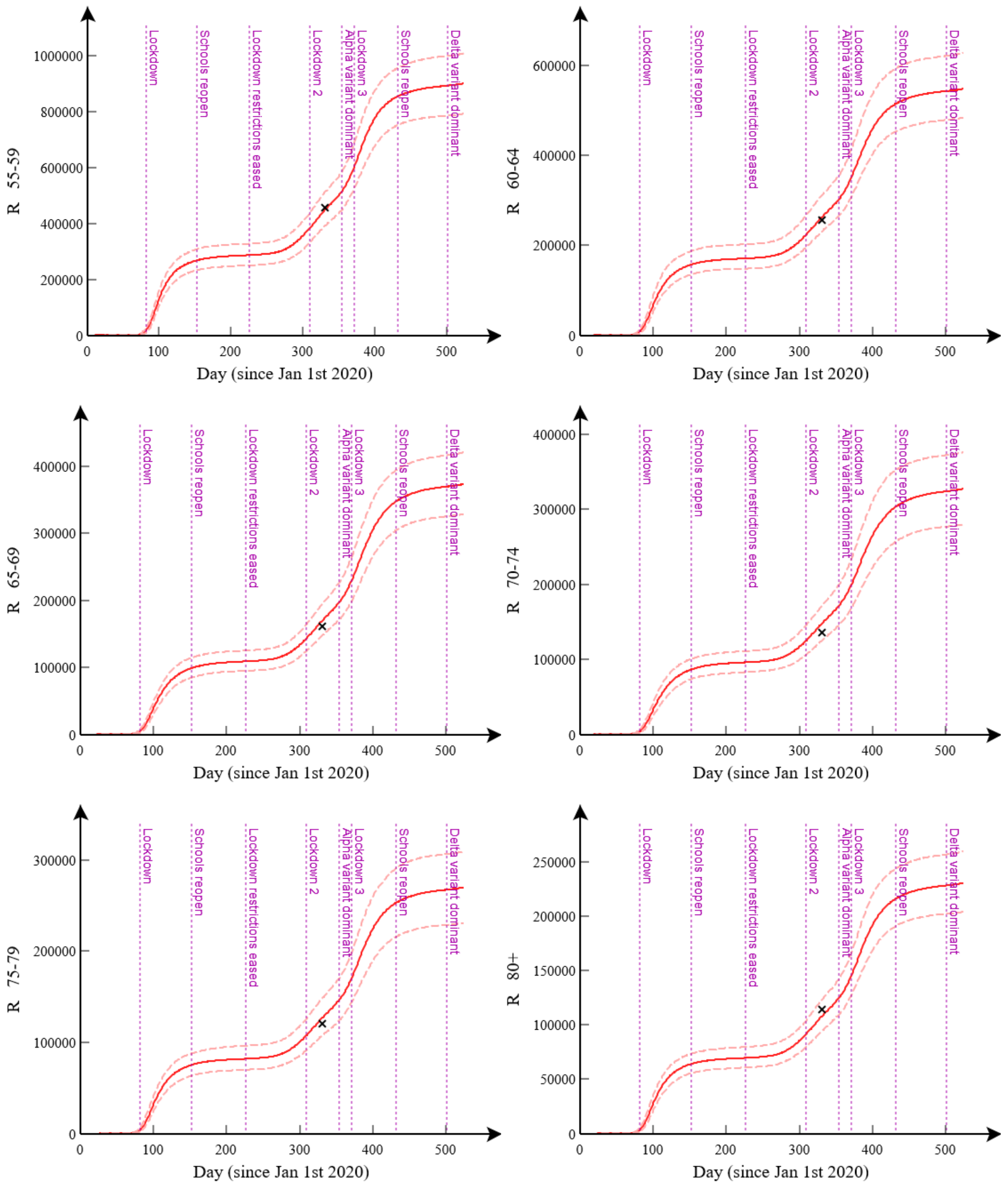


**Figure R1(p): Antibody seropositive 55-80+**. This shows the posterior distribution for the inferred number of individuals in the R compartment (red solid line gives the posterior mean and the dashed lines denote 95% credible intervals), and corresponding antibody seropositive data (black cross), which was taken from the very initial phase of the COVID-19 infection survey (importantly before vaccination had begun). Each graph represents a different age group.

### 3) Convergence over generations

The series of figures below show how posterior estimates for different model parameters change over generations $g$ up to the maximum generation number $G$=350. They show the progression of particles from direct samples of the prior (at $g$=1) to good posterior estimates ($g$=$G$). Pushing beyond $G$=350 may yield slightly better estimates for the posterior, but this would entail significantly more computational effort (generations become successively more computationally expensive to perform). Results from validation of the algorithm in Appendix N in the Supplementary Material suggest that $G$=350 is sufficiently high to estimate parameters with a high degree of accuracy.

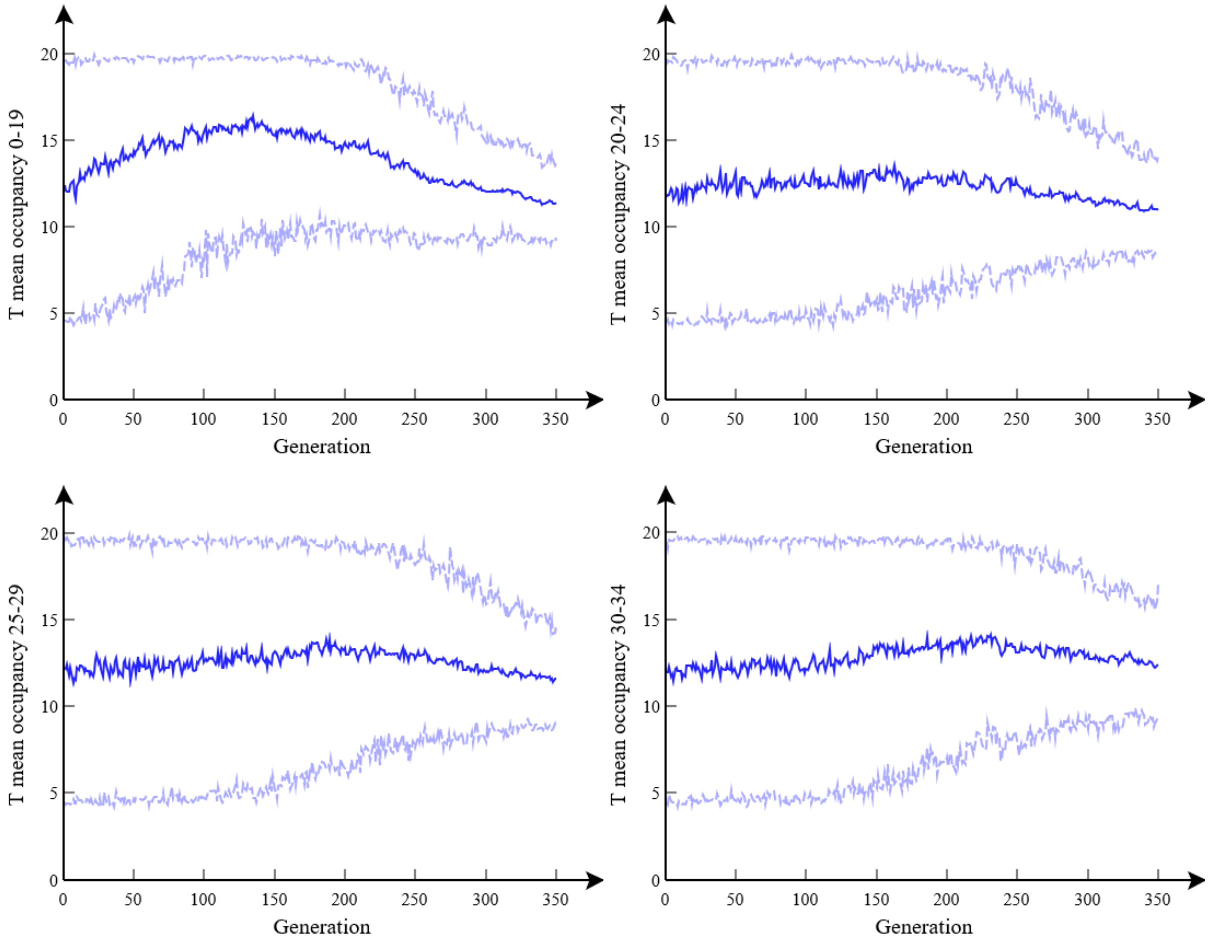


**Figure R2(a): Convergence of T residency time**. This shows how posterior estimates for different model parameters vary with generation number $g$ (up to a maximum $G$=350 used for the final posterior estimates). These graphs combine together $R$=16 separate runs each with $P$=16 particles. Blue lines show posterior means, with light blue dashed lines denoting 95% credible intervals.

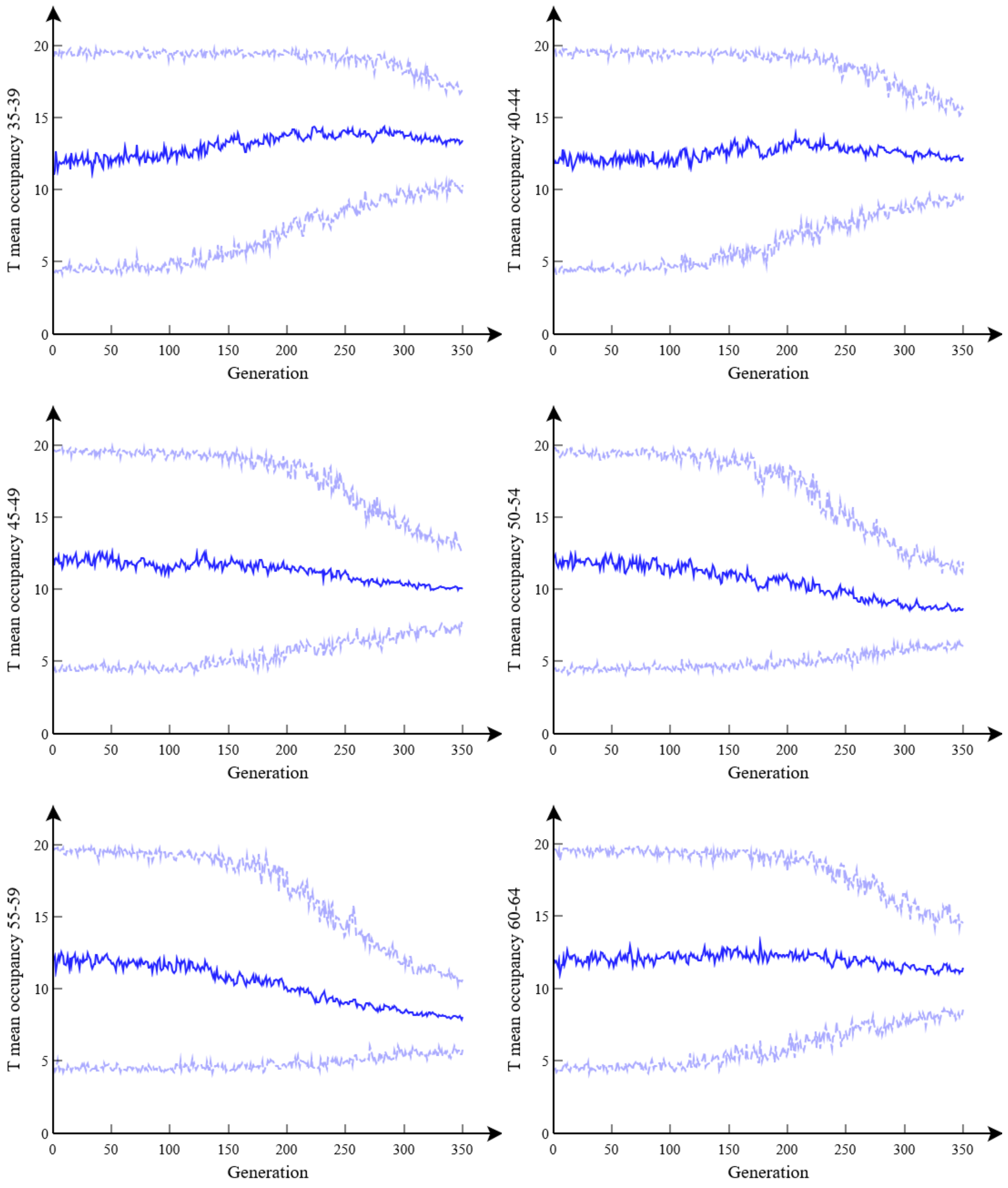


**Figure R2(b): Convergence of T residency time**. This shows how posterior estimates for different model parameters vary with generation number *g* (up to a maximum *G*=350 used for the final posterior estimates). These graphs combine together *R*=16 separate runs each with *P*=16 particles. Blue lines show posterior means, with light blue dashed lines denoting 95% credible intervals.

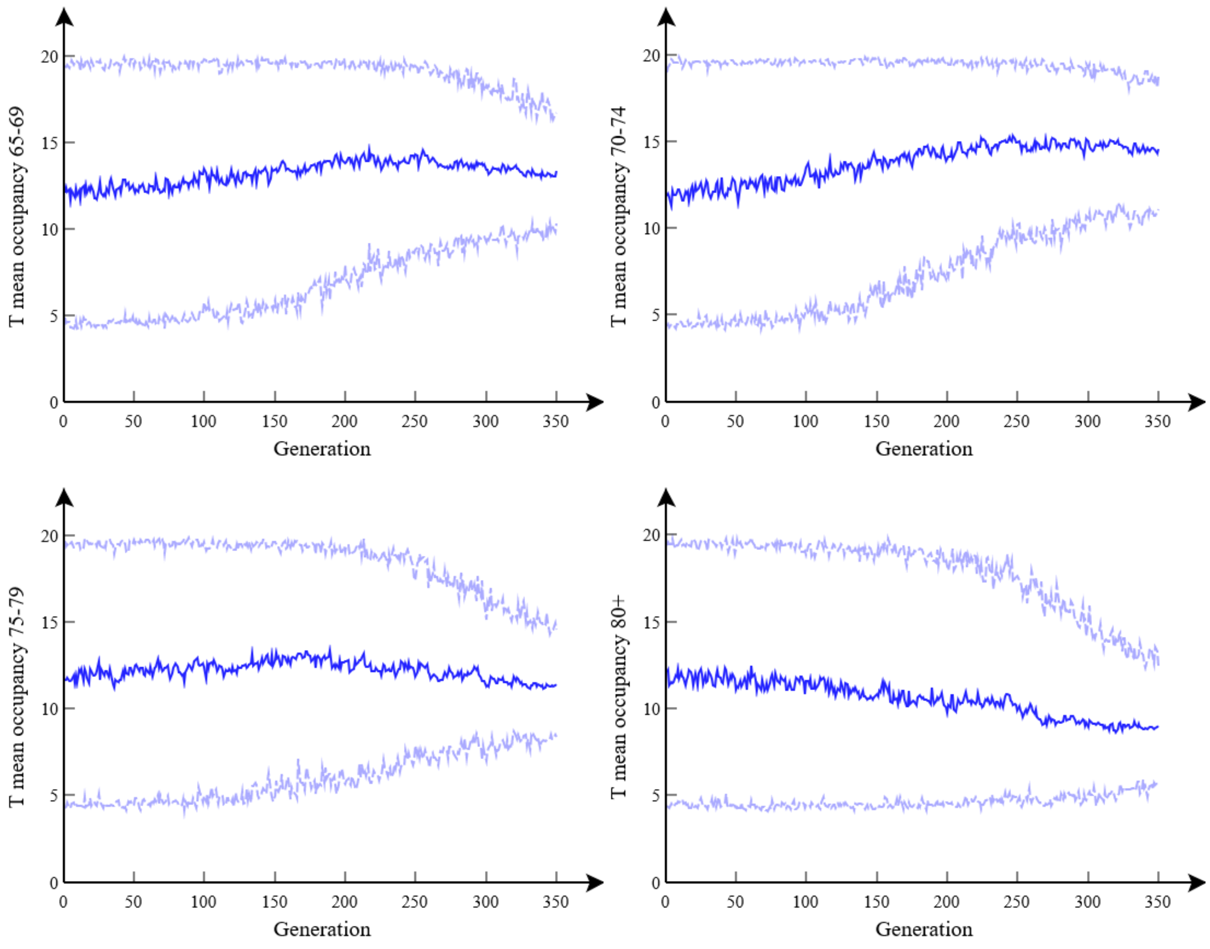


**Figure R2(c): Convergence of T residency time**. This shows how posterior estimates for different model parameters vary with generation number *g* (up to a maximum *G*=350 used for the final posterior estimates). These graphs combine together *R*=16 separate runs each with *P*=16 particles. Blue lines show posterior means, with light blue dashed lines denoting 95% credible intervals.

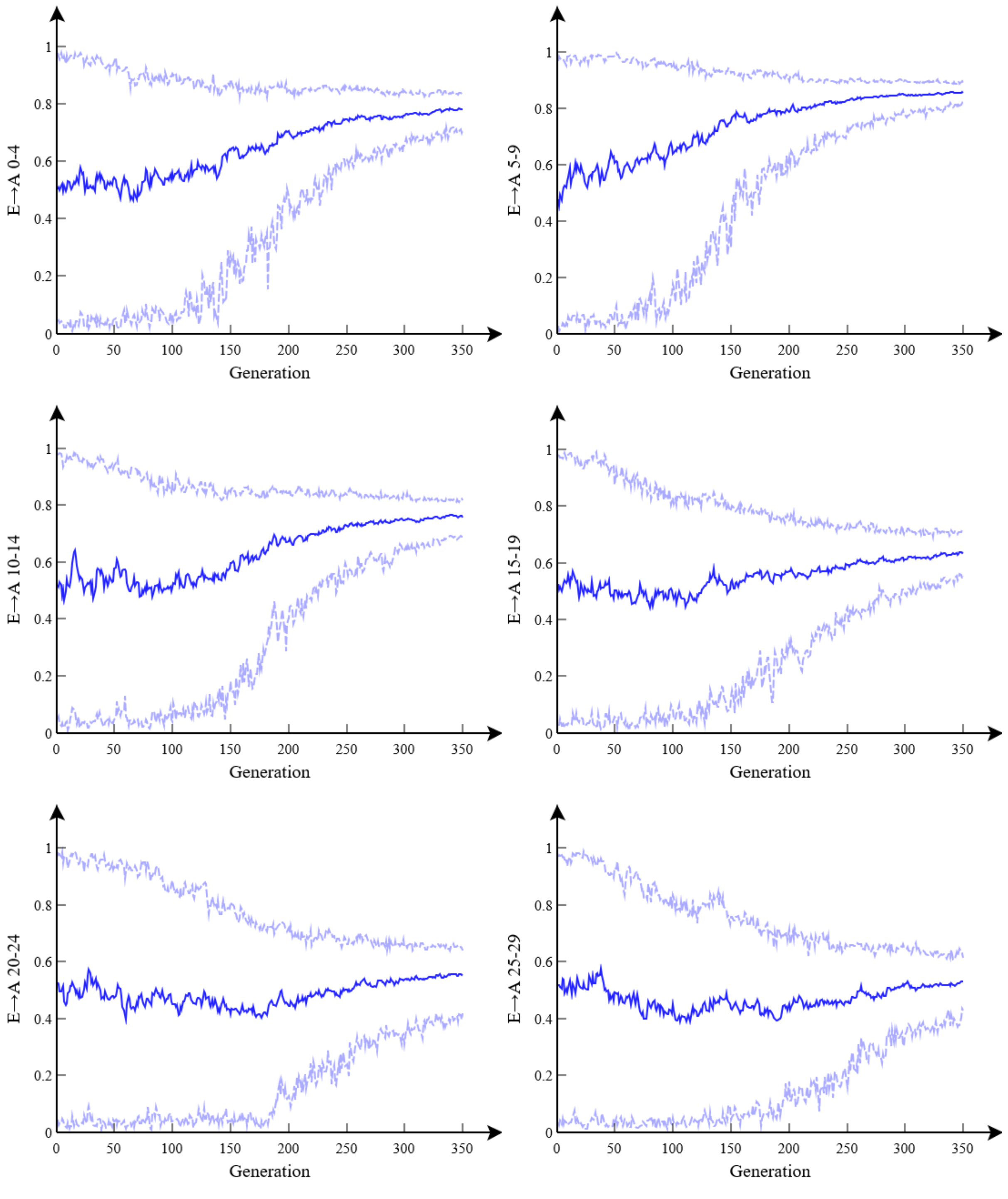


**Figure R2(d): Asymptomatic branching probability $b_a^{E \to A}$.** This shows how posterior estimates for different model parameters vary with generation number *g* (up to a maximum *G*=350 used for the final posterior estimates). These graphs combine together *R*=16 separate runs each with *P*=16 particles. Blue lines show posterior means, with light blue dashed lines denoting 95% credible intervals.

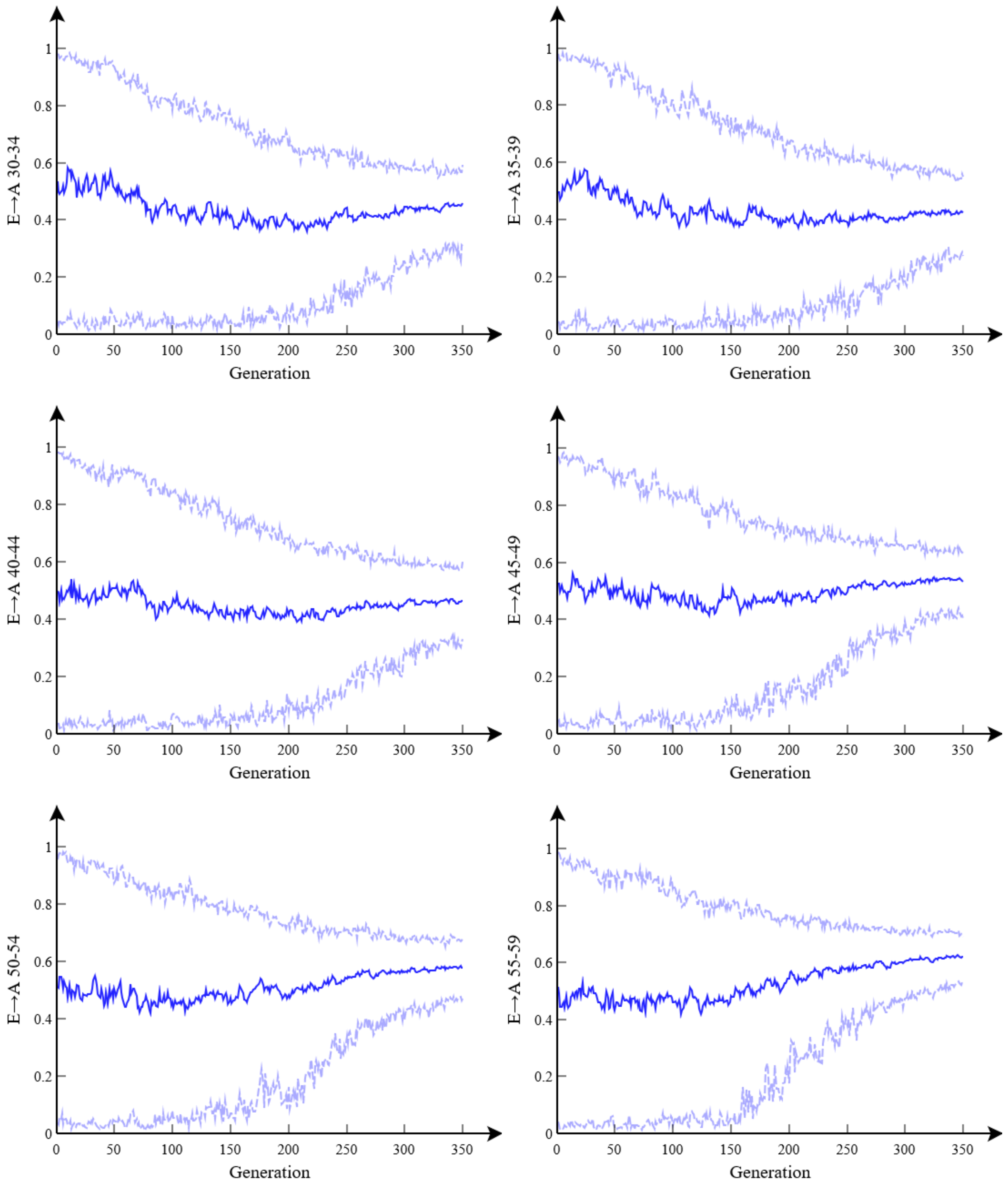


**Figure R2(e): Asymptomatic branching probability $b_a^{E\to A}$.** This shows how posterior estimates for different model parameters vary with generation number *g* (up to a maximum *G*=350 used for the final posterior estimates). These graphs combine together *R*=16 separate runs each with *P*=16 particles. Blue lines show posterior means, with light blue dashed lines denoting 95% credible intervals.

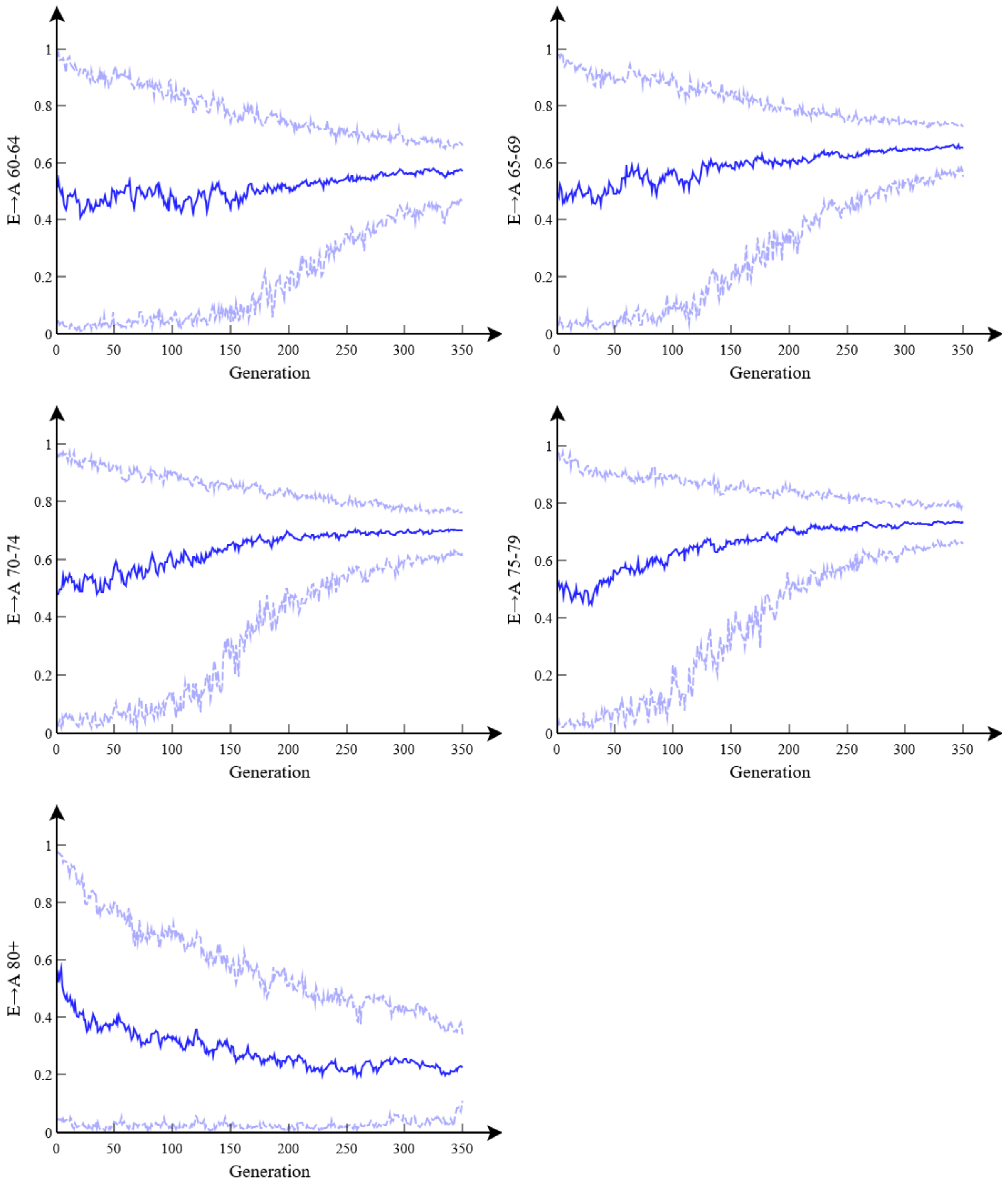


**Figure R2(f): Asymptomatic branching probability $b_a^{E\to A}$.** This shows how posterior estimates for different model parameters vary with generation number $g$ (up to a maximum $G$=350 used for the final posterior estimates). These graphs combine together $R$=16 separate runs each with $P$=16 particles. Blue lines show posterior means, with light blue dashed lines denoting 95% credible intervals.

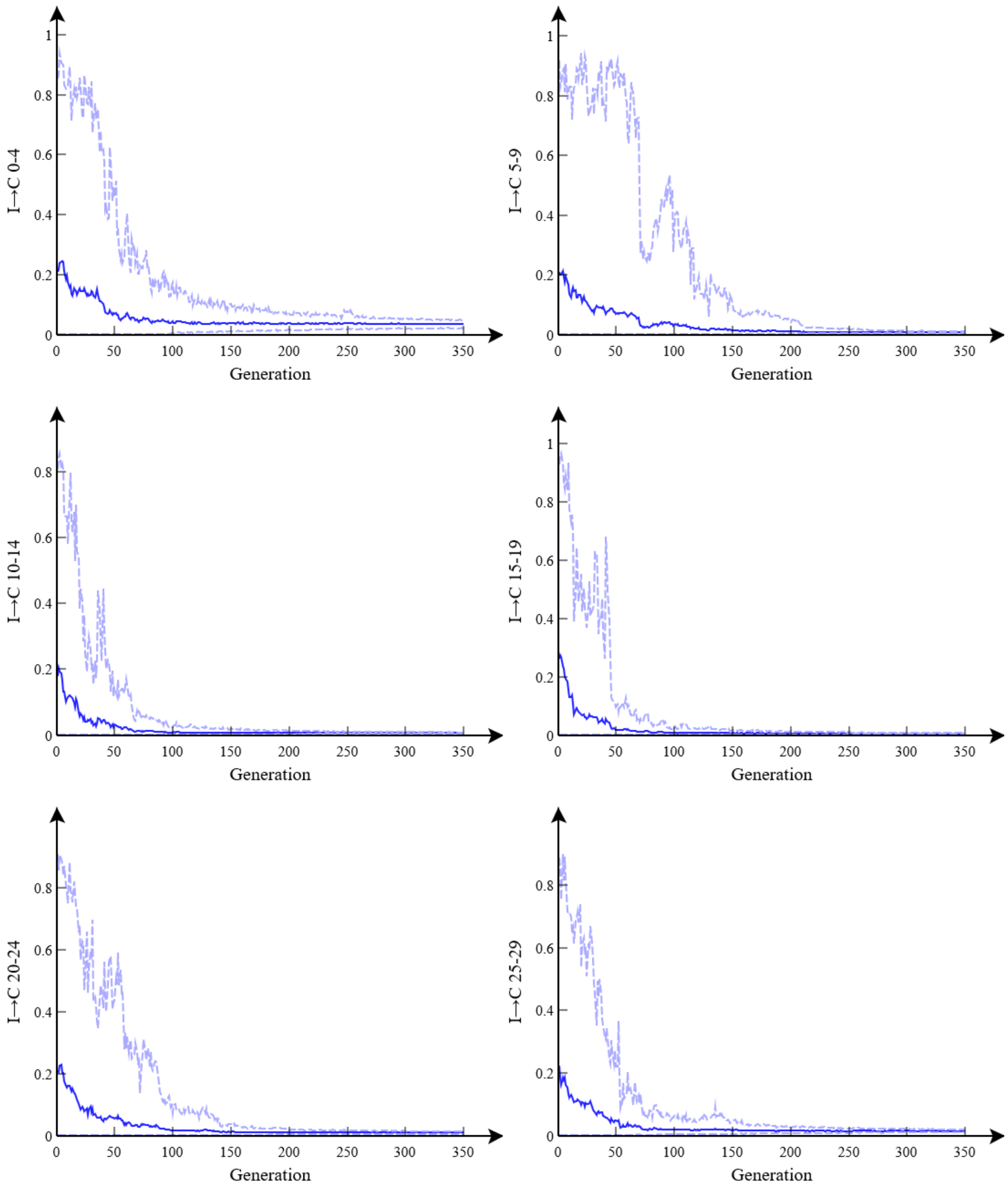


**Figure R2(g): Hospitalised branching probability $b_a^{I\to C}$.** This shows how posterior estimates for different model parameters vary with generation number *g* (up to a maximum *G*=350 used for the final posterior estimates). These graphs combine together *R*=16 separate runs each with *P*=16 particles. Blue lines show posterior means, with light blue dashed lines denoting 95% credible intervals.

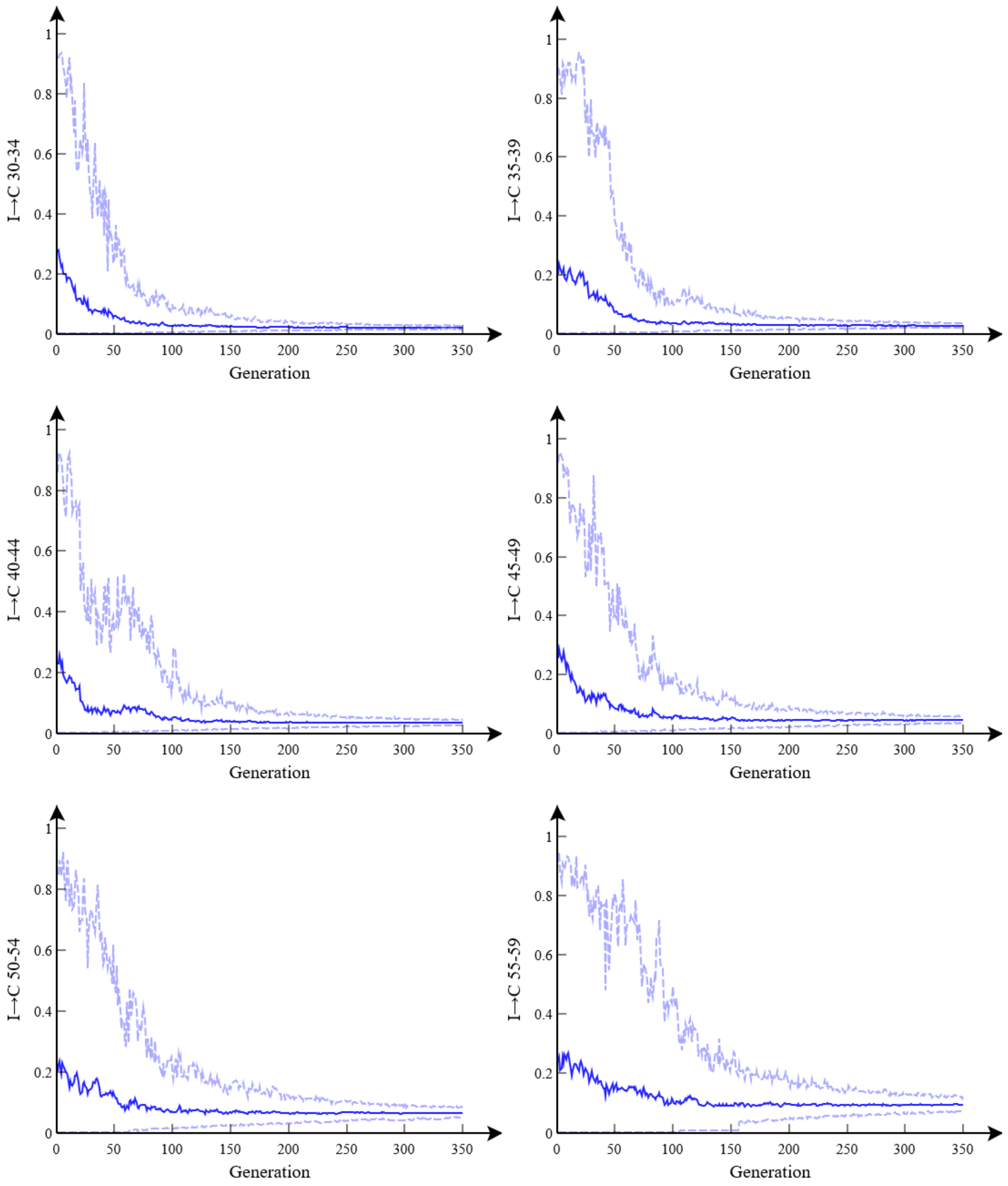


**Figure R2(h): Hospitalised branching probability $b_a^{I\to C}$.** This shows how posterior estimates for different model parameters vary with generation number *g* (up to a maximum *G*=350 used for the final posterior estimates). These graphs combine together *R*=16 separate runs each with *P*=16 particles. Blue lines show posterior means, with light blue dashed lines denoting 95% credible intervals.

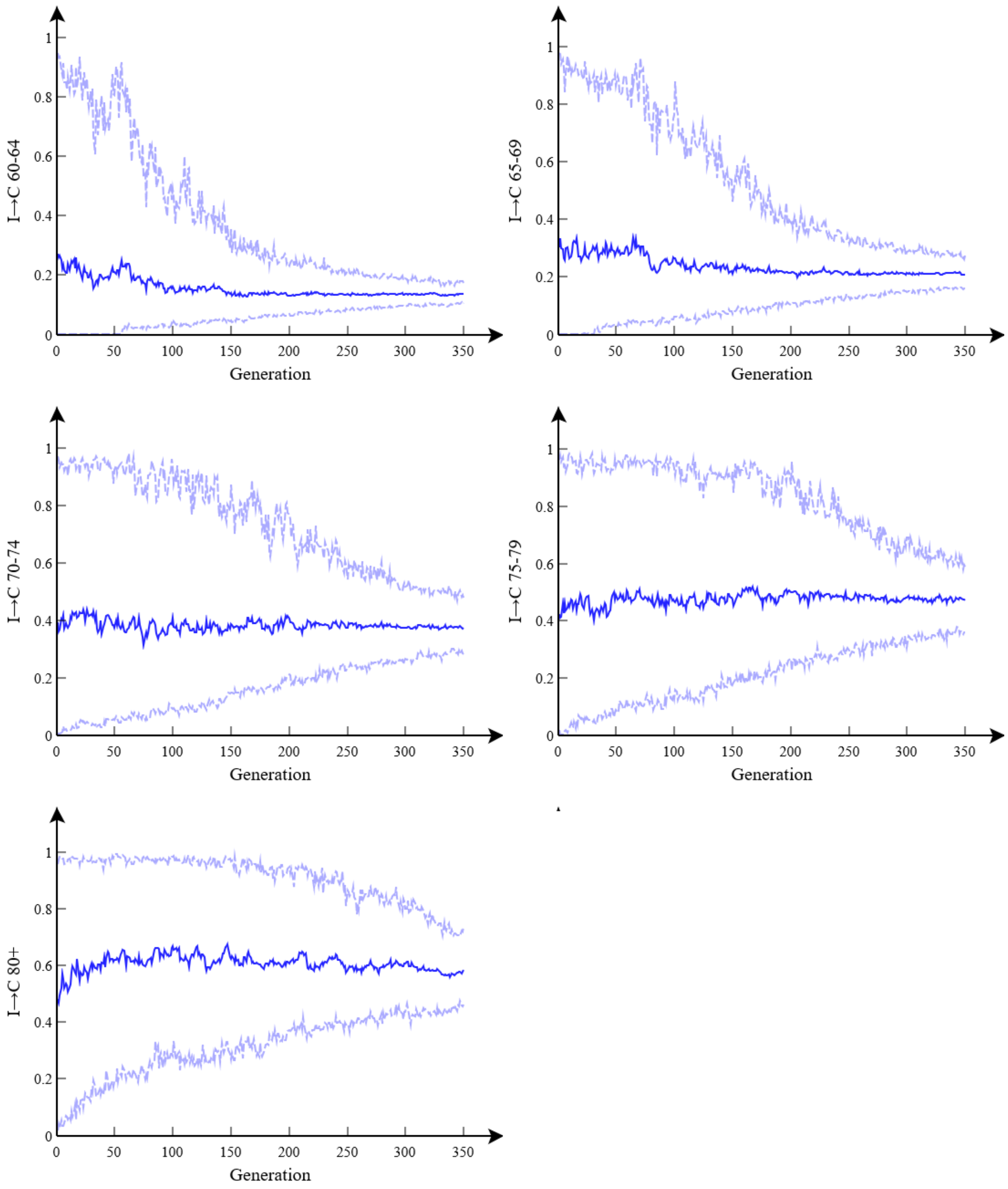


**Figure R2(i): Hospitalised branching probability $b_a^{I\to C}$.** This shows how posterior estimates for different model parameters vary with generation number $g$ (up to a maximum $G$=350 used for the final posterior estimates). These graphs combine together $R$=16 separate runs each with $P$=16 particles. Blue lines show posterior means, with light blue dashed lines denoting 95% credible intervals.

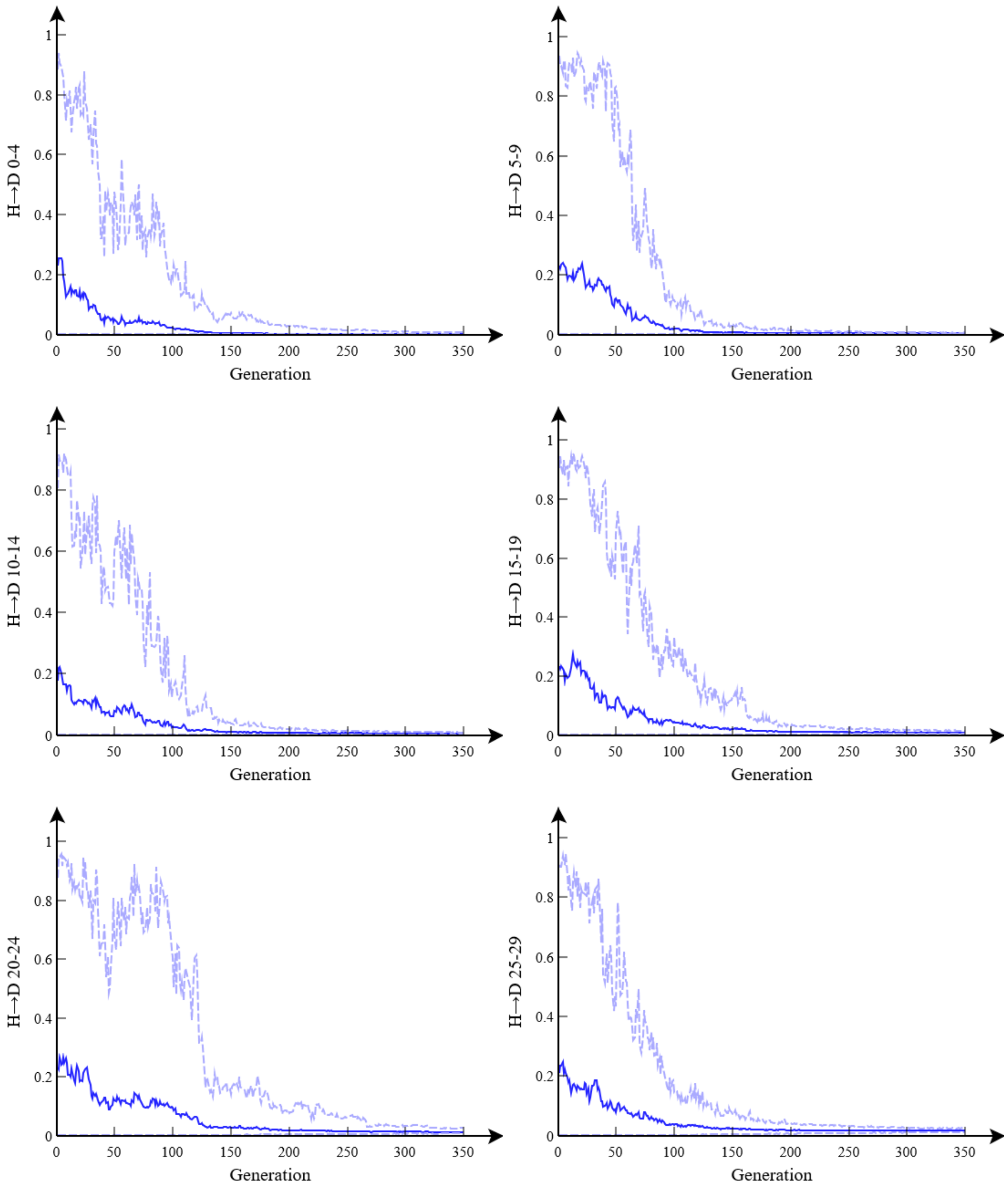


**Figure R2(j): Death branching probability $b_a^{H\to D}$.** This shows how posterior estimates for different model parameters vary with generation number $g$ (up to a maximum $G$=350 used for the final posterior estimates). These graphs combine together $R$=16 separate runs each with $P$=16 particles. Blue lines show posterior means, with light blue dashed lines denoting 95% credible intervals.

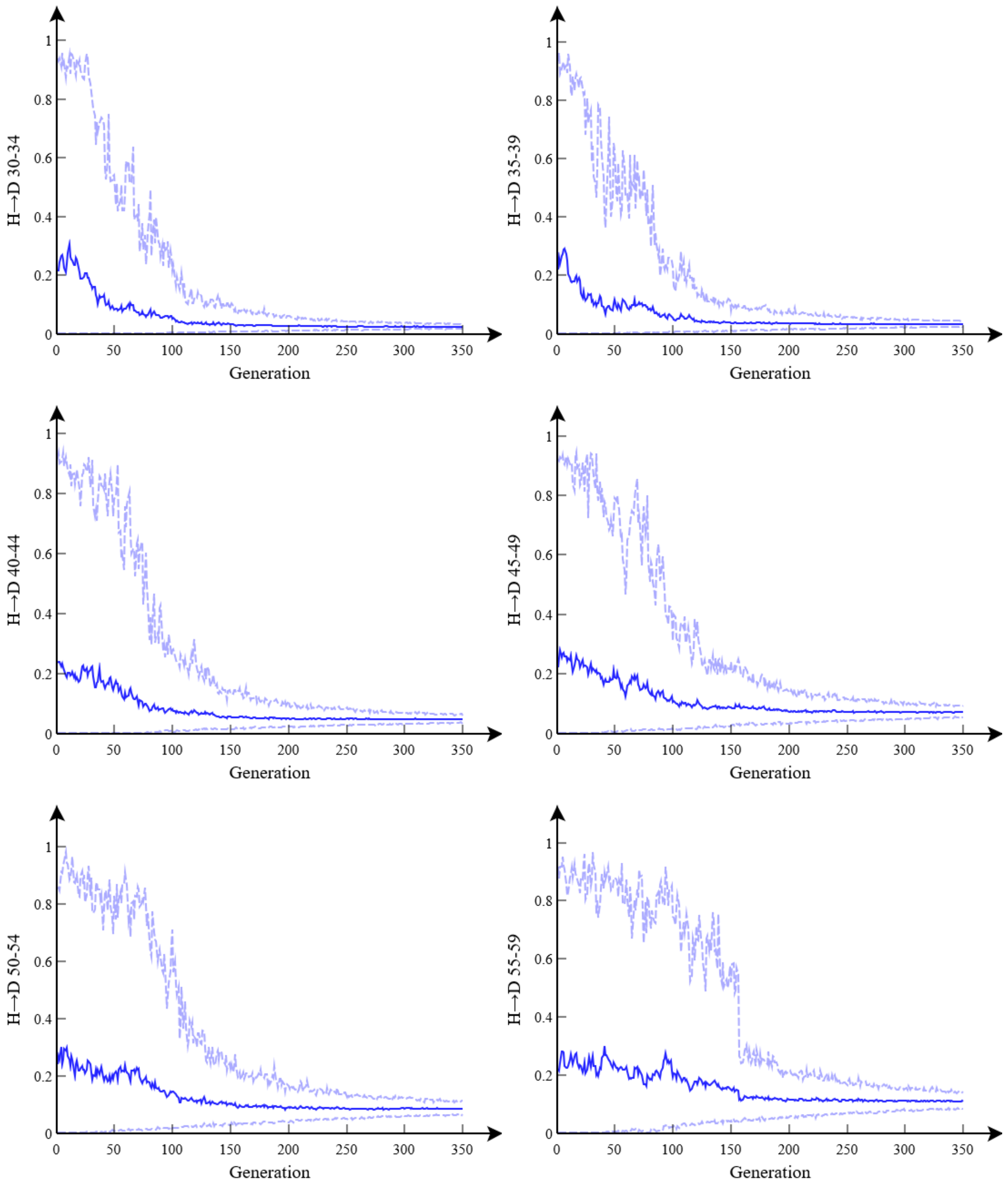


**Figure R2(k): Death branching probability $b_a^{H\to D}$.** This shows how posterior estimates for different model parameters vary with generation number $g$ (up to a maximum $G$=350 used for the final posterior estimates). These graphs combine together $R$=16 separate runs each with $P$=16 particles. Blue lines show posterior means, with light blue dashed lines denoting 95% credible intervals.

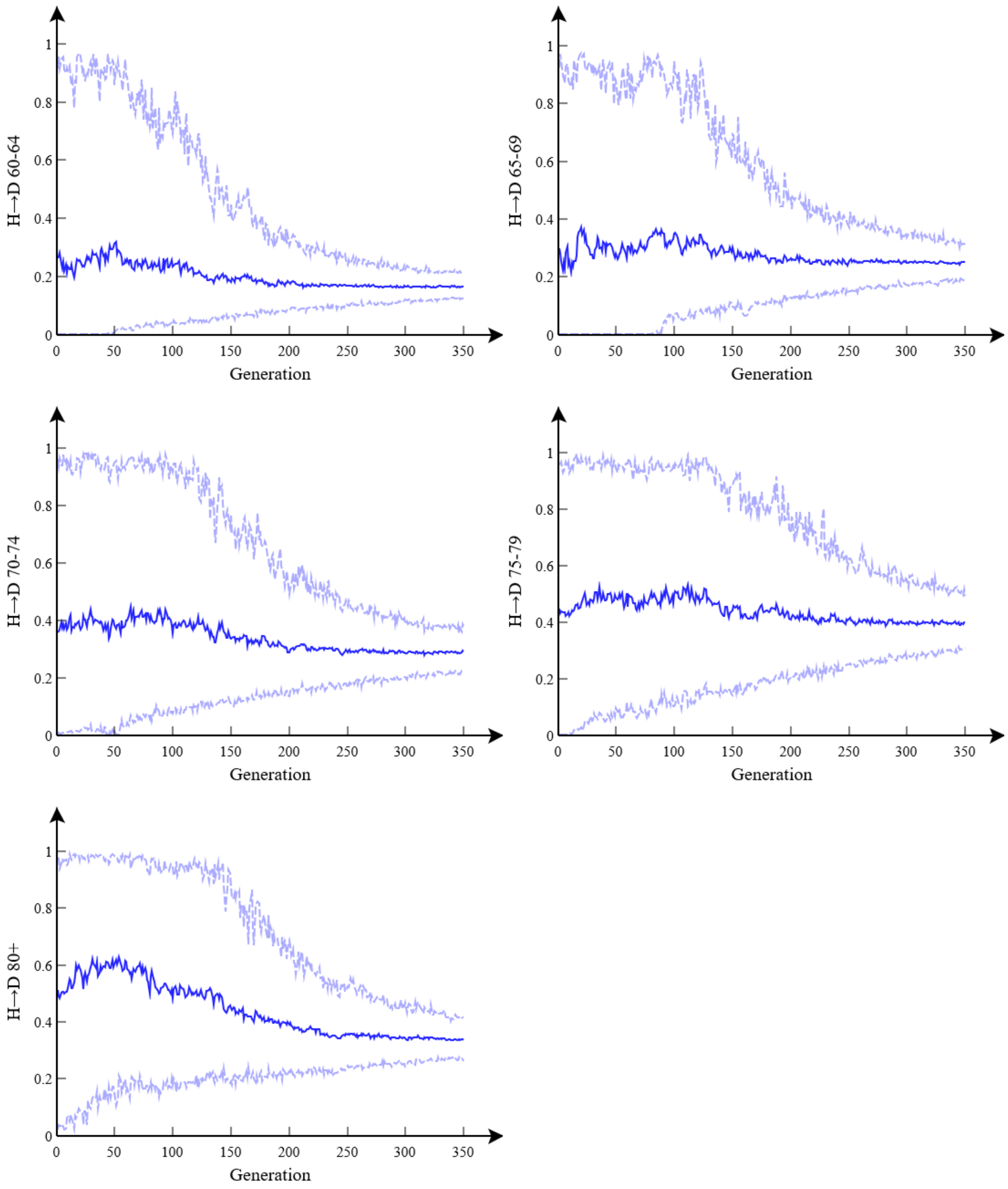


**Figure R2(l): Death branching probability $b_a^{H\to D}$.** This shows how posterior estimates for different model parameters vary with generation number $g$ (up to a maximum $G$=350 used for the final posterior estimates). These graphs combine together $R$=16 separate runs each with $P$=16 particles. Blue lines show posterior means, with light blue dashed lines denoting 95% credible intervals.

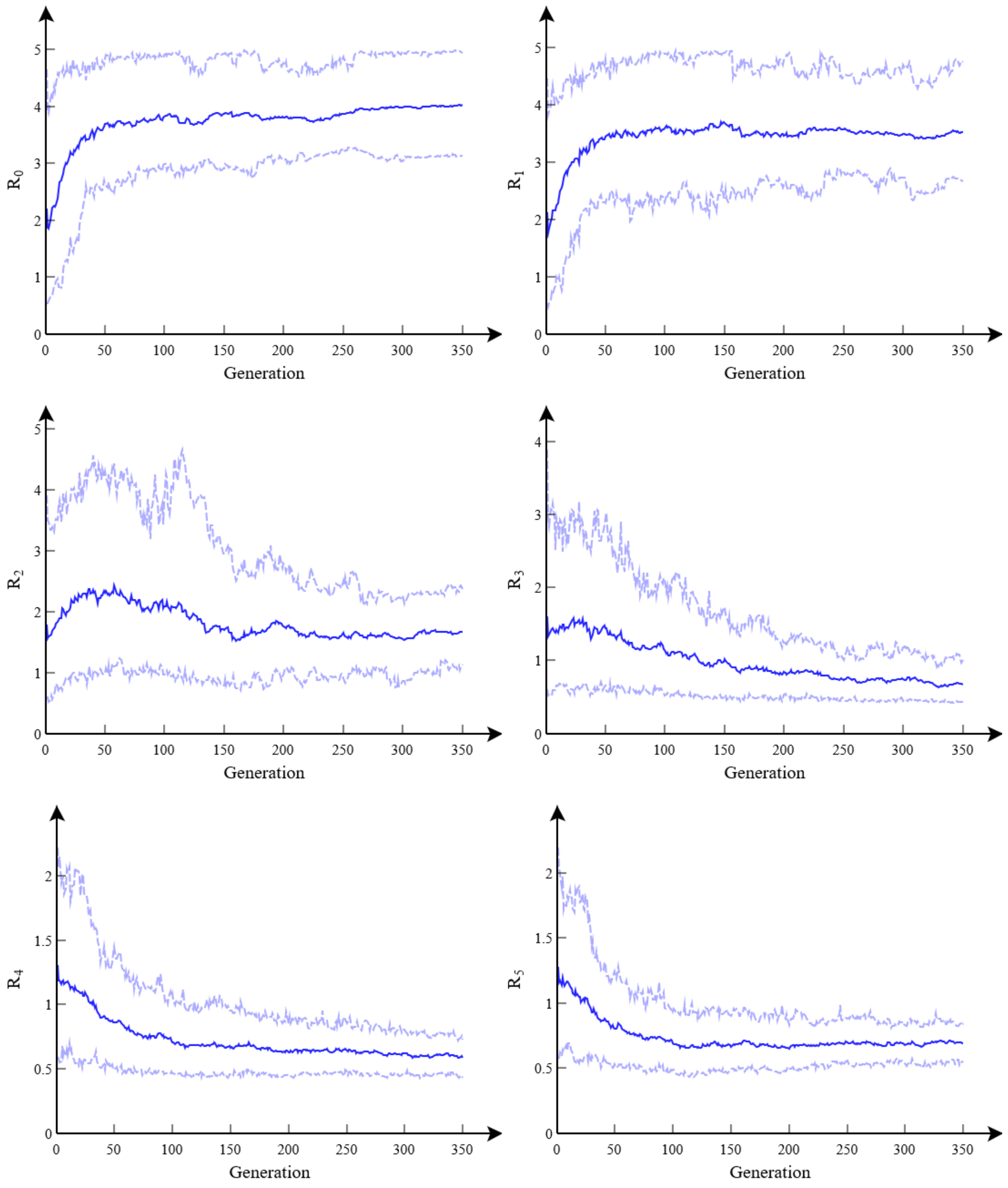


**Figure R2(m): Reproduction number $R_t$ spline points**. This shows how posterior estimates for different model parameters vary with generation number $g$ (up to a maximum $G$=350 used for the final posterior estimates). These graphs combine together $R$=16 separate runs each with $P$=16 particles. Blue lines show posterior means, with light blue dashed lines denoting 95% credible intervals.

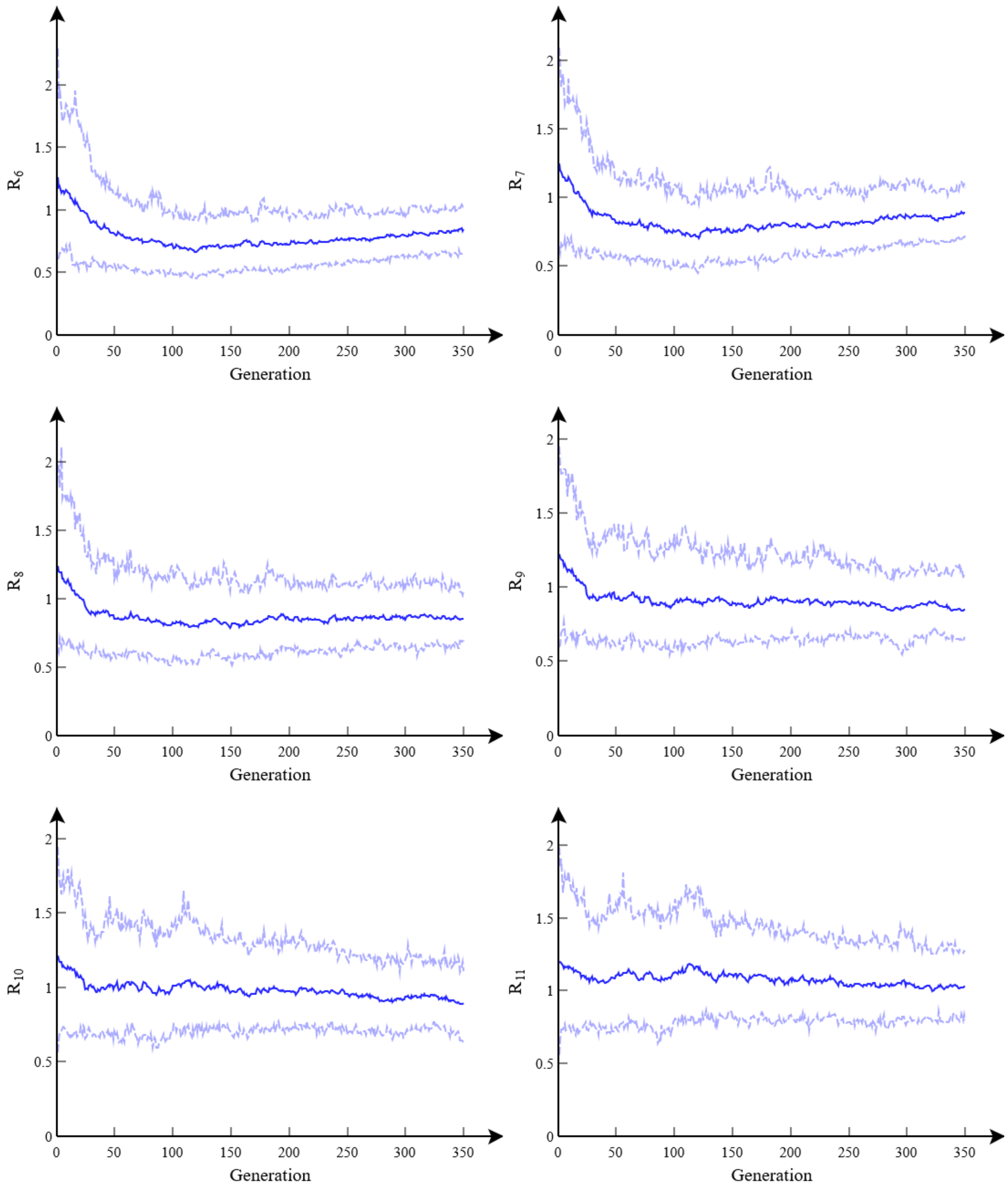


**Figure R2(n): Reproduction number $R_t$ spline points**. This shows how posterior estimates for different model parameters vary with generation number *g* (up to a maximum *G*=350 used for the final posterior estimates). These graphs combine together *R*=16 separate runs each with *P*=16 particles. Blue lines show posterior means, with light blue dashed lines denoting 95% credible intervals.

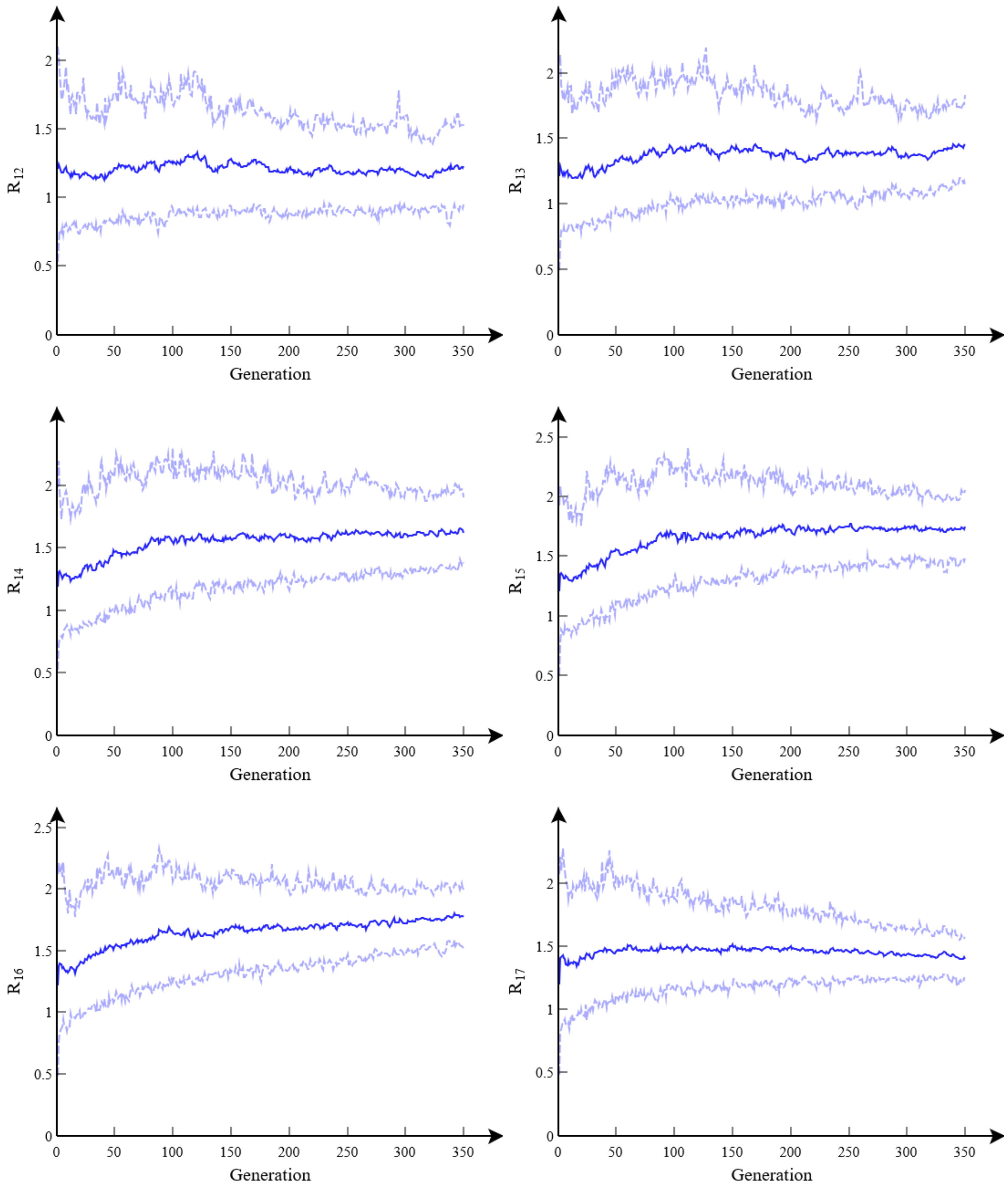


**Figure R2(o): Reproduction number $R_t$ spline points**. This shows how posterior estimates for different model parameters vary with generation number $g$ (up to a maximum $G$=350 used for the final posterior estimates). These graphs combine together $R$=16 separate runs each with $P$=16 particles. Blue lines show posterior means, with light blue dashed lines denoting 95% credible intervals.

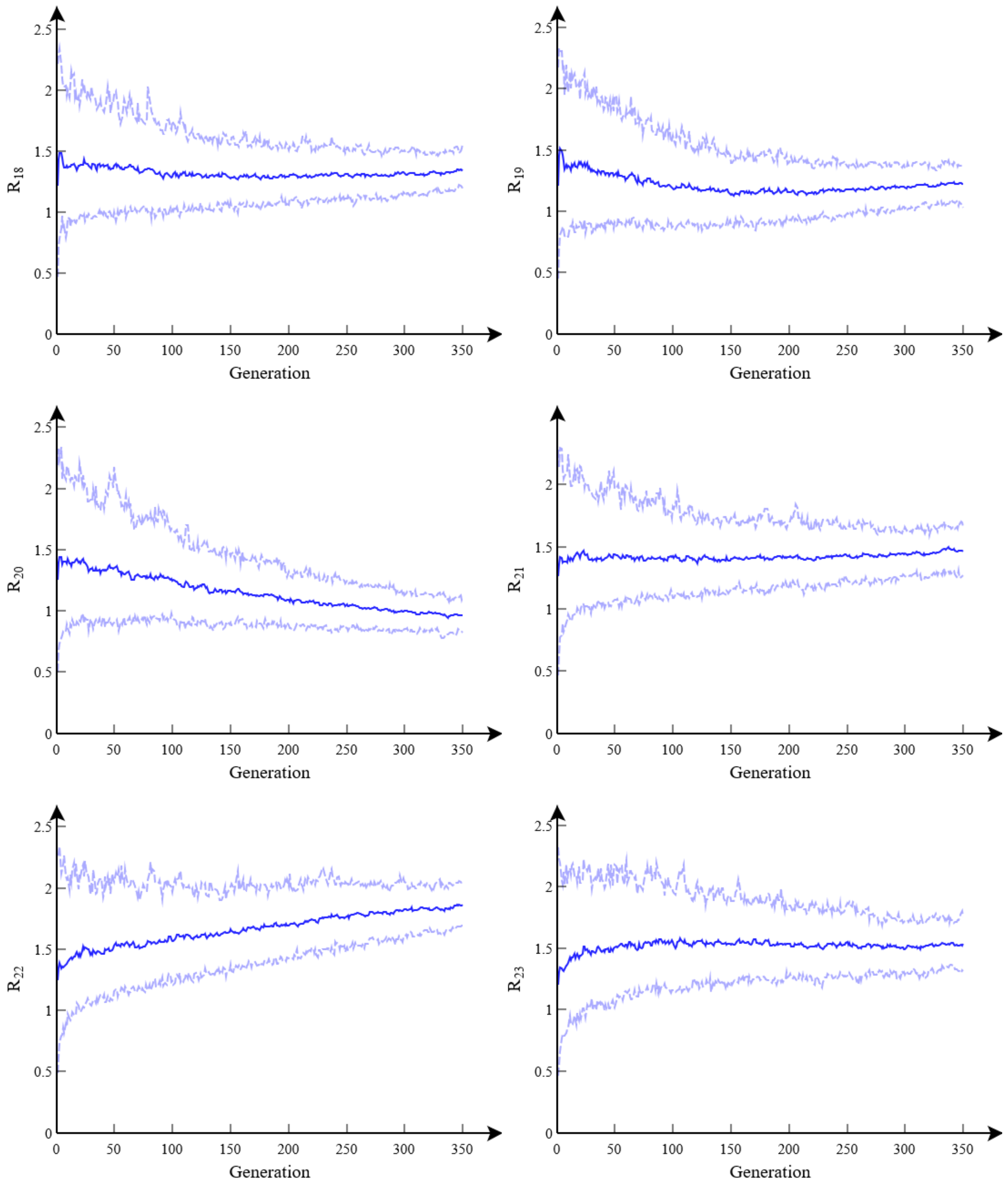


**Figure R2(p): Reproduction number $R_t$ spline points**. This shows how posterior estimates for different model parameters vary with generation number $g$ (up to a maximum $G$=350 used for the final posterior estimates). These graphs combine together $R$=16 separate runs each with $P$=16 particles. Blue lines show posterior means, with light blue dashed lines denoting 95% credible intervals.

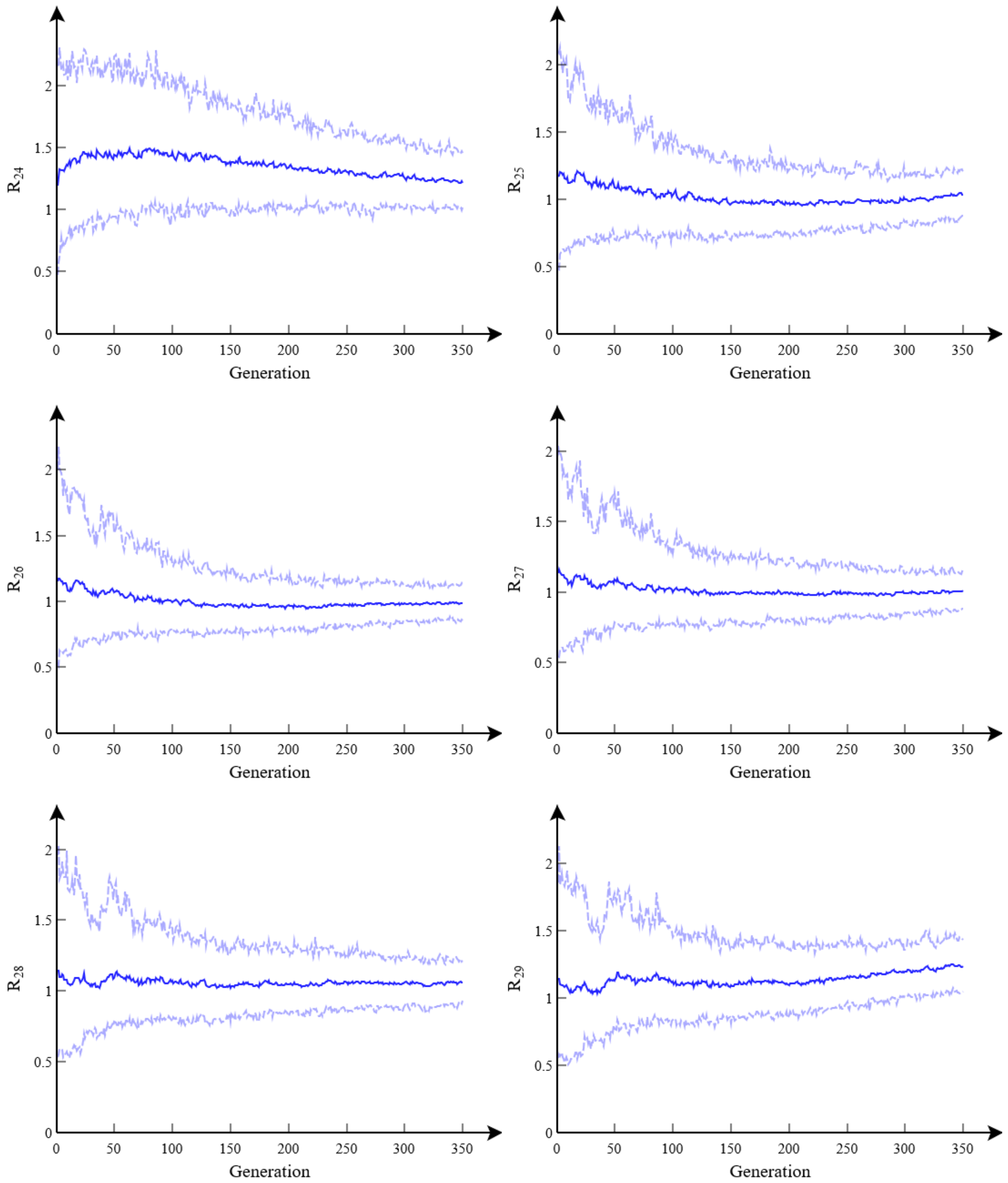


**Figure R2(q): Reproduction number $R_t$ spline points**. This shows how posterior estimates for different model parameters vary with generation number $g$ (up to a maximum $G$=350 used for the final posterior estimates). These graphs combine together $R$=16 separate runs each with $P$=16 particles. Blue lines show posterior means, with light blue dashed lines denoting 95% credible intervals.

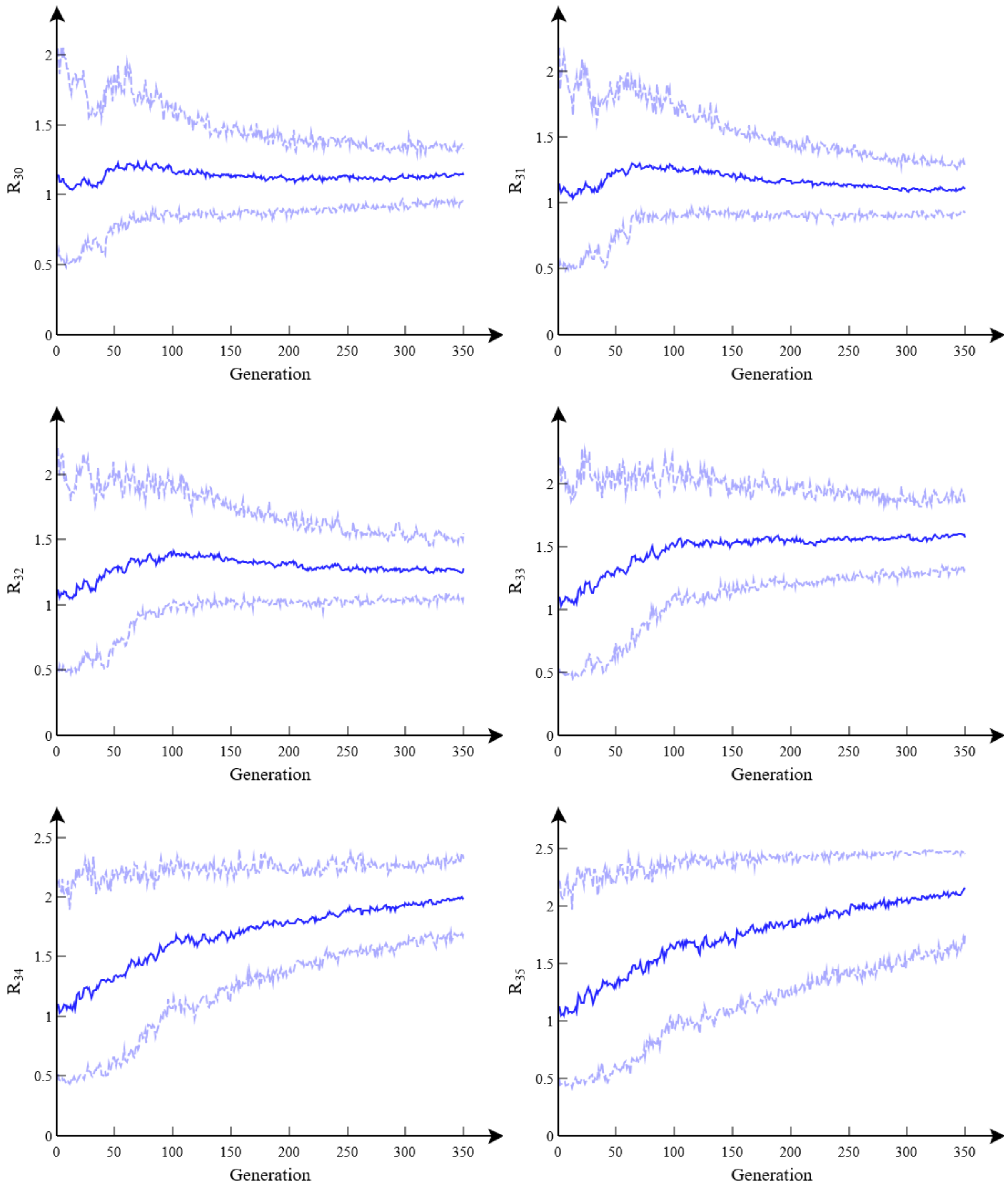


**Figure R2(r): Reproduction number $R_t$ spline points**. This shows how posterior estimates for different model parameters vary with generation number $g$ (up to a maximum $G$=350 used for the final posterior estimates). These graphs combine together $R$=16 separate runs each with $P$=16 particles. Blue lines show posterior means, with light blue dashed lines denoting 95% credible intervals.

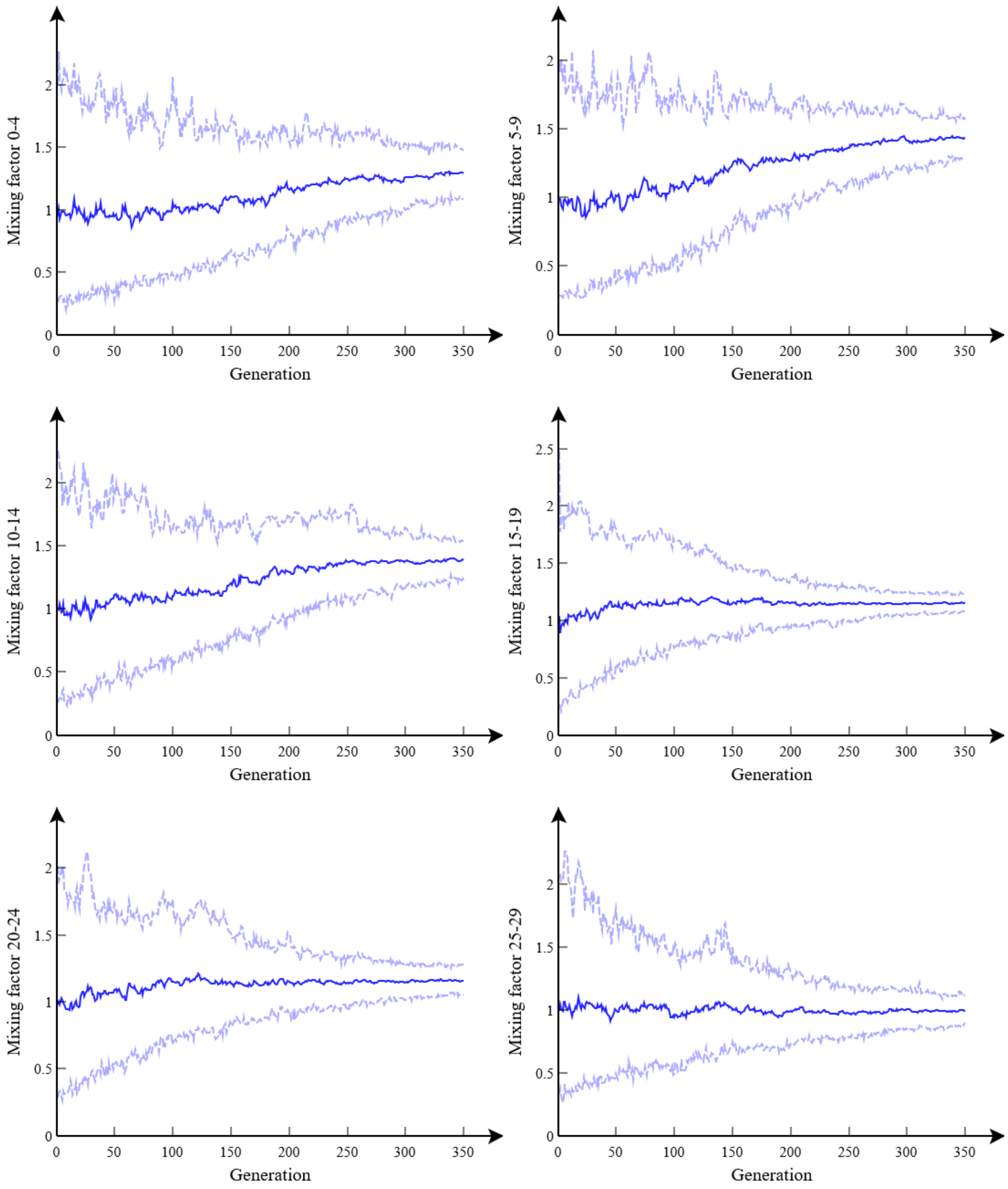


**Figure R2(s): Age contact factors $v_a$.** This shows how posterior estimates for different model parameters vary with generation number $g$ (up to a maximum $G$=350 used for the final posterior estimates). These graphs combine together $R$=16 separate runs each with $P$=16 particles. Blue lines show posterior means, with light blue dashed lines denoting 95% credible intervals.

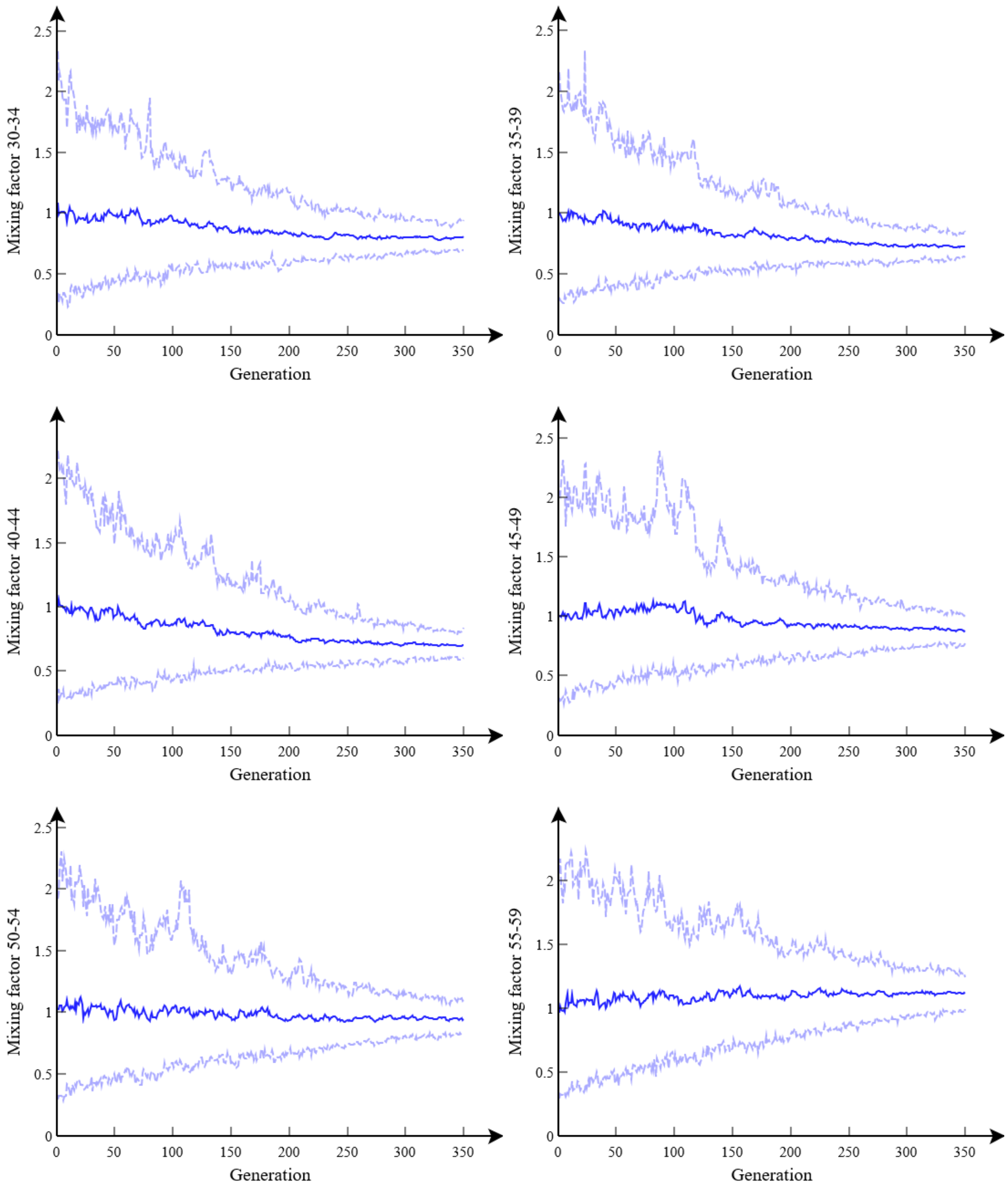


**Figure R2(t): Age contact factors $v_a$.** This shows how posterior estimates for different model parameters vary with generation number $g$ (up to a maximum $G$=350 used for the final posterior estimates). These graphs combine together $R$=16 separate runs each with $P$=16 particles. Blue lines show posterior means, with light blue dashed lines denoting 95% credible intervals.

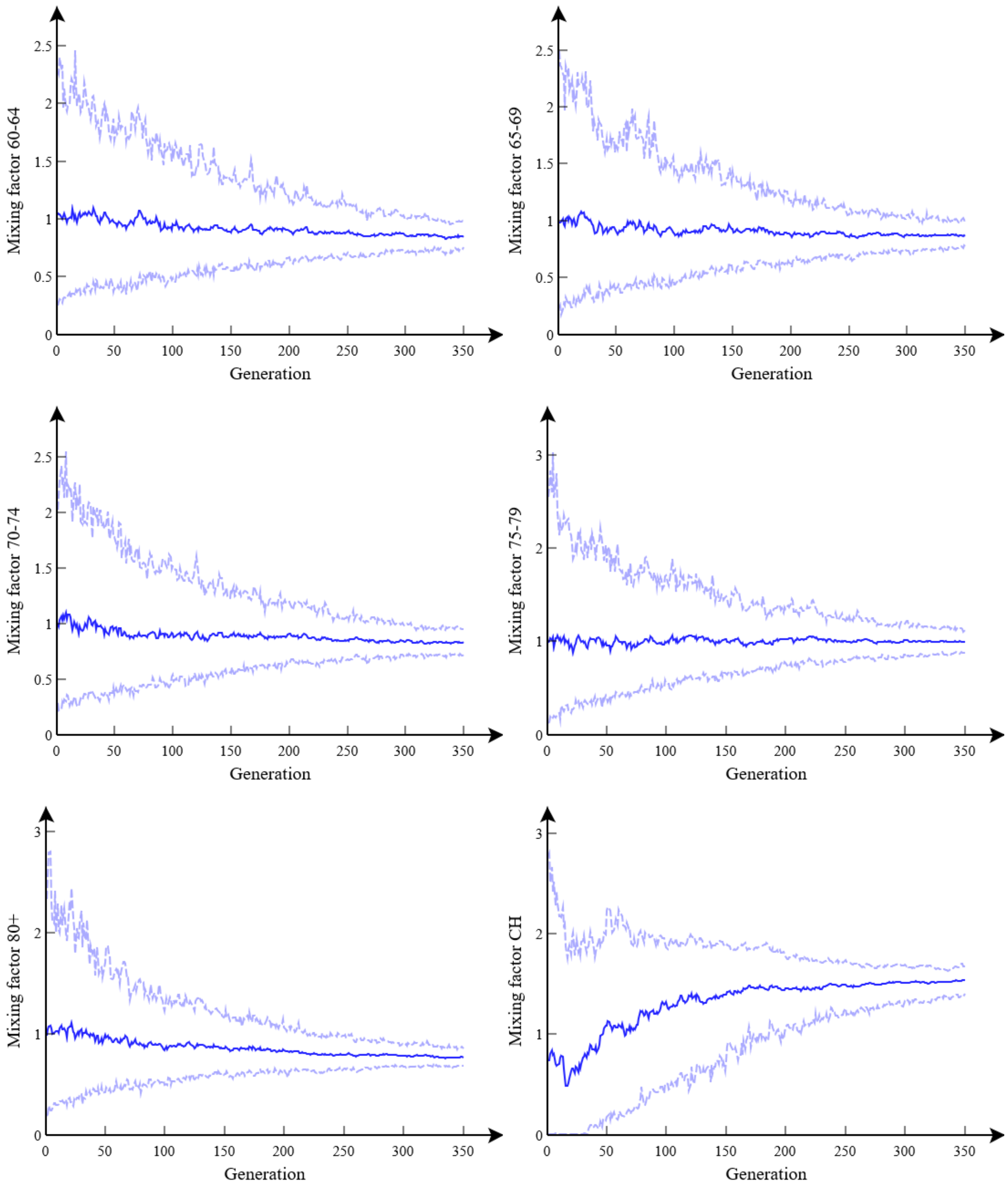


**Figure R2(u): Age contact factors $v_a$.** This shows how posterior estimates for different model parameters vary with generation number $g$ (up to a maximum $G$=350 used for the final posterior estimates). These graphs combine together $R$=16 separate runs each with $P$=16 particles. Blue lines show posterior means, with light blue dashed lines denoting 95% credible intervals.
